# The Interaction Effects of Age, APOE and Common Environmental Risk Factors on Human brain Structure

**DOI:** 10.1101/2022.10.30.22281690

**Authors:** Jie Chen, Tengfei Li, Bingxin Zhao, Hui Chen, Changzheng Yuan, Gwenn A. Gardern, Guorong Wu, Hongtu Zhu

## Abstract

Mounting evidence suggests considerable diversity in brain aging trajectories, primarily arising from the complex interplay between age, genetic and environmental risk factors, leading to distinct patterns of micro- and macro-cerebral aging. The underlying mechanisms of such effects still remain unclear. We conducted a comprehensive association analysis between cerebral structural measures and prevalent risk factors, using data from 35,035 UK Biobank subjects aged 44-82. Participants were assessed for brain volume, white matter diffusivity, Apolipoprotein E (*APOE*) genotypes, polygenic risk scores, lifestyles and socioeconomic status. We examined genetic and environmental effects and their interactions with age and sex, and identified 726 signals, with education, alcohol, and smoking affecting most brain regions. Our analysis revealed negative age-*APOE*-ε4 and positive age-*APOE*-ε2 interaction effects, respectively, especially in females on the volume of amygdala, positive age-sex-*APOE*-ε4 interaction on the cerebellar volume, positive age-excessive-alcohol interaction effect on the mean diffusivity of the splenium of the corpus callosum, positive age-healthy-diet interaction effect on the paracentral volume, and negative *APOE*-ε4-moderate-alcohol interaction effects on the axial diffusivity of the superior fronto-occipital fasciculus. These findings highlight the need of considering age, sex, genetic and environmental joint effects in elucidating normal or abnormal brain aging.

## Introduction

Aging is a complex biological process involving numerous molecular, metabolic, and cellular pathway alterations (1–3). Brain aging, characterized by age-related changes in brain structure such as regional morphometrics and white matter (WM) tract wiring patterns, plays a crucial role in reflecting and mediating aging risk factors (4–7). A wealth of literature exists on human brain aging, with common findings including diminishing brain volumes, expanding ventricles, and cognitive decline (8), loss of WM microstructural integrity (9), and brain regions particularly susceptible to aging-related changes such as frontal and temporal cortex, putamen, thalamus and accumbens (10). However, the heterogeneity in aging trajectories is complex in terms of brain structure, brain function, and presence or absence of specific cognitive and behavioral symptoms (11–13). These heterogeneities primarily stem from a wide range of demographic, genetic and environmental exposures, including non-modifiable factors such as age (14), sex (15–17) and genetics (18–20), and modifiable factors like lifestyle choices (21–24) and socioeconomic status (SES) (25, 26). These risk factors collectively and cumulatively affect an individual’s brain structure. Current studies primarily focus on individual risk factors, lacking a comprehensive understanding of brain structural regulation within this multifactorial biological system. Thus, it is essential to investigate how the interplay of these factors influences brain health and aging disparities.

Age, sex, and Apolipoprotein E (*APOE*) genotypes are three major non-modifiable risk factors that have been extensively studied for their impact on human brain aging. Age serves as a fundamental reference point for examining changes that occur over time. Research has shown that *APOE*-related effects display sex-dependent (17, 27, 28) and age-dependent associations. One longitudinal investigation (29) discovered that *APOE*-ε4 carriers exhibit increased annual atrophy in brain temporal regions, highlighting a significant age-*APOE*-ε4 interaction. In contrast, another study (30) found no statistically significant interactions of *APOE*-ε4 with age on brain structure in a cross-sectional analysis involving 8,395 UK Biobank (UKB) subjects. Furthermore, research (31) has reported no age-*APOE*-ε4 interactions on WM diffusivity. While other investigations (32) have examined the association of *APOE*-ε2 with aging-related clinical outcomes, the existing literature does not report associations on interactions of *APOE*-ε2 with age. In response to these findings, one large-scale brain-wide association study is being conducted to explore the synergistic effects between age, sex, and *APOE-*ε2/ -ε4 genotypes.

Beyond the APOE gene, numerous other genetic factors may impact brain aging. One approach to account for these factors is through the polygenic risk score (PRS), a cumulative index that synthesizes the effects of numerous common genetic variants on a particular trait or disorder. PRSs have been used to investigate the associations of brain structure and various brain disorders, such as schizophrenia (SCH) (33–36) or Alzheimer’s disease (AD) (37). In this study, to differentiate the potential impacts of PRS and APOE on brain aging, we extend our analyses beyond the *APOE* gene by including PRSs from a wide array of brain diseases and behavioral factors. These factors encompass AD (38), attention deficit hyper-activity disorder (ADHD) (39), anxiety disorder (40), Autism spectrum disorder (ASD) (41), bipolar disorder (42), major depression disorder (MDD) (43), obsessive-compulsive disorder (OCD) (44), Panic disorder (PD) (45), SCH (46), and Tourette syndrome (TS) (47).

In addition to the non-modifiable factors discussed above, numerous modifiable factors, such as lifestyle factors and SES, contribute to brain diseases and structural changes. The *2020 report of the Lancet Commission on dementia prevention, intervention, and care* estimated that 40% of dementia cases could potentially be prevented or delayed through modifications of 12 major risk factors. Among these lifestyle factors, smoking, alcohol consumption, healthy diet and physical activity have been extensively studied, as they are closely related to the risk of cardiovascular and neurodegenerative diseases.

Smoking has been identified as a well-established cardiovascular risk factor (48–50) and is associated with risks of brain disorders (51, 52) and accelerated brain aging (53–56). Similarly, heavy alcohol intake has been extensively researched and linked to cardiovascular disease (57, 58) as well as negative impact on brain health, such as accelerated brain volume loss (59) and the loss of WM integrity (24). However, the impact of moderate alcohol consumption on brain aging is more complex and remains a subject of debate.

Moderate alcohol consumption, defined as up to one drink per day for women or up to two drinks per day for men, has been associated with a lower risk of cardiovascular disease due to its potential effects on high-density lipoprotein cholesterol levels (60, 61) and possible anti-inflammatory and antithrombotic properties (62). Since cardiovascular health is closely related to brain health, it is important to consider the potential impact of moderate alcohol consumption on brain structure. The findings in this area have been controversial, potentially due to differences in study design, sample size or population characteristics. Some studies have reported moderate alcohol intake as being positively associated with whole brain volume (WBV) in elderly subjects (63), while others found no effect (64) or even adverse effect (65) on brain outcomes or cognition with moderate alcohol consumption. Study (66) reported moderate alcohol consumption could be associated with greater cognitive decline in learning and memory only among *APOE-ε4* carriers, suggesting genetic risk factors may modify the effects of lifestyle choices.

Although numerous studies have explored the effects of diet on overall brain health and cognitive decline (23, 67–70), research into the impact of diet on specific brain regions, such as the visual and sensorimotor areas remains sparse. Moreover, there is a lack of research examining the age-diet interactions. Additionally, other modifiable factors such as exercise, sleep quality, and education, also contribute to brain aging. A comprehensive understanding of the combined impact of these factors is essential for elucidating the interactions between modifiable and non-modifiable factors, as well as their relationship with age on brain structure. However, research in this domain remains limited.

The purpose of this study is to conduct a comprehensive association analysis of genetic factors, lifestyle factors, SES, age and sex with brain structure. To measure brain structure, we generated 101 brain regional volumes and 110 diffusion tensor imaging (DTI) measures of WM integrity based on the magnetic resonance imaging (MRI) data of more than 35,035 UKB subjects. We also extracted relevant risk factors of interest, including *APOE* genotypes, 16 PRSs, demographic information, lifestyle factors, and SES from these UKB subjects. **Fig. 1** depicts the study design, which integrates multi-modal data sources. The large sample size of UKB enables association analysis with high statistical power and replicability. Findings may provide new insights into the nature and timing of dementia risks underlying various health disparity conditions.

**Figure 1.**
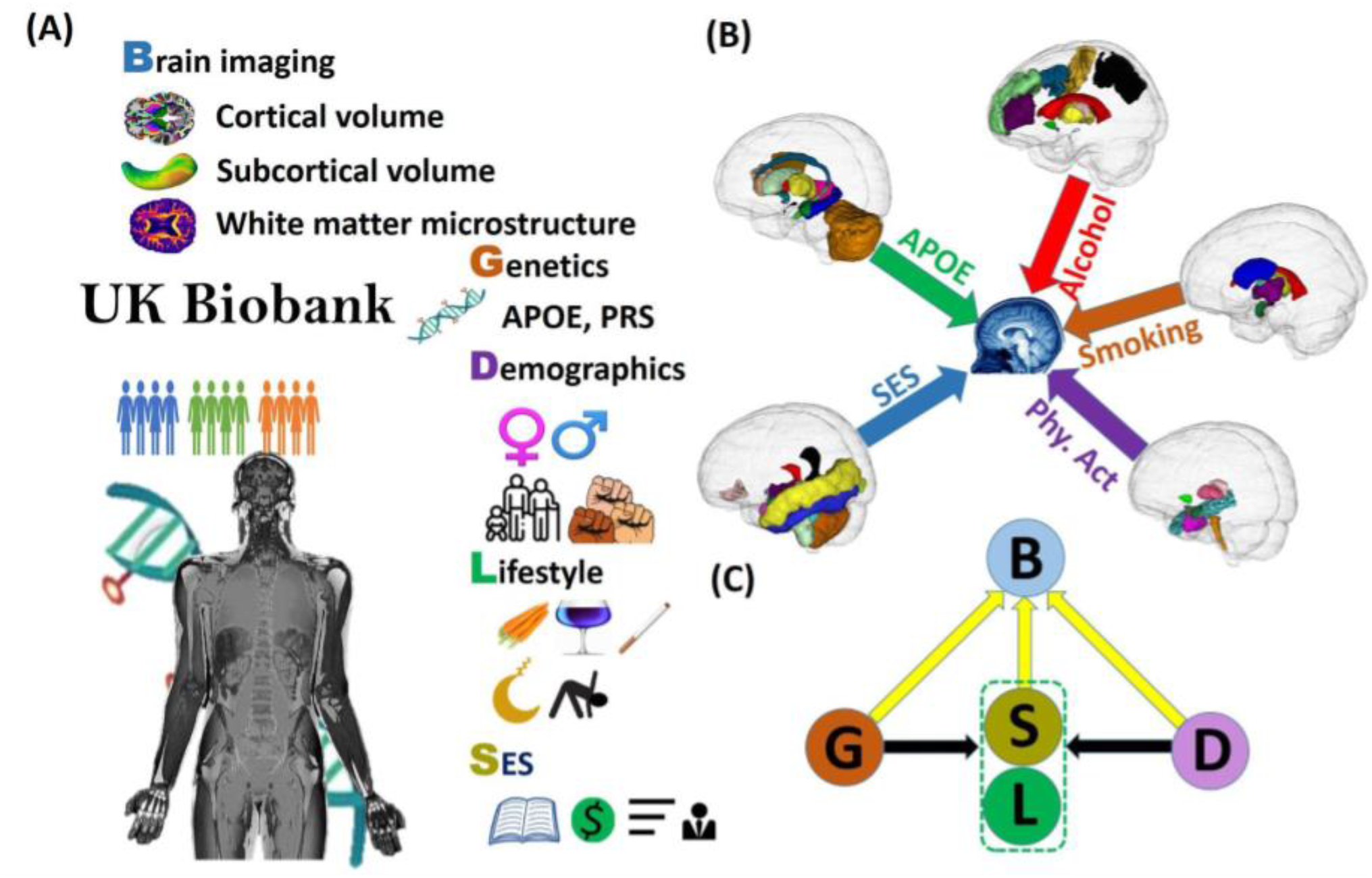
The study design. **(A)** Multi-omiics data in the UKB. **(B)** ldenti1fied brain regions with structural changes associated with APOE, alcohol, smoking, physical activity (Phy. Act.), and socioeconomic status (SES). **(C)** The causal graph among the genetics, demographics, lifestyle and SES factors and the brain structural traits.

## Results

In this study, we included 36,969 unrelated participants aged 44-82 for brain volume analysis, of which 34,097 were white British. For WM diffusivity analyses, we included 35,035 participants, with 32,320 being white British. The sample of 35,035 participants comprised 16,515 (47%) males and 18,520 (53%) females. A total of 9,191 (26%) individuals carried one *APOE*-ε4 allele, while 792 (2%) carried two. Demographic information is summarized in **Table 1**. Correlation plots for genetic factors, environmental (lifestyle and SES) factors, 101 region-of-interest (ROI) phenotypes, and 110 DTI traits (***<u>SI Appendix</u>*, <u>Figs. S1-S3</u>**) indicated strong mutual correlations between brain structural traits, while correlations between environmental factors were much weaker (< 0.1).

**Table 1.**
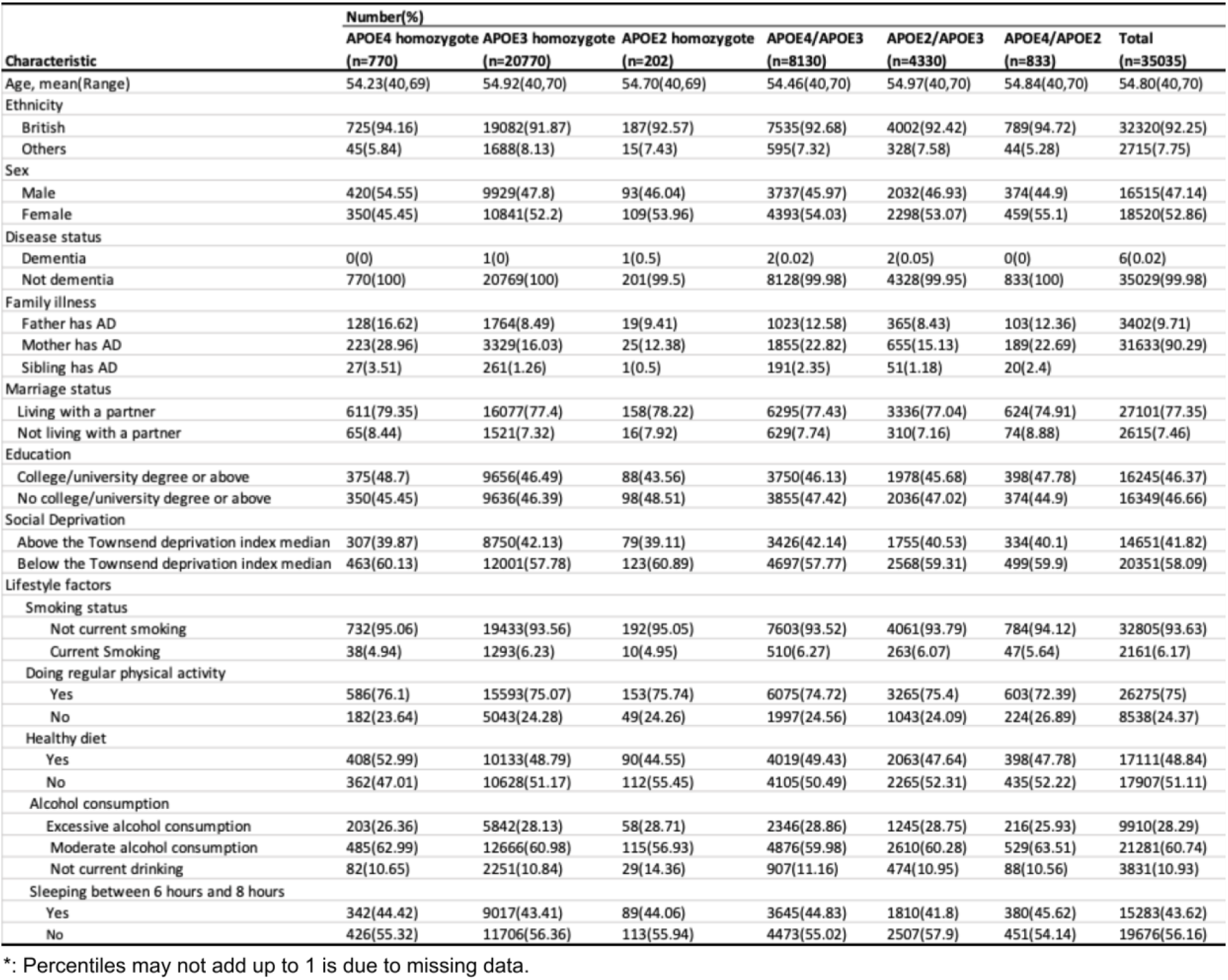
Demographic information, lifestyle factors and polygenic risk scores of the UK Biobank subjects used in the association studies on diffusion tensor imaging measurements. AD, Alzheimer’s Disease.

| Characteristic | Number(%) |  |  |  |  |  |  |
| --- | --- | --- | --- | --- | --- | --- | --- |
|  | APOE4 homozygote<br>(n=770) | APOE3 homozygote<br>(n=20770) | APOE2 homozygote<br>(n=202) | APOE4/APOE3<br>(n=8130) | APOE2/APOE3<br>(n=4330) | APOE4/APOE2<br>(n=833) | Total<br>(n=35035) |
| Age, mean(Range) | 54.23(40,69) | 54.92(40,70) | 54.70(40,69) | 54.46(40,70) | 54.97(40,70) | 54.84(40,70) | 54.80(40,70) |
| Ethnicity |  |  |  |  |  |  |  |
| British | 725(94.16) | 19082(91.87) | 187(92.57) | 7535(92.68) | 4002(92.42) | 789(94.72) | 32320(92.25) |
| Others | 45(5.84) | 1688(8.13) | 15(7.43) | 595(7.32) | 328(7.58) | 44(5.28) | 2715(7.75) |
| Sex |  |  |  |  |  |  |  |
| Male | 420(54.55) | 9929(47.8) | 93(46.04) | 3737(45.97) | 2032(46.93) | 374(44.9) | 16515(47.14) |
| Female | 350(45.45) | 10841(52.2) | 109(53.96) | 4393(54.03) | 2298(53.07) | 459(55.1) | 18520(52.86) |
| Disease status |  |  |  |  |  |  |  |
| Dementia | 0(0) | 1(0) | 1(0.5) | 2(0.02) | 2(0.05) | 0(0) | 6(0.02) |
| Not dementia | 770(100) | 20769(100) | 201(99.5) | 8128(99.98) | 4328(99.95) | 833(100) | 35029(99.98) |
| Family illness |  |  |  |  |  |  |  |
| Father has AD | 128(16.62) | 1764(8.49) | 19(9.41) | 1023(12.58) | 365(8.43) | 103(12.36) | 3402(9.71) |
| Mother has AD | 223(28.96) | 3329(16.03) | 25(12.38) | 1855(22.82) | 655(15.13) | 189(22.69) | 31633(90.29) |
| Sibling has AD | 27(3.51) | 261(1.26) | 1(0.5) | 191(2.35) | 51(1.18) | 20(2.4) |  |
| Marriage status |  |  |  |  |  |  |  |
| Living with a partner | 611(79.35) | 16077(77.4) | 158(78.22) | 6295(77.43) | 3336(77.04) | 624(74.91) | 27101(77.35) |
| Not living with a partner | 65(8.44) | 1521(7.32) | 16(7.92) | 629(7.74) | 310(7.16) | 74(8.88) | 2615(7.46) |
| Education |  |  |  |  |  |  |  |
| College/university degree or above | 375(48.7) | 9656(46.49) | 88(43.56) | 3750(46.13) | 1978(45.68) | 398(47.78) | 16245(46.37) |
| No college/university degree or above | 350(45.45) | 9636(46.39) | 98(48.51) | 3855(47.42) | 2036(47.02) | 374(44.9) | 16349(46.66) |
| Social Deprivation |  |  |  |  |  |  |  |
| Above the Townsend deprivation index median | 307(39.87) | 8750(42.13) | 79(39.11) | 3426(42.14) | 1755(40.53) | 334(40.1) | 14651(41.82) |
| Below the Townsend deprivation index median | 463(60.13) | 12001(57.78) | 123(60.89) | 4697(57.77) | 2568(59.31) | 499(59.9) | 20351(58.09) |
| Lifestyle factors |  |  |  |  |  |  |  |
| Smoking status |  |  |  |  |  |  |  |
| Not current smoking | 732(95.06) | 19433(93.56) | 192(95.05) | 7603(93.52) | 4061(93.79) | 784(94.12) | 32805(93.63) |
| Current Smoking | 38(4.94) | 1293(6.23) | 10(4.95) | 510(6.27) | 263(6.07) | 47(5.64) | 2161(6.17) |
| Doing regular physical activity |  |  |  |  |  |  |  |
| Yes | 586(76.1) | 15593(75.07) | 153(75.74) | 6075(74.72) | 3265(75.4) | 603(72.39) | 26275(75) |
| No | 182(23.64) | 5043(24.28) | 49(24.26) | 1997(24.56) | 1043(24.09) | 224(26.89) | 8538(24.37) |
| Healthy diet |  |  |  |  |  |  |  |
| Yes | 408(52.99) | 10133(48.79) | 90(44.55) | 4019(49.43) | 2063(47.64) | 398(47.78) | 17111(48.84) |
| No | 362(47.01) | 10628(51.17) | 112(55.45) | 4105(50.49) | 2265(52.31) | 435(52.22) | 17907(51.11) |
| Alcohol consumption |  |  |  |  |  |  |  |
| Excessive alcohol consumption | 203(26.36) | 5842(28.13) | 58(28.71) | 2346(28.86) | 1245(28.75) | 216(25.93) | 9910(28.29) |
| Moderate alcohol consumption | 485(62.99) | 12666(60.98) | 115(56.93) | 4876(59.98) | 2610(60.28) | 529(63.51) | 21281(60.74) |
| Not current drinking | 82(10.65) | 2251(10.84) | 29(14.36) | 907(11.16) | 474(10.95) | 88(10.56) | 3831(10.93) |
| Sleeping between 6 hours and 8 hours |  |  |  |  |  |  |  |
| Yes | 342(44.42) | 9017(43.41) | 89(44.06) | 3645(44.83) | 1810(41.8) | 380(45.62) | 15283(43.62) |
| No | 426(55.32) | 11706(56.36) | 113(55.94) | 4473(55.02) | 2507(57.9) | 451(54.14) | 19676(56.16) |

\*: Percentiles may not add up to 1 is due to missing data.

We conducted a four-fold association analysis to study the effects of age, sex and *APOE* two-way interactions, age-sex-*APOE* three-way interactions, *APOE*-environment interactions, and age-environment interactions on 110 DTI measurements and 101 ROI volumes. We considered PRSs, UKB study phase, marriage status, and WBV (for ROI volume only) as additional covariates to control for potential confounders (see **Methods** and ***<u>SI Appendix</u>*, <u>Supplementary Text</u>**). Our primary findings were based on multiple linear regression models for each brain imaging trait using white British subjects, which were validated in non-British populations.

We identified a total of 331 significant associations for ROI volumes and 395 for DTI traits, including both main and interaction effects. The identified effects size (standardized coefficients) were reproducible, with correlations 0.87 (***<u>SI Appendix</u>*, <u>Fig. S15</u>**) for ROI volumes and 0.96 (**Fig. S16)** for DTI traits between British and non-British populations. Our analysis revealed that education, alcohol and smoking were the top three common environmental factors that influence most regions (***<u>SI Appendix</u>*, <u>Figs. S17 and S18</u>**).

### Genetic Associations with Brain Structural Architecture

Our investigation into the relationships between brain structure, the *APOE* gene, age, sex, and PRSs yielded several intriguing discoveries.

Of particular note, for the *APOE*-environment interaction, we identified a positive interaction (*p = 2*.*22×10-4*) between *APOE*-ε4 and moderate alcohol consumption on the axial diffusivity (AxD) of the superior fronto-occipital fasciculus (SFO) (***<u>SI Appendix</u>*, <u>Fig. S20</u>**). This positive interaction suggests that the combination of the presence of the *APOE*-ε4 gene and moderate alcohol consumption may synergistically increase the risk of losing WM integrity. No significant *APOE*-environmental interaction effect was found on brain volumes (***<u>SI Appendix</u>*, <u>Fig. S19</u>**).

A positive age-sex-*APOE*-ε4 interaction (*p = 1*.*9E-04*) was observed in relation to volumetric variations in the left cerebellum. Specifically, we observed that among *APOE*-ε4 carriers, the volume of the left cerebellar exterior decreases at a slower rate in males than in females (**Fig. 2B**). Furthermore, we found a negative age-*APOE*-ε4 interaction effect on the volume of amygdala only in females (***<u>SI Appendix</u>*, <u>Fig. S5</u>**). These findings support prior studies (27) suggesting that female carriers of *APOE*-ε4 have a heightened susceptibility to AD. In contrast, among *APOE*-ε4 non-carriers, negative age-sex interactions were widely observed on the volume of WM, grey matter (GM), and the majority of cortical and subcortical regional volumes (***<u>SI Appendix</u>*, <u>Fig. S7</u>**), suggesting accelerated brain volume reduction among males as opposed to females in the general population. This implied that the effect of sex diverges between *APOE*-ε4 carriers and non-carriers.

**Figure 2.**
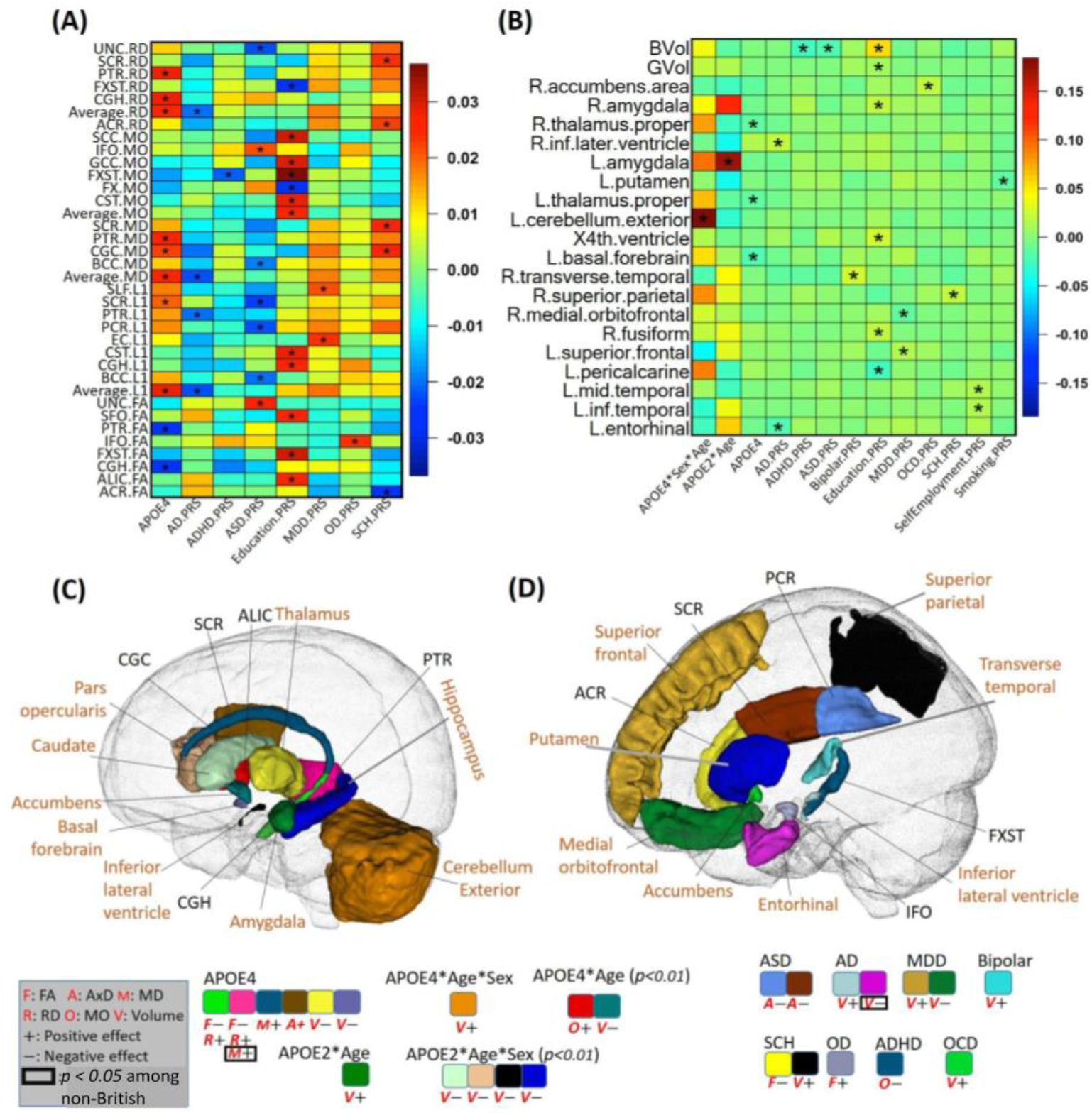
Results on genetic components. The heatmaps display standardized coefficients of the genetic (APOE, PRS) and age, sex, and gene-related interaction effects on white matter diffusivity **(A)** and volume **(B)**. The identified brain regions and signals on volume (shown with “V”) and diffusivity (shown with “F”, “A”, **“M”**, “R” and “O” for FA, AxD, MD, RD and MO, respectively) are shown for APOE-related effects **(C)** and PRS-related effects **(D)**. An asterisk (*) denotes the *2*.*37E-4* significance level. In **(C)** and **(D)**, we display the statistical association results consistent between British and non-British ancestry populations. Imaging phenotypes at particular brain regions with *p* < *0*.*05* in non-British ancestry populations are marked by black box. Colors indicate brain regions, while the direction of effects is denoted by “+” for positive and “-” for negative. PRS, polygenic risk score; FA, fractional anisotropy; AxD, axial diffusivity; MD, mean diffusivity; RD, radial diffusivity; MO, mode of diffusivity.

More interestingly, we found a positive age-*APOE*-ε2 interaction effect on left amygdala volume (*p = 4*.*84E-06*), while for right amygdala volume, such effects were only present in females **(Figs. 2B** and **2C** and ***<u>SI Appendix</u>*, <u>Figs. S5 and S12</u>)**. This suggests that the directions of *APOE*-ε4 and *APOE*-ε2 interactions with age are opposite for amygdala volume, and both interactions are more pronounced in females.

In terms of well-established findings, our results corroborate the association of *APOE*-ε4 with reduced volumes of thalamus and basal forebrain, as well as the fractional anisotropy (FA) of the cingulum hippocampus (CGH) and posterior thalamic radiation (PTR). Additionally, *APOE*-ε4 was positively associated with the radial diffusivity (RD), mean diffusivity (MD), and AxD of the CGH, PTR, superior corona radiata (SCR), and cingulum cingulate (CGC) (**Figs. 2A, 2B** and **2C**). Such findings regarding the main effect of *APOE*-ε4 align well with previous studies <u>(71, 72)</u>.

As for the PRS, we discovered that 15 PRSs were associated with brain structure (**Figs. 2A, 2B**, and **2D**). Specifically, for the larger AD PRS, we observed an increase in ventricular volume (inferior lateral ventricle, *p = 1*.*1E-4*) and a decrease in the volume of the entorhinal cortex (*p = 4*.*8E-5*). This suggests that AD-related genetic information, beyond the *APOE* gene, also contributes to changes in brain structure.

### Associations between smoking and Brain Structure

No interaction effects between smoking and age were observed, indicating that the impact of smoking on brain aging within this age range (44-82) may predominantly manifest as alterations to baseline brain structure values rather than influencing the rate of change over time.

Smoking negatively affected overall GM volumes (*p = 2*.*6E-22*), and average FA (*p = 4*.*5E-9*), while increasing average RD (*p = 7*.*5E-11*) and MD (*p = 2*.*0E-8*). The identified associations included four subcortical regions (the thalamus, amygdala, putamen, and ventral diencephalon), three ventricular regions (the third ventricle, lateral ventricle, and inferior lateral ventricle), and 16 WM tracts (**Figs. 3A** and **4B**).

**Figure 3.**
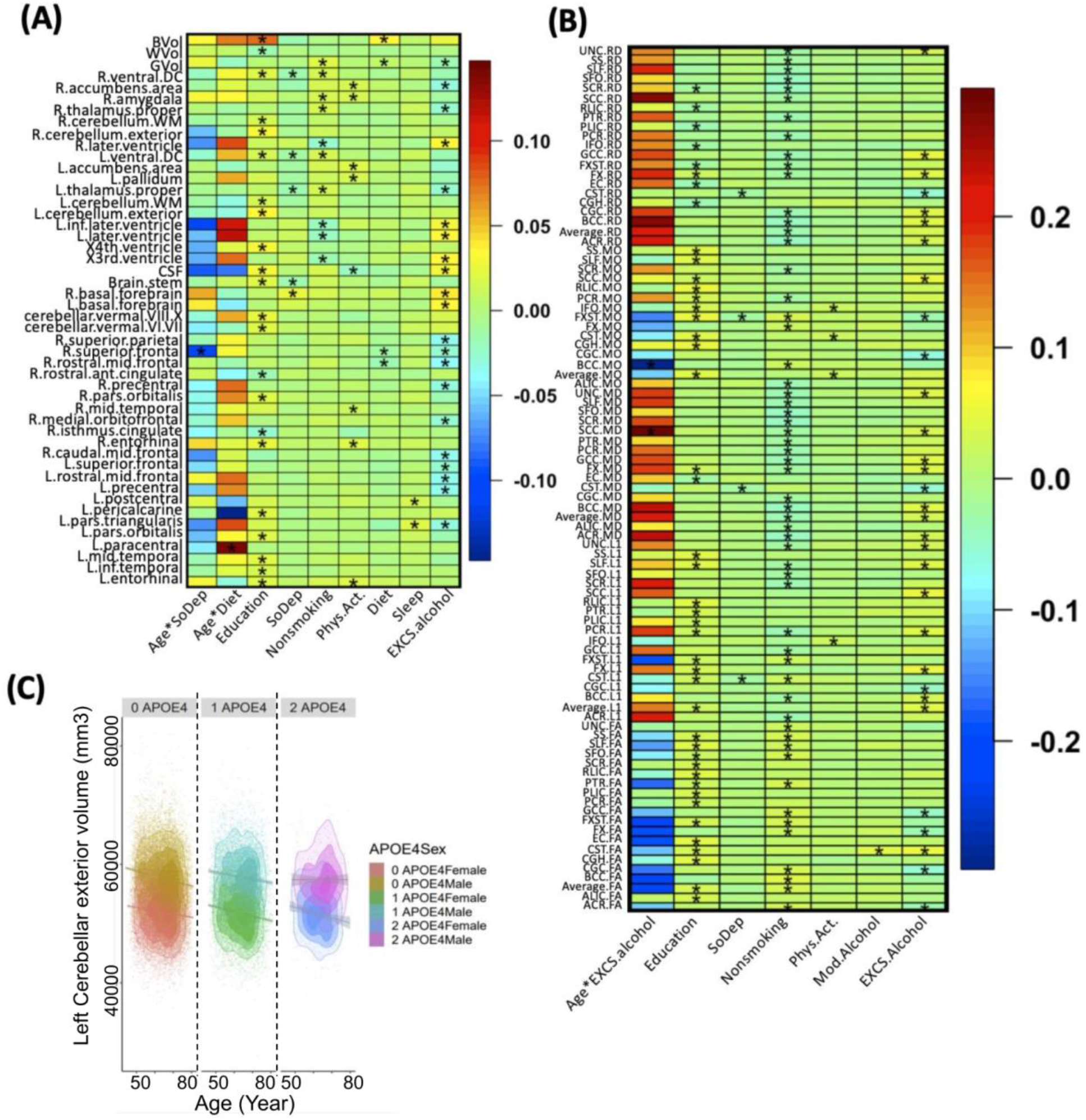
Heatmaps of associations between lifestyle, socioeconomic status, their age-interactions and brain volumes (A) and white matter diffusion tensor imaging measurements (B). We use (*) to denote the significant effect with *p* < *2*.*37E-4*. (C) Scatterplots of left cerebellum exterior volume against age among three *APOE* groups, including, from left to right, zero *APOE*-ε, one *APOE*-ε4, and two *APOE*-ε4 allele groups. Contour plots are used to show the density of two-dimensional distributions. Regression lines and 95% confidence intervals are also displayed.

**Figure 4.**
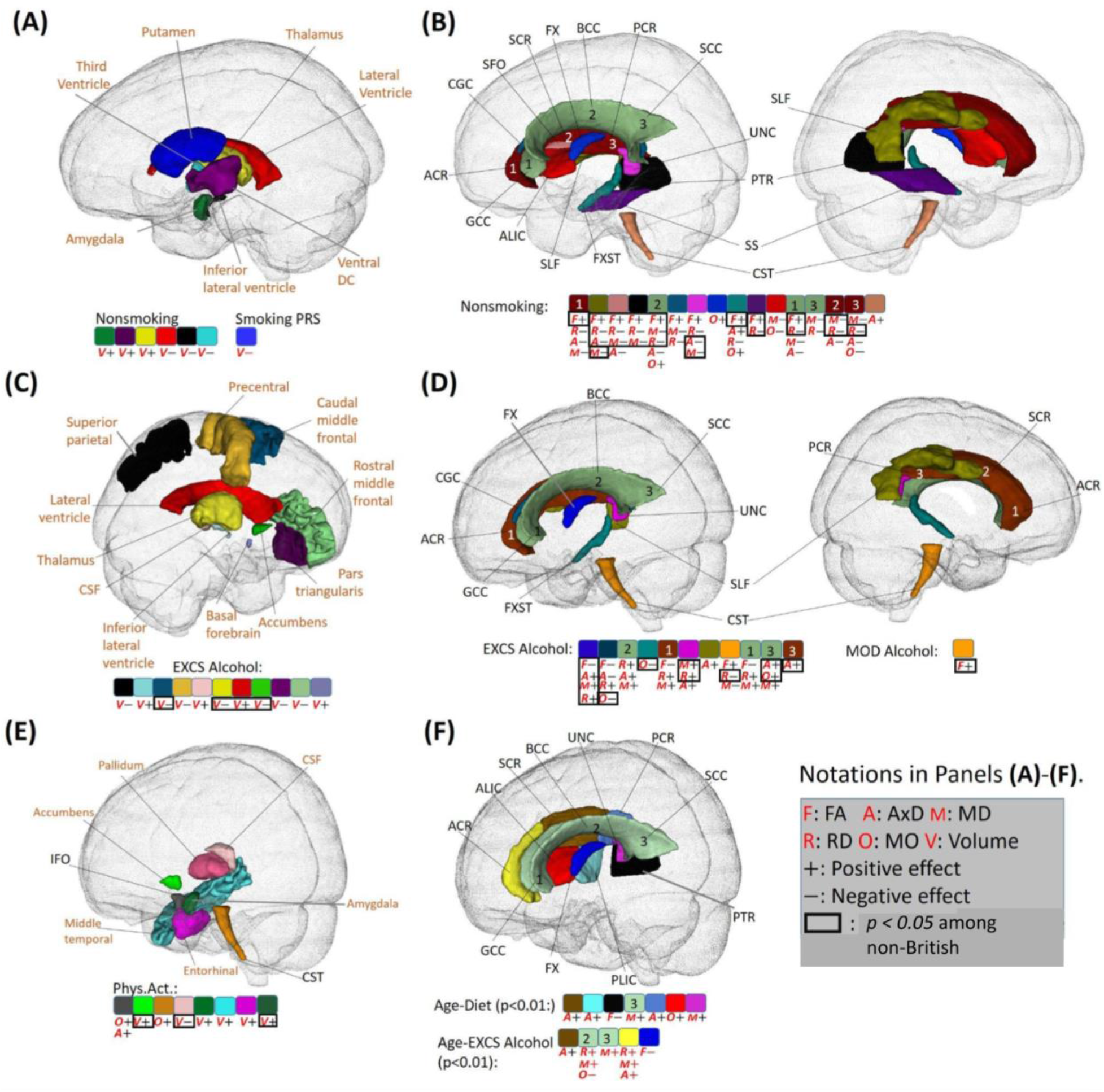
Identified brain regions with the effects of lifestyle factors. **(A)** Regions with effects of non-smoking phenotype and smoking PRS score on volume (shown with “V”; directions of effects are denoted by ”+” for positive and “-” for negative). **(B)** Regions with effects of non-smoking phenotype on white matter diffusivity (shown with “F”, “A”, “M”, “R” and “O” for FA, AxD, MD, RD and MO, respectively). **(C)** Regions with effects of excessive alcohol (EXCS Alcohol) on volume. **(D)** Regions with effects of excessive alcohol (EXCS Alcohol) on diffusivity. **(E)** Regions with effects of physical activity (Phys.Act.) on volume and diffusivity. **(F)** Regions with effects of Age-diet and Age-EXCS Alcohol interactions on diffusivity. In **(A)-(E)**, only signals with *p* < *2*.*37E-4* in the British population that are validated by non-British ancestry populations (showing the same direction) are displayed. In **(F)**, signals with p-values less than *0*.*01* in the British population that are validated by non-British ancestry populations are displayed. Signals with *p* < *0*.*05* for non-British ancestry populations are enclosed with a black rectangle. PRS, polygenic risk score; FA, fractional anisotropy; AxD, axial diffusivity; MD, mean diffusivity; RD, radial diffusivity; MO, mode of diffusivity.

The patterns of these effects were consistent across the imaging traits, including reduced subcortical volume, increased ventricular volume, decreased FA, and increased AxD, RD, and MD in WM tracts (**Figs. 3A, 3B, 4A**, and **4B**). These results align with previous research on the negative effects of smoking on brain structure (21, 73–76), particularly for the GM, thalamus, cerebellum, putamen and lateral ventricle volumes, and diffusivity parameters (77–81) on the corpus callosum (CC), anterior limb of internal capsule (ALIC), superior longitudinal fasciculus (SLF), and uncinate fasciculus (UNC).

The directions of these effects were consistent between white British and non-British ancestry racial groups. Specifically, the directions of signals were all (10 out of 10) consistent in terms of volume and 93% (53 out of 57) consistent in terms of WM diffusivity.

### Associations between Alcohol Consumption and Brain Structure

We observed a positive interaction effect between age and excessive alcohol on the mean diffusivity of the splenium of the corpus callosum (*p = 2*.*0E-4*; **Figs. 3B** and **4F**, and ***<u>SI Appendix</u>*, <u>Fig. S14</u>**). Besides, although not reaching Bonferron significance level, there were other age interactions that passed *p < 0*.*01*, including positive effects on lateral ventricle volume, and AxD, RD and MD of the corpus callosum (CC) and corona radiata (CR), and negative effects on the volumes of inferior temporal, precentral and caudate regions and FA of the fornix (FX) (**Figs. 4F** and **5C**). These findings warrant attention as they were all (6 out of 6) validated by non-British ancestry populations with the same directions. The consistency supported the notion of increased brain aging with excessive alcohol consumption.

Regarding the main effects of alcohol consumption, excessive alcohol was associated with a reduction in WBV (*p = 9*.*3E-15*) and an increase in average AxD (*p = 7*.*8E-7*). The identified associations spanned various brain regions and WM tracts, with consistent directions of reduced cortical and subcortical volumes and inflated ventricular volume, decreased FA, and increased AxD, RD, and MD (**Figs. 3, 4C**, and **4D**). Furthermore, when comparing the directions of these main effects between white British and non-British ancestry populations, they were more consistent for WM diffusivity (30 out of 31) than brain volume (11 out of 21).

### Associations between Diet, Physical Activity, Sleep and Brain Structure

Our investigations revealed several notable findings related to age-diet interactions, physical activity, and sleep.

Regarding age-diet interactions, we observed positive effects on left paracentral volume (*p = 2*.*0E-4*). Additionally, several age-diet interactions with *p < 0*.*01* were identified and validated by non-British ancestry populations. In general, we found some brain structural changes caused by diet were more evident among younger adults (ventricular and visual regions cuneus, pericalcarine) while some other regions were more affected among older adults (paracentral and pars opercularis) (**Figs. 3A** and **5C**, and ***<u>SI Appendix</u>*, <u>Fig</u>**. **<u>S13</u>**). For the main effects, individuals with higher adherence to a healthy diet exhibited an increase in WBV (*p = 9*.*3E-15*).

**Figure 5.**
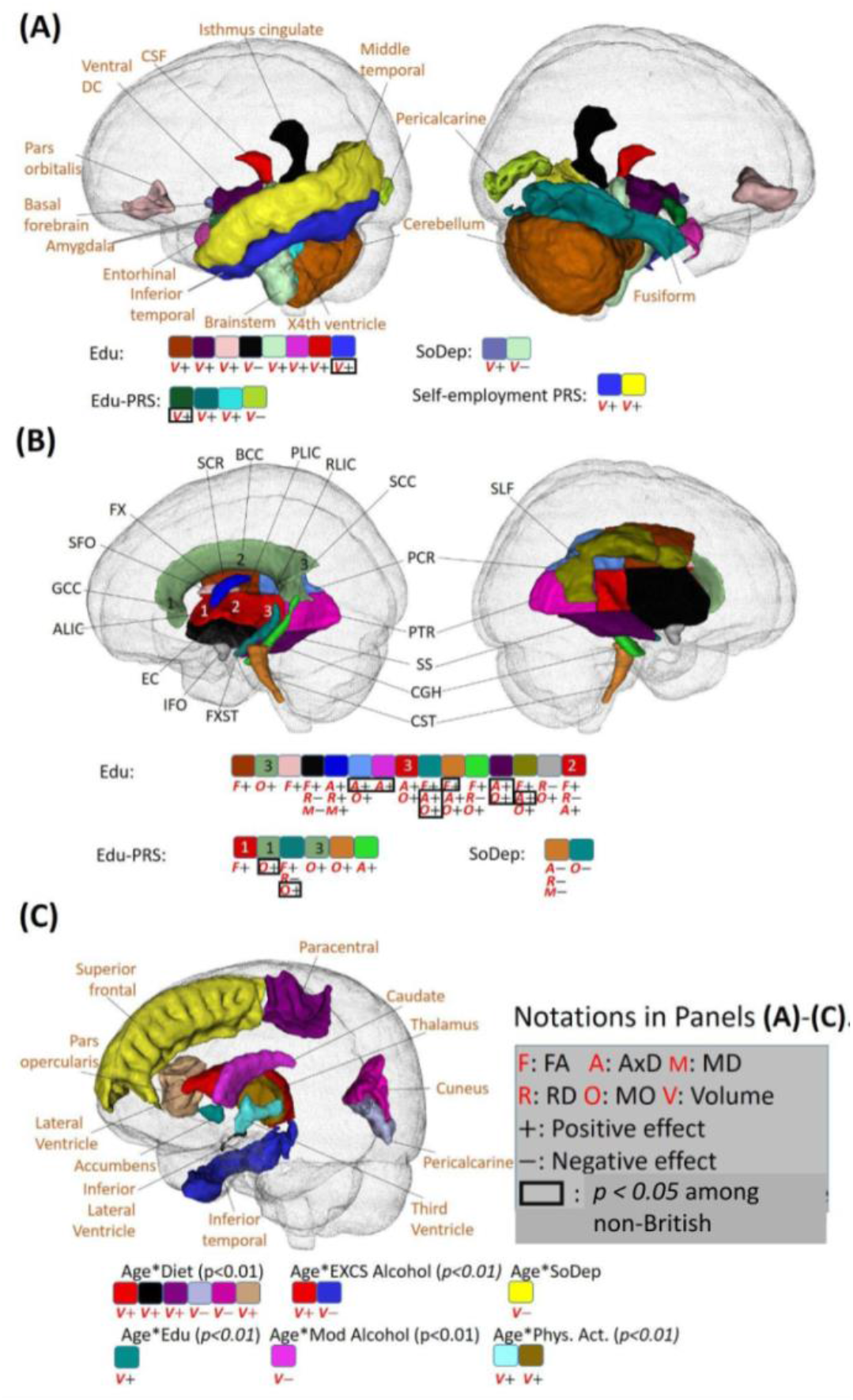
Identified brain regions with the effects of socioeconomic status (SES). **(A)** Regions with effects of SES phenotype on volume (shown with “V”; directions of effects are denoted by “+” for positive and “-” for negative). **(B)** Regions with effects of SES on white matter diffusivity (shown with “F”, “A”, “M”, “R” and “O” for FA, AxD, MD, RD and MO, respectively). **(C)** Regions with effects of age-lifestyle and age-SES interactions on volume. In **(A)** and **(B)**, signals with *p* < *2*.*37E-4* in the British ancestry population that are validated by non-British ancestry population (the same direction) are displayed. In **(C)**, the statistical association results that are consistent between British and non-British ancestry populations are displayed. Additionally, the imaging phenotypes at particular brain regions with *p* < *0*.*05* in non-British ancestry populations are marked by a black box. Colors indicate brain regions. PRS, polygenic risk score; FA, fractional anisotropy; AxD, axial diffusivity; MD, mean diffusivity; RD, radial diffusivity; MO, mode of diffusivity.

No significant age-physical activity interactions were found. For the main effects, beneficial effects of physical activity were identified, including (**Figs. 3A** and **4E**) positive associations with regional volume in temporal (middle temporal and entorhinal) and subcortical (amygdala, accumens and pallium) regions, and negative associations with the cerebrospinal fluid (CSF) volume. These findings are in line with previous literature on the entorhinal region (82), nucleus accumbens (83), and CSF (84).

Finally, we observed negative age-sleep interactions on the right superior frontal volume, suggesting that healthy sleep may benefit superior frontal regions more in young adults. In addition, healthy sleep demonstrated positive effects on the volume of the pars triangularis and postcentral regions (**Fig. 3A**). However, the directions of both those main and age-interaction effects could not be validated in non-British ancestry populations, highlighting the need for further investigations into minority populations to validate findings on sleep.

### Associations between Education and Brain Structure

We discovered inconsistencies in age-education interactions across racial groups. The directions of all identified age-interaction effects were 20.0% (1 out of 5) consistent for volume measures and 75.0% (6 out of 8) for WM diffusivity measures across racial groups (**<u>SI Appendix</u>, <u>Figs. S7</u>** and **<u>S8</u>**). This might indicate race-dependent age-education interaction associations with brain volume or insufficiency of sample size for the non-British ancestry subjects.

Regarding the main effects, we observed 77.3% (17 out of 22) associations for ROI volumes and 79.5% (35 out of 44) for WM diffusivity measures that showed consistent directions when compared to non-British populations. They included brain temporal, cingulate, and ventricular regions, brainstem, cerebellum, and 15 WM tracts (**Figs 5A** and **5B)**. Most associations followed the patterns of increased cortical, ventricular, subcortical and cerebellar volumes, increased FA, AD, and MO, and decreased MD and RD with higher education levels.

Furthermore, our analysis revealed that directions of associations between the education PRS and brain structure were consistent with the education phenotype. These associations included increased volume of the amygdala, fusiform, and fourth ventricle, as well as increased FA, AD, RD and MD (**Figs. 5A** and **5B**). This suggests that the corresponding brain structural changes affected by education may be partially genetically driven.

## Discussion

In this study, we performed a comprehensive multi-factorial association analysis of brain structure with various risk factors using data from UKB subjects. Our findings underscore the impact of complex interactions between age, sex, genetics, and environmental exposures such as smoking, alcohol consumption, and education on brain structure.

### A Fine-grained Underpinning of APOE Effect on Aging Brains

We provided detailed association analyses of genetics, sex, and age with brain structure. As far as we know, our research is the first to systematically investigate age-sex-*APOE*-ε2/ -ε4 three way interactions on brain structure while accounting for many confounders. Previous research on interactions of *APOE*-ε2 are rare, limited by small sample sizes. Besides, the effects of *APOE*-ε2 are multifaceted and require careful examination. While *APOE*-ε2 has been demonstrated to have a protective effect against Alzheimer’s disease, it has also been associated with an increased risk of various other neurological disorders, such as post-traumatic stress disorder and argyrophilic grain disease <u>(85–89)</u>.

In our study, we found that *APOE*-*ε*4 accelerated the reduction of amygdala volume, whereas *APOE*-*ε*2 mitigated this decrease. These effects were more pronounced in females. In contrast, the brain structural changes in the general population (mostly *APOE* non-carriers) were more evident in males. These sex-disparate differences between *APOE* carriers and non-carriers could potentially be attributed to the greater hypometabolism and atrophy among female *APOE*-ε4 carriers (90) and protective influence of female sex hormones among *APOE*-ε4 non-carriers (16, 17, 91–93).

Our study examined the *APOE*-alcohol interaction, contributing to the understanding of the complex relationship between alcohol consumption and brain health. Although some research suggests heavy alcohol consumption may be associated with a decline in brain volume (59), other studies indicate that moderate alcohol consumption might have a protective effect (63). Findings on moderate alcohol consumption have been inconsistent, and the underlying mechanisms remain unclear (63, 64, 66, 94). By using a larger sample size our study found associations between moderate alcohol consumption and diffusivity measures in the superior fronto-occipital fasciculus. This result aligned with a previous study (66), which demonstrated that moderate alcohol consumption could be harmful to learning and memory among *APOE*-ε4 carriers, but not in non-carriers. Additional investigation is warranted to better understand the underlying mechanisms for this association.

### Environmental Factors are extensively Associated with Brain Structure Changes

Smoking and excessive alcohol consumption, both well-established cardiovascular and brain structural risk factors, negatively affect the cardiovascular system and harm brain health (58, 95). Conversely, healthy diet, education and physical activity are three major factors extensively studied for their contributions to building brain reserve throughout life (67, 96). The effects of these lifestyle factors vary across the brain. Specifically, in our analyses, smoking primarily affected subcortical and ventricular regions, alcohol affected frontal, parietal, subcortical, and ventricular regions, and education influenced the temporal, inferior frontal, subcortical and ventricular regions (**Figs. 3-5**).

Many of our findings are supported by previous literature. Smoking has been linked to reduction of subcortical volume (76, 97) and dysfunction of limbic network (98). Also based on the UKB data, (99) checked 166 regional brain GM volumes using 33,293 subjects and found that the small volume was most evident in thalamus. For WM microstructure, our findings were consistent with another study (100) based on 17,760 UKB subjects, suggesting reduced FA and increased MD with smoking. For alcohol, a recent study (101) identified that alcohol was even more associated with reduced GM volume than smoking, BMI, blood pressure and cholesterol considered based on 25,378 UKB subjects. Specifically, our findings of alcohol were also validated on the volume of ventricle, frontal, parietal, accumbens and thalamus (24, 101–103), and CC, anterior corona radiata (ACR), posterior corona radiata (PCR), CGC, FX, ALIC, posterior limb of internal capsule (PLIC) and SLF (65, 104, 105). In addition, a previous study on physical activity discovered an association with the increase of GM volume among older adults (> 60 years old) (22) based on the 5272 participants from the UKB.

In our findings, the identified effects of excessive alcohol and smoking shared many similarities, including lower cortical and subcortical volumes, larger ventricular volume reduced WM integrity (as evidenced by smaller FA and enlarged AxD, RD, and MD). Actually a previous large-scale study (48) involving 9,772 participants from the UKB has identified associations between vascular risk factors and smaller frontal, temporal cortical and subcortical volumes, reduced integrity of WM and thalamic pathways.

However, compared to smoking, excessive alcohol consumption appears to have more widespread detrimental effects on the brain, affecting both cortical and subcortical regions, and exhibiting stronger interaction effects with age. These differences may be attributed to the distinct mechanisms of brain damage caused by alcohol and smoking. Alcohol has direct neurotoxic effects on brain cells (106) and neurotransmitter systems (107), leading to neuronal death and degeneration, while smoking exerts its effects on the brain indirectly through oxidative stress, vascular changes, and inflammation (108). Additionally, older adults are more likely to be taking medications, and alcohol interacts with a wider range of medications, potentially causing further brain damage (109). Therefore the age interactions may differ between smoking and excessive alcohol consumption.

While there are studies focusing on the effects of diets on overall brain health or cognitive decline (23, 67–70), research on specific brain regions during brain aging remain scarce, and rare existing literature addresses the age-related differences. A large study (110) looked into the association between obesity and brain volume and WM microstructure based on 12,087 participants from the UKB, but did not study the diet structure. A recent large-scale study specifically worked on the Mediterranean diet and the hippocampal and WM hyperintensity volumes based on 21,933 participants (111). Another study (112) pointed out that the frontal, parietal and temporal volumes may mediate the association between diet composition with behavioral disinhibition. We here discovered that diet may lead to ventricular and visual regional changes among younger adults affecting older adults more significantly among paracentral and and inferior frontal regions. As the definition of a healthy diet was based on categories of fruits, vegetables, whole grains, refined grains, fish, unprocessed meat, and processed meat, further research is necessary to identify specific food categories that lead to the age-interaction patterns.

Although several studies explored the importance of healthy sleep for different age groups (113– 115), they do not directly compare sleep’s impact on specific age groups. Our results, showing negative age-sleep interactions on the right superior frontal volume suggest that healthy sleep may have greater benefits for younger adults in superior frontal regions. This might be due to age-related declines in neural plasticity and differing sleep patterns in old adults, such as fragmented sleep, reduced slow-wave sleep, and earlier sleep timing. Aging is associated with natural structural and functional changes in the brain, which could make it less responsive to the positive effects of sleep. It is important to note that these explanations are speculative, and further research is needed to confirm these findings and better understand the underlying mechanisms behind this age-related difference.

Higher education level is considered a protective factor that benefits subcortical volumes and regional WM integrity. Previous research has shown that individuals with higher levels of education exhibit improved FA in fiber tracts of the parietal, temporal, and occipital lobes, including fusiform and parahippocampal gyrus (116). Additionally, higher education has been associated with larger volumes of frontal and temporal lobes (117), larger CSF volume (118, 119), and improved functional connectivities between the anterior cingulate cortex and frontal, temporal and parietal lobes (120).

However, the relationship between higher education and AxD is not well understood. Our observations suggest that higher education may be related to an increased AxD, which is different from the absence of smoking or excessive alcohol consumption. It is believed that higher education can increase brain plasticity and stimulate the formation of new connections between neurons, leading to an increase in AxD. This increase in AxD may indicate larger axons, which could result in more efficient signal transmission. In contrast, increases in AxD, RD and MD due to smoking and excessive alcohol consumption suggest a less organized and less tightly packed WM network, as water molecules can move more freely. Further research is needed to fully understand differences of the relationship between AxD and education, smoking and alcohol on the WM network.

### Limitations and Future Work

Our study differs from previous similar research in two ways. Firstly, we investigated a wide range of modifiable and non-modifiable risk factors and explored their joint, conditional and interaction effects on brain structure. Secondly, we validated our findings from the white British population by using an independent non-British ancestry population as a test set. This ensured the robustness of our results against potential racial differences, outlier sensitivities and sample size limitations.

There are several limitations in our current work that should be acknowledged. First, we observed some inconsistent effects across the white British and non-British ancestry populations. This may be due to several reasons, including differences in race, sample size, and outliers. It is unrealistic to further subdivide the non-British ancestry subjects into subgroups (Asian, African, or Hispanic) due to the limited sample size. Second, there may be measurement errors in some risk factors, as all questionnaire variables were measured only once. Third, since the UKB is a volunteer-based study, the participants may be healthier than the general population, which may lead to inconsistent conclusions when compared to previous community-based studies. Lastly, further analyses such as mediation and causal analyses would be needed to fully understand the underlying mechanisms of the direct and indirect main and interaction effects of risk factors considered in this study. Studies on causal pathways of nicotine dependence (7) using 23,624 subjects from UKB revealed that 272 single-nucleotide polymorphisms (SNP) effects on smoking status were mediated by the FA, particularly at the right ALIC. In addition, another literature (6) recognized the causal relationship between heavy alcohol use, smoking and the reduced subcortical volume. As our future work, we will perform similar causal pathway analyses to further elucidate the relationships between risk factors and brain structure.

## Methods

### Study Population

The UKB released a dataset of 500,000 participants, of which more than 40,000 had MRI data. Through data processing, we obtained DTI phenotype data of 37,123 subjects from UKB phase 1, 2 and 3, comprising both genetic and lifestyle information. Our analyses of WM were restricted to 35,035 unrelated individuals with DTI phenotypes, genetic and lifestyle information available. Our primary discoveries were based on the 32,320 white British UKB subjects, which we validated on the remaining 2,715 non-British ancestry subjects. For the study of brain volume, we obtained volumetric phenotype data for 39,216 UKB samples and restricted the brain volumetric analysis to 36,969 independent individuals with volumes for ROIs, genetic information and lifestyle information available. Our primary discoveries were based on 34,097 white British subjects, which we validated on the remaining 2,872 non-British ancestry subjects.

### Imaging Data Processing

The T1 MRI and diffusion-weighted MRI datasets were processed using the procedures described in (19) and (121), respectively. Further details on image acquisition, preprocessing, and phenotype generation can be found in ***<u>SI Appendix</u>***. We generated 101 brain global and regional volumetric phenotypes of 39,216 UKB individuals, including the WBV, WM, GM, and CSF, and 110 DTI phenotypes of 37,123 UKB individuals, including the FA, MD, AxD, RD, and mode of diffusivity (MO) for 21 WM tracts. Subjects with any values greater than five times the median absolute deviation from the median value were considered as outliers and removed from the regression analyses.

### APOE Genotyping

The ε4 allele of the *APOE* gene is a well-established risk factor of AD. Two variants of SNPs, the rs429358 and rs7412, are used to determine the *APOE* genotype, resulting in 3 different alleles (*APOE*-ε2, *APOE*-ε3 and *APOE*-ε4) and six genotypes (*APOE*-ε3/ *APOE*-ε3, *APOE*-ε3/ *APOE*- ε4, *APOE*-ε2/ *APOE*-ε3, *APOE*-ε4/ *APOE*-ε4, *APOE*-ε2/ *APOE*-ε4 and *APOE*-ε2/ *APOE*-ε2) (122). Unlike the *APOE*-ε4, the *APOE*-ε2 has been found to reduce the risk of AD risk and is the least common allele in the population, while the *APOE*-ε3 is the most common and considered to be risk-neutral (123). The *APOE* genotyping information was extracted from the imputed SNP data from the UKB resources. Among the 35,035 unrelated subjects used in the main discovery and validation of 110 DTI phenotypes, *APOE* genotypes are distributed as follows: 770 subjects (2.2%) are *APOE*-ε4 homozygous, with 420 male and 350 female; 202 subjects (0.58%) are *APOE*-ε2 homozygous, with 93 male and 109 female; 20770 subjects (59.3%) are APOE-ε3 homozygous, with 9,929 male and 10,841 female; 4330 subjects (12.4%) are *APOE*-ε2/ *APOE*- ε3, with 2,032 male and 2,298 female; 833 subjects (2.4%) are *APOE*-ε2/ *APOE*-ε4, with 374 male and 459 female; and 8,130 subjects (23.2%) are *APOE*-ε3/ *APOE*-ε4, with 3,737 male and 4,393 female. The allele frequencies of ε2, ε3 and ε4 are 7.9%, 77.1% and 15.0%, respectively. **Table 1** provides demographic information for each genotype.

In our analyses, we included the number of *APOE*-ε4 allele and *APOE*-ε2 allele as two separate covariates in our models, to account for the additive effects of *APOE*-ε4 and *APOE*-ε2 alleles, respectively. This approach differs from some previous studies which categorized *APOE*-ε4 into two categories (*APOE*-ε4 homozygotes and all others), or considered *APOE*-ε2 homozygote and *APOE*-ε2/ *APOE*-ε3 as *APOE*-ε2 carriers and the remaining 4 genotypes as controls. The results of models using such dichotomous categorization of *APOE*-ε4 and *APOE*-ε2 can be found in the ***<u>SI Appendix</u>***.

### Polygenic Risk Score

A wide range of heritable brain diseases and behavioral traits have been linked to variations in brain structure (33, 124, 125). In this study, we will focus on 10 diseases from the UNC PGC (126), which include AD, ADHD, anxiety disorder, ASD, bipolar disorder, MDD, OCD, PD, SCH and TS, as well as 6 behavioral factors, such as alcohol dependence, cognitive performance, education, opioid dependence, self-employment and smoking. Each of these disorders and behavioral traits has been associated with common genetic variants through large-scale genome-wide association studies (GWAS) meta-analysis (127–141), which can be combined to generate PRSs. A PRS represents an individual’s genetic susceptibility to a particular condition or behavior. We incorporated these PRSs in our models to investigate how genetic variations affect brain structure. Additional information regarding the generation and characterization of PRSs for each of the aforementioned clinical factors can be found in the ***<u>SI Appendix</u>***.

### Lifestyle and SES factors

Most of the lifestyle variables in our analysis were drawn following the same criterion as in the healthy lifestyle and dementia study (142). Not being a current smoker, engaging in regular physical activity, and maintaining a healthy diet are well-established healthy lifestyle factors associated with a healthy brain. We categorized alcohol assumption into abstinence, moderate consumption and excessive consumption due to contradicting evidence on moderate alcohol consumption (24, 143). These four healthy lifestyle variables were collected through touchscreen questionnaires during the UKB baseline assessment. Smoking status was dichotomized into current-smokers and non-smokers. Following the American Heart Association (AHA) guidelines, a minimum of 150 minutes or 5 days of moderate exercise per week, or a minimum of 75 minutes or at least 1 day of vigorous activity per week is considered regular physical activity. For alcohol, the maximum threshold of moderate alcohol consumption is 14g/d for women and 28g/d for men, and the abstinence includes people not currently drinking or drinking only on special occasions with an overall alcohol intake of 0 units per day. A healthy diet is defined as meeting the adequate intake of at least 4 out of the 7 dietary recommendations for cardiometabolic health (vegetables, fruits, fish, unprocessed meat, processed meat, whole grains and refined grains) (144). Additionally, short and long sleep duration (sleeping less than 6 hours or longer than 8 hours) is also a risk factor for developing dementia (145). Besides the above lifestyle factors, we also included education level and social deprivation factors in our analysis. Education level was divided into with or without a college or university degree or above. Social deprivation (SoDep) was measured by the Townsend deprivation index which corresponds to the postcode of each participant’s location. It was dichotomized into two groups, above the Townsend deprivation index median and below the median. Details with respect to how we produced these lifestyle and SES factors are provided in ***<u>SI Appendix</u>*, <u>Supplementary Text</u>** and **<u>Table S1</u>**.

### Covariates

In all the models, we adjusted for the participants’ partnership status (categorized into whether participants live with their partner), study phase (categorized into phase 1, phase 2 and phase3), age at imaging, sex, and sex and age interaction. We adjusted for the WBV as a confounding covariate for analyses of all the other brain volumetric phenotypes.

### Statistical Analysis

Missing value imputations for covariates and imaging traits were included in the ***<u>SI Appendix</u>***. We carried out four levels of multivariate linear regression models on the white British UKB subjects to answer the following questions: (1) Is there the *APOE* genotype and age interaction effect on DTI phenotypes and ROI volumes? (2) Does the *APOE*-age interaction effect vary between males and females? (3) Are there associations between DTI phenotypes or ROI volumes and healthy lifestyle factors, SES, and PRSs, and age interactions (4) Is there *APOE* genotype and environmental factor interaction effects on DTI phenotypes and ROI volumes?

To answer questions (1), we used the standardized 110 DTI measurements and 101 ROI volumes as the dependent variable and *APOE*-ε2, *APOE*-ε4 count, sex, age, PRSs, lifestyle factors, UKB study phase, marriage status, sex and age as the predictor variables. We implemented Bonferroni correction for the p-values to adjust for multiple testing, with the threshold of significance for 211 independent testing set at approximately 2.37E-4 (0.05/211). To investigate the effect of *APOE* genotype and age interaction on DTI phenotypes and ROI volumes, we added these two-way interaction terms to our models. For identifying the difference of *APOE* genotype and age interaction effect on DTI phenotypes and ROI volumes in different sex groups in question (2), the 3-way interaction term of age, sex and *APOE*, and three 2-way interaction terms between age, sex and *APOE* were included in the regression models, along with the full set of covariates. To address question (3), two way interaction terms between age and healthy lifestyle factors and SES were added separately. To address question (4), we included *APOE*-environmental interactions and the age-environmental interaction, separately. Detailed statistical models we considered for the association between main covariates, two-way and three-way interactions and brain structure were displayed in ***<u>SI Appendix</u>*, <u>Supplementary text</u>**.

We verified our discovery on an independent set of non-British ancestry UKB subjects. The same association analyses were conducted in the non-British UKB populations using a confidence level of 0.05 as the cutoff. Due to the relatively small sample size of the validation set, we considered associations to be significant if they passed the Bonferroni corrected threshold of *2*.*37E-4* in our main discovery. The validation results serve to validate and demonstrate the robustness of findings. Throughout the paper, unless otherwise specified, we only report findings if they meet the following criteria: (1) they are significant among the white British population; (2) They can be validated (showing the same direction of effects) in the non-British ancestry population. Additionally, we discuss interaction effects with *p < 0*.*01* among the white British population that can be validated by the non-British ancestry population. However, these effects will not be counted as significant findings unless their p-values pass the threshold of *2*.*37E-4*. To interpret the interaction effect related with sex, the analyses were carried out separately for male and female groups (***<u>SI Appendix</u>*, <u>Supplementary text</u>**).

## Data Availability

All data produced in the present study are available upon reasonable request to the authors
All data produced in the present work are contained in the manuscript
All data produced are available online at

https://biobank.ndph.ox.ac.uk/

## Acknowledgement

This research was supported in part by U.S. NIH grants NS110791 (J.C. and G.W.) and MH116527 (H.Z. and T.L.). We thank the individuals represented in the UK Biobank, studies for their participation and the research teams for their work in collecting, processing and disseminating these datasets for analysis. We gratefully acknowledge all the studies and databases that made GWAS summary data available. This research has been conducted using the UK Biobank resource (application number 22783), subject to a data transfer agreement.

## References

1. F. B. Johnson, D. A. Sinclair, L. Guarente, Molecular Biology of Aging. Cell 96, 291–302 (1999).

2. L. Ferrucci, et al., Measuring biological aging in humans: A quest. Aging Cell 19, e13080 (2020).

3. C. E. Finch, D. M. Cohen, Aging, Metabolism, and Alzheimer Disease: Review and Hypotheses. Exp. Neurol. 143, 82–102 (1997).

4. T. A. D. Jolly, et al., Microstructural white matter changes mediate age-related cognitive decline on the Montreal Cognitive Assessment (MoCA). Psychophysiology 53, 258–267 (2016).

5. F. Gomez-Pinilla, S. Vaynman, Z. Ying, Brain-derived neurotrophic factor functions as a metabotrophin to mediate the effects of exercise on cognition. Eur. J. Neurosci. 28, 2278– 2287 (2008).

6. E. Logtenberg, et al., Investigating the causal nature of the relationship of subcortical brain volume with smoking and alcohol use. Br. J. Psychiatry J. Ment. Sci., 1–9 (2021).

7. Z. Ye, et al., White Matter Integrity and Nicotine Dependence: Evaluating Vertical and Horizontal Pleiotropy. Front. Neurosci. 15, 738037 (2021).

8. E. Fletcher, et al., Brain volume change and cognitive trajectories in aging. Neuropsychology 32, 436–449 (2018).

9. M. W. Vernooij, et al., White matter microstructural integrity and cognitive function in a general elderly population. Arch. Gen. Psychiatry 66, 545–553 (2009).

10. A. M. Fjell, K. B. Walhovd, Structural brain changes in aging: courses, causes and cognitive consequences. Rev. Neurosci. 21, 187–221 (2010).

11. G. Hueluer, H. Dodge, New developments in cognitive aging research. Innov. Aging 2, 382 (2018).

12. F. Tang, Successful Aging: Multiple Trajectories and Population Heterogeneity. Int. J. Soc. Sci. Stud. 2, 12–22 (2014).

13. N. Raz, P. Ghisletta, K. M. Rodrigue, K. M. Kennedy, U. Lindenberger, Trajectories of brain aging in middle-aged and older adults: Regional and individual differences. NeuroImage 51, 501–511 (2010).

14. R. a. I. Bethlehem, et al., Brain charts for the human lifespan. Nature 604, 525–533 (2022).

15. M. Ingalhalikar, et al., Sex differences in the structural connectome of the human brain. Proc. Natl. Acad. Sci. 111, 823–828 (2014).

16. D. G. Murphy, et al., Sex differences in human brain morphometry and metabolism: an in vivo quantitative magnetic resonance imaging and positron emission tomography study on the effect of aging. Arch. Gen. Psychiatry 53, 585–594 (1996).

17. P. E. Cowell, et al., Sex differences in aging of the human frontal and temporal lobes. J. Neurosci. 14, 4748–4755 (1994).

18. L. T. Elliott, et al., Genome-wide association studies of brain imaging phenotypes in UK Biobank. Nature 562, 210–216 (2018).

19. B. Zhao, et al., Genome-wide association analysis of 19,629 individuals identifies variants influencing regional brain volumes and refines their genetic co-architecture with cognitive and mental health traits. Nat. Genet. 51, 1637–1644 (2019).

20. P. M. Thompson, et al., Genetic influences on brain structure. Nat. Neurosci. 4, 1253– 1258 (2001).

21. J. Gallinat, et al., Smoking and structural brain deficits: a volumetric MR investigation. Eur. J. Neurosci. 24, 1744–1750 (2006).

22. M. Hamer, N. Sharma, G. D. Batty, Association of objectively measured physical activity with brain structure: UK Biobank study. J. Intern. Med. 284, 439–443 (2018).

23. D. E. A. Jensen, V. Leoni, M. C. Klein-Flügge, K. P. Ebmeier, S. Suri, Associations of dietary markers with brain volume and connectivity: A systematic review of MRI studies. Ageing Res. Rev. 70, 101360 (2021).

24. R. Daviet, et al., Associations between alcohol consumption and gray and white matter volumes in the UK Biobank. Nat. Commun. 13, 1175 (2022).

25. L. Nyberg, et al., Educational attainment does not influence brain aging. Proc. Natl. Acad. Sci. 118, e2101644118 (2021).

26. H. Kweon, et al., Human brain anatomy reflects separable genetic and environmental components of socioeconomic status. Sci. Adv. 8, eabm2923 (2022).

27. A. Altmann, L. Tian, V. W. Henderson, M. D. Greicius, Alzheimer’s Disease Neuroimaging Initiative Investigators, Sex modifies the APOE-related risk of developing Alzheimer disease. Ann. Neurol. 75, 563–573 (2014).

28. C. E. Coffey, et al., Sex differences in brain aging: a quantitative magnetic resonance imaging study. Arch. Neurol. 55, 169–179 (1998).

29. M. Régy, et al., Association of APOE ε4 with cerebral gray matter volumes in non-demented older adults: The MEMENTO cohort study. NeuroImage 250, 118966 (2022).

30. D. M. Lyall, et al., Is there association between APOE e4 genotype and structural brain ageing phenotypes, and does that association increase in older age in UK Biobank? (N = 8,395). 230524 (2017).

31. G. Operto, et al., Interactive effect of age and APOE-ε4 allele load on white matter myelin content in cognitively normal middle-aged subjects. NeuroImage Clin. 24, 101983 (2019).

32. C.-L. Kuo, L. C. Pilling, J. L. Atkins, G. A. Kuchel, D. Melzer, ApoE e2 and aging-related outcomes in 379,000 UK Biobank participants. Aging 12, 12222–12233 (2020).

33. S. Ranlund, et al., Associations between polygenic risk scores for four psychiatric illnesses and brain structure using multivariate pattern recognition. NeuroImage Clin. 20, 1026–1036 (2018).

34. C. van der Merwe, et al., Polygenic risk for schizophrenia and associated brain structural changes: A systematic review. Compr. Psychiatry 88, 77–82 (2019).

35. E.-M. Stauffer, et al., Grey and white matter microstructure is associated with polygenic risk for schizophrenia. Mol. Psychiatry 26, 7709–7718 (2021).

36. S. Grama, et al., Polygenic risk for schizophrenia and subcortical brain anatomy in the UK Biobank cohort. Transl. Psychiatry 10, 309 (2020).

37. H. L. Chandler, C. J. Hodgetts, X. Caseras, K. Murphy, T. M. Lancaster, Polygenic risk for Alzheimer’s disease shapes hippocampal scene-selectivity. Neuropsychopharmacology 45, 1171–1178 (2020).

38. M. N. Braskie, et al., Common Alzheimer’s Disease Risk Variant Within the CLU Gene Affects White Matter Microstructure in Young Adults. J. Neurosci. 31, 6764–6770 (2011).

39. A. L. Krain, F. X. Castellanos, Brain development and ADHD. Clin. Psychol. Rev. 26, 433–444 (2006).

40. W. Wang, et al., Reduced white matter integrity and its correlation with clinical symptom in first-episode, treatment-naive generalized anxiety disorder. Behav. Brain Res. 314, 159–164 (2016).

41. D. K. Shukla, B. Keehn, R.-A. Müller, Tract-specific analyses of diffusion tensor imaging show widespread white matter compromise in autism spectrum disorder. J. Child Psychol. Psychiatry 52, 286–295 (2011).

42. S. M. Strakowski, et al., Brain Magnetic Resonance Imaging of Structural Abnormalities in Bipolar Disorder. Arch. Gen. Psychiatry 56, 254 (1999).

43. N. Vasic, et al., Baseline brain perfusion and brain structure in patients with major depression: a multimodal magnetic resonance imaging study. J. Psychiatry Neurosci. 40, 412–421 (2015).

44. J. Pujol, et al., Mapping Structural Brain Alterations in Obsessive-Compulsive Disorder. Arch. Gen. Psychiatry 61, 720 (2004).

45. D. H. Han, et al., Altered cingulate white matter connectivity in panic disorder patients. J. Psychiatr. Res. 42, 399–407 (2008).

46. K. L. Davis, et al., White Matter Changes in Schizophrenia: Evidence for Myelin-Related Dysfunction. Arch. Gen. Psychiatry 60, 443 (2003).

47. K. A. Fredericksen, et al., Disproportionate increases of white matter in right frontal lobe in Tourette syndrome. Neurology 58, 85–89 (2002).

48. S. R. Cox, et al., Associations between vascular risk factors and brain MRI indices in UK Biobank. Eur. Heart J. 40, 2290–2300 (2019).

49. R. Doll, R. Peto, J. Boreham, I. Sutherland, Mortality in relation to smoking: 50 years’ observations on male British doctors. BMJ 328, 1519 (2004).

50. J. A. Ambrose, R. S. Barua, The pathophysiology of cigarette smoking and cardiovascular disease: An update. J. Am. Coll. Cardiol. 43, 1731–1737 (2004).

51. G. Zhong, Y. Wang, Y. Zhang, J. J. Guo, Y. Zhao, Smoking Is Associated with an Increased Risk of Dementia: A Meta-Analysis of Prospective Cohort Studies with Investigation of Potential Effect Modifiers. PLOS ONE 10, e0118333 (2015).

52. H. van Ewijk, et al., Smoking and the developing brain: Altered white matter microstructure in attention-deficit/hyperactivity disorder and healthy controls: Smoking and White Matter Microstructure. Hum. Brain Mapp. 36, 1180–1189 (2015).

53. J. H. Cole, Multimodality neuroimaging brain-age in UK biobank: relationship to biomedical, lifestyle, and cognitive factors. Neurobiol. Aging 92, 34–42 (2020).

54. K. Ning, L. Zhao, W. Matloff, F. Sun, A. W. Toga, Association of relative brain age with tobacco smoking, alcohol consumption, and genetic variants. Sci. Rep. 10, 10 (2020).

55. S. M. Smith, D. Vidaurre, F. Alfaro-Almagro, T. E. Nichols, K. L. Miller, Estimation of brain age delta from brain imaging. NeuroImage 200, 528–539 (2019).

56. M. Habes, et al., Advanced brain aging: relationship with epidemiologic and genetic risk factors, and overlap with Alzheimer disease atrophy patterns. Transl. Psychiatry 6, e775 (2016).

57. P. E. Ronksley, S. E. Brien, B. J. Turner, K. J. Mukamal, W. A. Ghali, Association of alcohol consumption with selected cardiovascular disease outcomes: a systematic review and meta-analysis. The BMJ 342, d671 (2011).

58. M. R. Piano, Alcohol’s Effects on the Cardiovascular System. Alcohol Res. Curr. Rev. 38, 219–241 (2017).

59. A. Pfefferbaum, et al., Brain Gray and White Matter Volume Loss Accelerates with Aging in Chronic Alcoholics: A Quantitative MRI Study. Alcohol. Clin. Exp. Res. 16, 1078–1089 (1992).

60. ‘Keefe James H. OK. A. Bybee, C. J. Lavie, Alcohol and Cardiovascular Health. J. Am. Coll. Cardiol. 50, 1009–1014 (2007).

61. E. B. Rimm, P. Williams, K. Fosher, M. Criqui, M. J. Stampfer, Moderate alcohol intake and lower risk of coronary heart disease: meta-analysis of effects on lipids and haemostatic factors. BMJ 319, 1523–1528 (1999).

62. S. E. Brien, P. E. Ronksley, B. J. Turner, K. J. Mukamal, W. A. Ghali, Effect of alcohol consumption on biological markers associated with risk of coronary heart disease: systematic review and meta-analysis of interventional studies. BMJ 342, d636 (2011).

63. Y. Gu, et al., Alcohol intake and brain structure in a multiethnic elderly cohort. Clin. Nutr. Edinb. Scotl. 33, 662–667 (2014).

64. E. A. de Bruin, et al., Associations Between Alcohol Intake and Brain Volumes in Male and Female Moderate Drinkers. Alcohol Clin. Exp. Res. 29, 656–663 (2005).

65. A. Topiwala, et al., Moderate alcohol consumption as risk factor for adverse brain outcomes and cognitive decline: longitudinal cohort study. BMJ 357, j2353 (2017).

66. B. Downer, F. Zanjani, D. W. Fardo, The Relationship Between Midlife and Late Life Alcohol Consumption, APOE e4 and the Decline in Learning and Memory Among Older Adults. Alcohol Alcohol. Oxf. Oxfs. 49, 17–22 (2014).

67. Y. Gu, et al., Mediterranean diet and brain structure in a multiethnic elderly cohort. Neurology 85, 1744–1751 (2015).

68. M. Luciano, et al., Mediterranean-type diet and brain structural change from 73 to 76 years in a Scottish cohort. Neurology 88, 449–455 (2017).

69. H. Macpherson, S. A. McNaughton, K. E. Lamb, C. M. Milte, Associations of Diet Quality with Midlife Brain Volume: Findings from the UK Biobank Cohort Study. J. Alzheimers Dis. JAD 84, 79–90 (2021).

70. P. H. Croll, et al., Better diet quality relates to larger brain tissue volumes: The Rotterdam Study. Neurology 90, e2166–e2173 (2018).

71. F. Novellino, et al., Association Between Hippocampus, Thalamus, and Caudate in Mild Cognitive Impairment APOEε4 Carriers: A Structural Covariance MRI Study. Front. Neurol. 10 (2019).

72. G. Operto, et al., White matter microstructure is altered in cognitively normal middle-aged APOE-ε4 homozygotes. Alzheimers Res. Ther. 10, 48 (2018).

73. R. Zivadinov, et al., Smoking is associated with increased lesion volumes and brain atrophy in multiple sclerosis. Neurology 73, 504–510 (2009).

74. M. Vňuková, R. Ptáček, J. Raboch, G. B. Stefano, Decreased Central Nervous System Grey Matter Volume (GMV) in Smokers Affects Cognitive Abilities: A Systematic Review. Med. Sci. Monit. Int. Med. J. Exp. Clin. Res. 23, 1907–1915 (2017).

75. M. T. Sutherland, et al., Chronic cigarette smoking is linked with structural alterations in brain regions showing acute nicotinic drug-induced functional modulations. Behav. Brain Funct. 12, 16 (2016).

76. D. Das, N. Cherbuin, K. J. Anstey, P. S. Sachdev, S. Easteal, Lifetime cigarette smoking is associated with striatal volume measures. Addict. Biol. 17, 817–825 (2012).

77. C. E. Sexton, et al., Accelerated Changes in White Matter Microstructure during Aging: A Longitudinal Diffusion Tensor Imaging Study. J. Neurosci. 34, 15425–15436 (2014).

78. F. Lin, G. Wu, L. Zhu, H. Lei, Heavy smokers show abnormal microstructural integrity in the anterior corpus callosum: a diffusion tensor imaging study with tract-based spatial statistics. Drug Alcohol Depend. 129, 82–87 (2013).

79. W. Umene-Nakano, et al., Abnormal White Matter Integrity in the Corpus Callosum among Smokers: Tract-Based Spatial Statistics. PLoS ONE 9, e87890 (2014).

80. T. D. Kristensen, et al., Changes in negative symptoms are linked to white matter changes in superior longitudinal fasciculus in individuals at ultra-high risk for psychosis. Schizophr. Res. 237, 192–201 (2021).

81. R. R. Savjani, et al., Characterizing white matter changes in cigarette smokers via diffusion tensor imaging. Drug Alcohol Depend. 145, 134–142 (2014).

82. A. S. Whiteman, D. E. Young, A. E. Budson, C. E. Stern, K. Schon, Entorhinal volume, aerobic fitness, and recognition memory in healthy young adults: A voxel-based morphometry study. NeuroImage 126, 229–238 (2016).

83. M. Yamamoto, et al., Association between exercise habits and subcortical gray matter volumes in healthy elderly people: A population-based study in Japan. eNeurologicalSci 7, 1–6 (2017).

84. T. W. Scheewe, et al., Exercise therapy improves mental and physical health in schizophrenia: a randomised controlled trial. Acta Psychiatr. Scand. 127, 464–473 (2013).

85. T. Y. Kim, et al., Apolipoprotein E Gene Polymorphism, Alcohol Use, and Their Interactions in Combat-Related Posttraumatic Stress Disorder. Depress. Anxiety 30, 1194–1201 (2013).

86. G. J. McKay, et al., Evidence of association of APOE with age-related macular degeneration - a pooled analysis of 15 studies. Hum. Mutat. 32, 1407–1416 (2011).

87. Z. Li, F. Shue, N. Zhao, M. Shinohara, G. Bu, APOE2: protective mechanism and therapeutic implications for Alzheimer’s disease. Mol. Neurodegener. 15, 63 (2020).

88. N. Zhao, et al., APOE ε2 is associated with increased tau pathology in primary tauopathy. Nat. Commun. 9, 4388 (2018).

89. E. Ghebremedhin, et al., Argyrophilic grain disease is associated with apolipoprotein E ε2 allele. Acta Neuropathol. (Berl.) 96, 222–224 (1998).

90. F. Sampedro, et al., APOE-by-sex interactions on brain structure and metabolism in healthy elderly controls. Oncotarget 6, 26663–26674 (2015).

91. R. C. Gur, et al., Gender differences in age effect on brain atrophy measured by magnetic resonance imaging. Proc. Natl. Acad. Sci. 88, 2845–2849 (1991).

92. J. Golomb, et al., Hippocampal Atrophy in Normal Aging: An Association With Recent Memory Impairment. Arch. Neurol. 50, 967–973 (1993).

93. N. Raz, et al., Selective aging of the human cerebral cortex observed in vivo: differential vulnerability of the prefrontal gray matter. Cereb. Cortex 7, 268–282 (1997).

94. B. J. K. Davis, et al., The Alcohol Paradox: Light-to-Moderate Alcohol Consumption, Cognitive Function, and Brain Volume. J. Gerontol. Ser. A 69, 1528–1535 (2014).

95. B. Messner, D. Bernhard, Smoking and Cardiovascular Disease. Arterioscler. Thromb. Vasc. Biol. 34, 509–515 (2014).

96. L. Fratiglioni, H.-X. Wang, Brain reserve hypothesis in dementia. J. Alzheimers Dis. JAD 12, 11–22 (2007).

97. T. C. Durazzo, D. J. Meyerhoff, K. K. Yoder, D. E. Murray, Cigarette smoking is associated with amplified age-related volume loss in subcortical brain regions. Drug Alcohol Depend. 177, 228–236 (2017).

98. A. C. Janes, L. D. Nickerson, B. deB. Frederick, M. J. Kaufman, Prefrontal and limbic resting state brain network functional connectivity differs between nicotine-dependent smokers and non-smoking controls. Drug Alcohol Depend. 125, 252–259 (2012).

99. Z. Linli, et al., Smoking is associated with lower brain volume and cognitive differences: A large population analysis based on the UK Biobank. Prog. Neuropsychopharmacol. Biol. Psychiatry 123, 110698 (2023).

100. J. C. Gray, et al., Associations of cigarette smoking with gray and white matter in the UK Biobank. Neuropsychopharmacology 45, 1215–1222 (2020).

101. A. Topiwala, K. Ebmeier, T. Maullin-Sapey, T. Nichols, No safe level of alcohol consumption for brain health: observational cohort study of 25,378 UK Biobank participant (2021) https://doi.org/10.1101/2021.05.10.21256931 (June 9, 2022).

102. D. Tomasi, et al., Accelerated Aging of the Amygdala in Alcohol Use Disorders: Relevance to the Dark Side of Addiction. Cereb. Cortex 31, 3254–3265 (2021).

103. G. Fein, R. Shimotsu, R. Chu, J. Barakos, Parietal gray matter volume loss is related to spatial processing deficits in long-term abstinent alcoholic men. Alcohol. Clin. Exp. Res. 33, 1806–1814 (2009).

104. M. A. Monnig, et al., White matter integrity is associated with alcohol cue reactivity in heavy drinkers. Brain Behav. 4, 158–170 (2014).

105. P.-H. Yeh, K. Simpson, T. C. Durazzo, S. Gazdzinski, D. J. Meyerhoff, Tract-based spatial statistics (TBSS) of diffusion tensor imaging data in alcohol dependence: abnormalities of the motivational neurocircuitry. Psychiatry Res. 173, 22–30 (2009).

106. I. I. Kruman, G. I. Henderson, S. E. Bergeson, DNA damage and neurotoxicity of chronic alcohol abuse. Exp. Biol. Med. 237, 740–747 (2012).

107. G. Chastain, Alcohol, Neurotransmitter Systems, and Behavior. J. Gen. Psychol. 133, 329–335 (2006).

108. A. Csiszar, et al., Oxidative stress and accelerated vascular aging: implications for cigarette smoking. Front. Biosci. Landmark Ed. 14, 3128–3144 (2009).

109. A. A. Moore, E. J. Whiteman, K. T. Ward, Risks of Combined Alcohol-Medication Use in Older Adults. Am. J. Geriatr. Pharmacother. 5, 64–74 (2007).

110. I. A. Dekkers, P. R. Jansen, H. J. Lamb, Obesity, Brain Volume, and White Matter Microstructure at MRI: A Cross-sectional UK Biobank Study. Radiology 291, 763–771 (2019).

111. S. Gregory, et al., Mediterranean diet and structural neuroimaging biomarkers of Alzheimer’s and cerebrovascular disease: A systematic review. Exp. Gerontol. 172, 112065 (2023).

112. D. van Rooij, L. Schweren, H. Shi, C. A. Hartman, J. K. Buitelaar, Cortical and Subcortical Brain Volumes Partially Mediate the Association between Dietary Composition and Behavioral Disinhibition: A UK Biobank Study. Nutrients 13, 3542 (2021).

113. L. Tarokh, J. M. Saletin, M. A. Carskadon, Sleep in adolescence: Physiology, cognition and mental health. Neurosci. Biobehav. Rev. 70, 182–188 (2016).

114. M. K. Scullin, D. L. Bliwise Sleep cognition, and normal aging: integrating a half century of multidisciplinary research. Perspect. Psychol. Sci. J. Assoc. Psychol. Sci. 10, 97–137 (2015).

115. M. M. Ohayon, M. A. Carskadon, C. Guilleminault, M. V. Vitiello, Meta-Analysis of Quantitative Sleep Parameters From Childhood to Old Age in Healthy Individuals: Developing Normative Sleep Values Across the Human Lifespan. Sleep 27, 1255–1273 (2004).

116. S. J. Teipel, et al., White matter microstructure in relation to education in aging and Alzheimer’s disease. J. Alzheimers Dis. JAD 17, 571–583 (2009).

117. D. W. Kang, et al., Differential Impact of Education on Gray Matter Volume According to Sex in Cognitively Normal Older Adults: Whole Brain Surface-Based Morphometry. Front. Psychiatry 12, 644148 (2021).

118. C. E. Coffey, J. A. Saxton, G. Ratcliff, R. N. Bryan, J. F. Lucke, Relation of education to brain size in normal aging: implications for the reserve hypothesis. Neurology 53, 189– 196 (1999).

119. D. Kidron, et al., Quantitative MR volumetry in Alzheimer’s disease. Topographic markers and the effects of sex and education. Neurology 49, 1504–1512 (1997).

120. E. M. Arenaza-Urquijo, et al., Relationships between years of education and gray matter volume, metabolism and functional connectivity in healthy elders. NeuroImage 83, 450– 457 (2013).

121. B. Zhao, et al., Common genetic variation influencing human white matter microstructure. Science 372, eabf3736 (2021).

122. L. Zhong, et al., A rapid and cost-effective method for genotyping apolipoprotein E gene polymorphism. Mol. Neurodegener. 11, 2 (2016).

123. A. Ward, et al., Prevalence of apolipoprotein E4 genotype and homozygotes (APOE e4/4) among patients diagnosed with Alzheimer’s disease: a systematic review and metaanalysis. Neuroepidemiology 38, 1–17 (2012).

124. T. Kaufmann, et al., Common brain disorders are associated with heritable patterns of apparent aging of the brain. Nat. Neurosci. 22, 1617–1623 (2019).

125. N. Bittner, et al., Combining lifestyle risks to disentangle brain structure and functional connectivity differences in older adults. Nat. Commun. 10, 621 (2019).

126. Summary Statistics – PGC -UNC (October 10, 2022).

127. T. Otowa, et al., Meta-analysis of genome-wide association studies of anxiety disorders.Mol. Psychiatry 21, 1391–1399 (2016).

128. E. A. Stahl, et al., Genome-wide association study identifies 30 loci associated with bipolar disorder. Nat. Genet. 51, 793–803 (2019).

129. B. W. Kunkle, et al., Genetic meta-analysis of diagnosed Alzheimer’s disease identifies new risk loci and implicates Aβ, tau, immunity and lipid processing. Nat. Genet. 51, 414– 430 (2019).

130. D. Demontis, et al., Discovery of the first genome-wide significant risk loci for attention deficit/hyperactivity disorder. Nat. Genet. 51, 63–75 (2019).

131. J. Grove, et al., Identification of common genetic risk variants for autism spectrum disorder. Nat. Genet. 51, 431–444 (2019).

132. International Obsessive Compulsive Disorder Foundation Genetics Collaborative (IOCDF-GC) and OCD Collaborative Genetics Association Studies (OCGAS), Revealing the complex genetic architecture of obsessive-compulsive disorder using meta-analysis. Mol. Psychiatry 23, 1181–1188 (2018).

133. A. J. Forstner, et al., Genome-wide association study of panic disorder reveals genetic overlap with neuroticism and depression. Mol. Psychiatry 26, 4179–4190 (2021).

134. S. Ripke, et al., Biological insights from 108 schizophrenia-associated genetic loci. Nature 511, 421–427 (2014).

135. D. Yu, et al., Interrogating the Genetic Determinants of Tourette’s Syndrome and Other Tic Disorders Through Genome-Wide Association Studies. Am. J. Psychiatry 176, 217– 227 (2019).

136. R. K. Walters, et al., Transancestral GWAS of alcohol dependence reveals common genetic underpinnings with psychiatric disorders. Nat. Neurosci. 21, 1656–1669 (2018).

137. B. Benyamin, et al., Childhood intelligence is heritable, highly polygenic and associated with FNBP1L. Mol. Psychiatry 19, 253–258 (2014).

138. J. J. Lee, et al., Gene discovery and polygenic prediction from a genome-wide association study of educational attainment in 1.1 million individuals. Nat. Genet. 50, 1112–1121 (2018).

139. R. Polimanti, et al., Leveraging genome-wide data to investigate differences between opioid use vs. opioid dependence in 41,176 individuals from the Psychiatric Genomics Consortium. Mol. Psychiatry 25, 1673–1687 (2020).

140. M. J. H. M. van der Loos, et al., The Molecular Genetic Architecture of Self-Employment. PLoS ONE 8, e60542 (2013).

141. M. Liu, et al., Association studies of up to 1.2 million individuals yield new insights into the genetic etiology of tobacco and alcohol use. Nat. Genet. 51, 237–244 (2019).

142. I. Lourida, et al., Association of Lifestyle and Genetic Risk With Incidence of Dementia. JAMA 322, 430 (2019).

143. S. Sabia, et al., Alcohol consumption and risk of dementia: 23 year follow-up of Whitehall II cohort study. BMJ 362, k2927 (2018).

144. D. Mozaffarian, Dietary and Policy Priorities for Cardiovascular Disease, Diabetes, and Obesity. Circulation 133, 187–225 (2016).

145. E. Palpatzis, N. Bass, R. Jones, N. Mukadam, Longitudinal association of apolipoprotein E and sleep with incident dementia. Alzheimers Dement. J. Alzheimers Assoc. 18, 888– 898 (2022).

