## Supplementary files for "The Interaction Effects of Age, APOE and Common Environmental Risk Factors on Human brain Structure"

***Brain Structural Mapping of Multi-factorial Aging Mechanism Using 35,921 UK Biobank Subjects***

**Supplementary Text**

**PRS Score Generation**

We calculated the polygenic risk scores (PRSs) of common variants following the tutorial (1). The first step in deriving PRS is to obtain the GWAS summary statistics from the GWAS study results on these phenotypes. To avoid overfitting due to sample overlap, we only downloaded and used summary statistics from GWAS studies (2–16) that exclusively use subjects from non-UKB data. We carried out the standard quality control (QC) steps by removing SNPs with minor allele frequency smaller than 0.01 and low imputation quality (’info score’ <0.8) as they are more likely to generate unreliable results. The second step involves removing the variants from the *APOE* region. Next, we calculated the PRS at different significant levels by weighing each of the significant single-nucleotide polymorphisms (SNPs) based on their associations with the phenotypes in the GWAS studies. The PRSs we included in our models were generated from a significance level of 0.8.

**UKB Image Acquisition**

We obtained the T1 magnetic resonance imaging (MRI) and diffusion MRI (dMRI) data in NIFTI format from the UK Biobank (UKB) study, which received ethics approval from the North West Multicentre Research Ethics Committee (approval number: 11/NW/0382). The T1 MRI data were acquired as a 3D MPRAGE sequence with 1 × 1 mm in-plane resolution, 1 mm sagittal slices, in-plane acceleration iPAT = 2, and prescan normalization. The dMRI data were acquired at the 2 × 2 × 2 mm spatial resolution, with anterior-to-posterior phase - encoding, a multiband acceleration factor of three, two nonzero b-values of 1000 s/mm2 and 2000 s/mm2, and 50 diffusion encoding directions were adopted for each of the two b-values. The diffusion preparation was a standard (monopolar) Stejskal Tanner pulse sequence, with echo time (TE) of 92 ms and repetition time (TR) of 3600 ms. Detailed information can be found in the UKB primary brain imaging documentation (18).

**UKB Image Processing**

In total 211 imaging traits were generated, including 101 volumetric and 110 diffusivity phenotypes, based on T1 MRI and dMRI data, respectively. The T1 MRI data were preprocessed locally using the same method as described in (19) with Advanced Normalization Tools (ANTs) (20, 21) to generate brain regional volumes for each subject. Specifically, individual images were registered to the standard OASIS-30 Atropos template and the [OASIS-TRT-20 joint fusion atlas](https://osf.io/d2cmy/?action=download&version=1) (based on the Mindboggle-101 annotated data) was adopted for brain regions of interest (ROI) labeling. We generated 101 brain regional and overall volumetric phenotypes for 39,216 UKB individuals, including whole brain volume (WBV), and white matter (WM), gray matter, and cerebrospinal fluid volumes. Detailed information can be found at <https://mindboggle.info/data.html>.

The dMRI data underwent the same steps as described in (22) by incorporating the UKB preprocessing steps and the TBSS-ENIGMA pipelines. This process included eddy current correction, head motion correction, gradient distortion correction, outlier slice removal, diffusion tensor imaging (DTI) model fitting, standard linear and nonlinear registrations, QC steps, skeletonization and regional feature extraction. These steps enabled the fractional anisotropy (FA), mean diffusivity (MD), axial diffusivity (AD), radial diffusivity (RD), and mode of anisotropy (MO) feature maps to be registered to the MNI-ICBM-152 standard space and projected onto the TBSS skeleton in the common space. Regional averages of the skeletonized FA, MD, AD, RD, and MO images were taken within 21 predefined WM tracts, respectively, based on the JHU ICBM-DTI-81 WM atlas. After data processing, 110 DTI diffusivity parameters were obtained for 37,123 subjects from UKB phase 1, 2 and 3.

**Subject Exclusion and Data Merge**

There are 37,123 subjects from UKB phase 1, 2 and 3 with dMRI, genetic and lifestyle information available and 39,216 UKB samples with volumetric phenotypes were available after the processing steps of dMRI and T1 MRI data, respectively. We identified first-degree relatives with at least one relative in the dataset using UKB field 22021 derived from the method of KING v.2.0 (23). For each first-degree pair we randomly excluded one subject and restricted our white matter analysis to the final 35,921 and 37,939 independent individuals with DTI and volumetric phenotypes, respectively.

**Missing Data Imputation**

Some ROI volumes of the 37,939 individuals are missing. For the optic chiasm 11,340 out of the 37,939 subjects are missing (out of view). The other 100 ROI volumes have less than 1% missing rates of the data. All DTI phenotypes of the 35,921 samples have less than 3% missing rates of the data. When we ran multiple regression models, we did not impute the missing ROI volumes and DTI phenotypes, which were the outcome variables. The data with missing ROI volumes or DTI phenotypes were dropped automatically when we fitted the regression model. For covariates, all variables (age, sex, APOE, PRSs, lifestyle, SES) have less than 1% missing rate of the data except the marriage status and the education level. We imputed these missing values with the mode. Approximately 20% of subjects do not have partner information. Since the majority of the participants are living with their partner, we replaced these missing values with 1, which represents living with their partner. For the ROI association analysis, if one subject does not have the WBV, we removed that sample before running the analysis.

**Statistical Models for Association Analysis**

In general, we first examined the association of *APOE*-lifestyle interaction with brain structure according to the models M0-a) and M0-b), as shown in Row 1 of **Table S2**. No significant *APOE*-lifestyle interactions were identified except the *APOE*-ε4-moderate alcohol interaction on the AxD of the superior fronto-occipital fasciculus (**Fig. S20**). Next we only considered the reduced model (M1) as presented in Row 2, **Table S2**, where age-sex-*APOE* three way interaction results were examined. If the three-way interaction was not significant, we reported the effects of the relevant two-way interactions by removing the three-way term; if all interaction terms were not significant, we reported the main effects of the relevant covariates by removing all interaction terms will be reported (models M1 b)-d) in **Table S2**). Type II ANOVA F-test (17) were used to determine the significance of main effects and two-way interaction effects.

Additionally, post hoc analyses in models P1-a1), -a2), -b1), and -b2) in **Table S2** were performed in females and males, separately, to interpret the sex-related interactions. We also considered alternative models for comparison purposes. For example, we coded the APOE gene into two categories: ε4/ε4 and others in the reduced model P2-a), -b), -c) and -d) in **Table S2;** we removed the *APOE* gene from the regression in the reduced models P3-a), -b) and -c) in **Table S2**; we removed the lifestyle and SES factors from the regression in the reduced models P4-a), -b) and -c) in **Table S2**. These reduced models evaluated the confounding effects of *APOE* on the environmental effects and the confounding effects of the environmental factors on the *APOE* effects. The threshold of significance for 211 independent testing after Bonferroni adjustment is approximately *2.37x10-4* (*0.05/211*).

The identified significant results are shown in **Figs. S19** and **S20** for models in M0, in **Figs. S7, S8**, **S9** and **S10** for models in M1, in **Figs. S5** and **S6** for models in P1, **Figs. S21** and **S22** for models in P2, in **Figs**. **S23** and **S24** for models in P3, in **Figs. S25** and **S26** for models in P4. Significant APOE-lifestyle and APOE-SES effects listed in **Figs. S19** and **S20**, and significant main effects of APOE, PRSs, lifestyle and SES factors and effects of age-sex, sex-APOE, age-APOE, age-sex-APOE, age-lifestyle and age-SES interactions listed in **Figs. S19** and **S20** will be counted as our significant signals and summarized in **Figs. S17** and **S18**. Their reproducibility plots based on the UKB non-British ancestry populations were displayed in **Figs. S15** and **S16.**

Correlation plots for genetic and lifestyle factors, 101 brain regional volumes, and 110 DTI diffusion parameters are shown in **Figs. S1-S3**, respectively. We found that correlations between six healthy lifestyle factors were weakly correlated (correlation < 0.1) with each other. The 15 PRS used as covariates in our models were assumed to be normally distributed (**Fig. S4)**.

**References:**

1.	S. W. Choi, T. S.-H. Mak, P. F. O’Reilly, Tutorial: a guide to performing polygenic risk score analyses. Nat. Protoc. 15, 2759–2772 (2020).

2.	T. Otowa, et al., Meta-analysis of genome-wide association studies of anxiety disorders. Mol. Psychiatry 21, 1391–1399 (2016).

3.	E. A. Stahl, et al., Genome-wide association study identifies 30 loci associated with bipolar disorder. Nat. Genet. 51, 793–803 (2019).

4.	B. W. Kunkle, et al., Genetic meta-analysis of diagnosed Alzheimer’s disease identifies new risk loci and implicates Aβ, tau, immunity and lipid processing. Nat. Genet. 51, 414–430 (2019).

5.	D. Demontis, et al., Discovery of the first genome-wide significant risk loci for attention deficit/hyperactivity disorder. Nat. Genet. 51, 63–75 (2019).

6.	J. Grove, et al., Identification of common genetic risk variants for autism spectrum disorder. Nat. Genet. 51, 431–444 (2019).

7.	International Obsessive Compulsive Disorder Foundation Genetics Collaborative (IOCDF-GC) and OCD Collaborative Genetics Association Studies (OCGAS), Revealing the complex genetic architecture of obsessive-compulsive disorder using meta-analysis. Mol. Psychiatry 23, 1181–1188 (2018).

8.	A. J. Forstner, et al., Genome-wide association study of panic disorder reveals genetic overlap with neuroticism and depression. Mol. Psychiatry 26, 4179–4190 (2021).

9.	S. Ripke, et al., Biological insights from 108 schizophrenia-associated genetic loci. Nature 511, 421–427 (2014).

10.	D. Yu, et al., Interrogating the Genetic Determinants of Tourette’s Syndrome and Other Tic Disorders Through Genome-Wide Association Studies. Am. J. Psychiatry 176, 217–227 (2019).

11.	R. K. Walters, et al., Transancestral GWAS of alcohol dependence reveals common genetic underpinnings with psychiatric disorders. Nat. Neurosci. 21, 1656–1669 (2018).

12.	B. Benyamin, et al., Childhood intelligence is heritable, highly polygenic and associated with FNBP1L. Mol. Psychiatry 19, 253–258 (2014).

13.	J. J. Lee, et al., Gene discovery and polygenic prediction from a genome-wide association study of educational attainment in 1.1 million individuals. Nat. Genet. 50, 1112–1121 (2018).

14.	R. Polimanti, et al., Leveraging genome-wide data to investigate differences between opioid use vs. opioid dependence in 41,176 individuals from the Psychiatric Genomics Consortium. Mol. Psychiatry 25, 1673–1687 (2020).

15.	M. J. H. M. van der Loos, et al., The Molecular Genetic Architecture of Self-Employment. PLoS ONE 8, e60542 (2013).

16.	M. Liu, et al., Association studies of up to 1.2 million individuals yield new insights into the genetic etiology of tobacco and alcohol use. Nat. Genet. 51, 237–244 (2019).

17.	Ø. Langsrud, ANOVA for unbalanced data: Use Type II instead of Type III sums of squares. Stat. Comput. 13, 163–167 (2003).

18. ,	UKB primary brain imaging document (September 2, 2022).

19.	B. Zhao, et al., Genome-wide association analysis of 19,629 individuals identifies variants influencing regional brain volumes and refines their genetic co-architecture with cognitive and mental health traits. Nat. Genet. 51, 1637–1644 (2019).

20.	B. B. Avants, et al., A reproducible evaluation of ANTs similarity metric performance in brain image registration. NeuroImage 54, 2033–2044 (2011).

21.	N. J. Tustison, et al., Large-scale evaluation of ANTs and FreeSurfer cortical thickness measurements. NeuroImage 99, 166–179 (2014).

22.	B. Zhao, et al., Common genetic variation influencing human white matter microstructure. Science 372, eabf3736 (2021).

23.	A. Manichaikul, et al., Robust relationship inference in genome-wide association studies. Bioinformatics 26, 2867–2873 (2010).

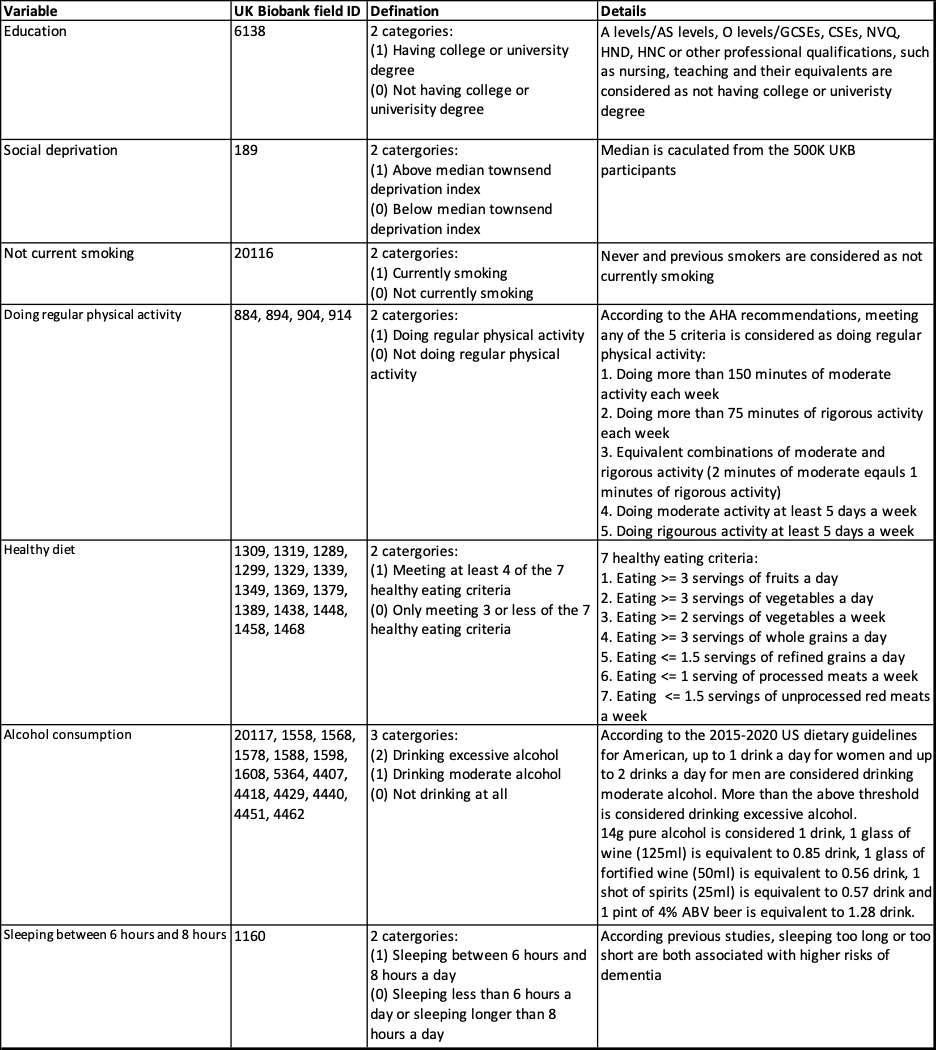

**Supplementary Table S1.** Criteria for the SES variables and lifestyle factors generated from the UKB data fields. All variables are measured at the baseline.

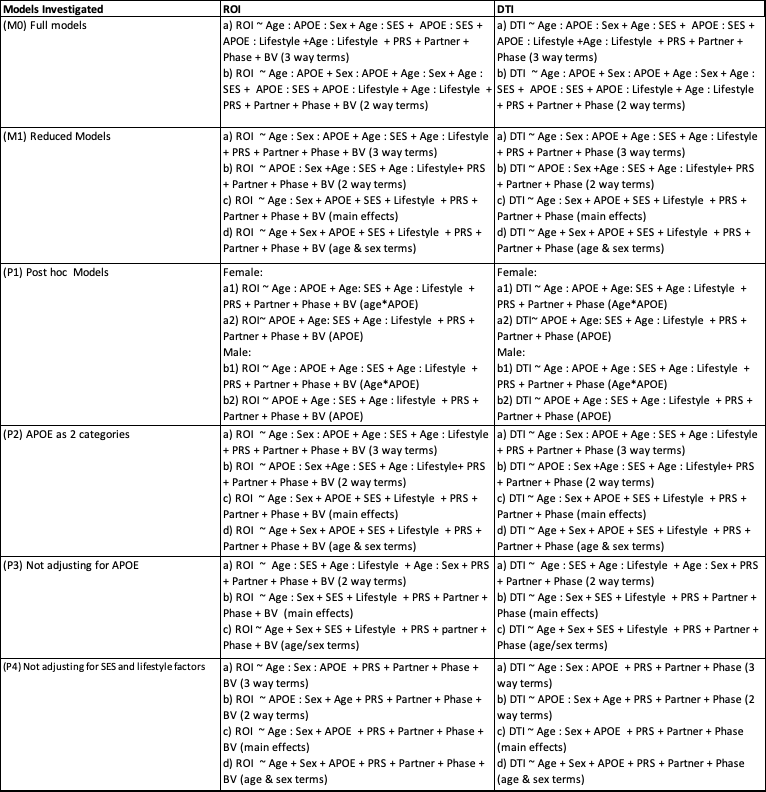

**Supplementary Table S2.** Models investigated and results are displayed in the following heatmaps. Throughout all the models, a three-way interaction term implied that all two-way interactions and main effects of covariates relevant to this three-way interaction term were also included; a two-way interaction term implied that all main effects of the covariates relevant to this two-way interaction term were also included. “A:B” denotes the interaction of covariates A and B; “A:B:C” denotes the three-way interaction of covariates A, B, and C. PRS, polygenic risk score; ROI, regions of interest; SES, socioeconomic status; DTI, diffusion tensor imaging phenotype; BV, whole brain volume.

**Figures**

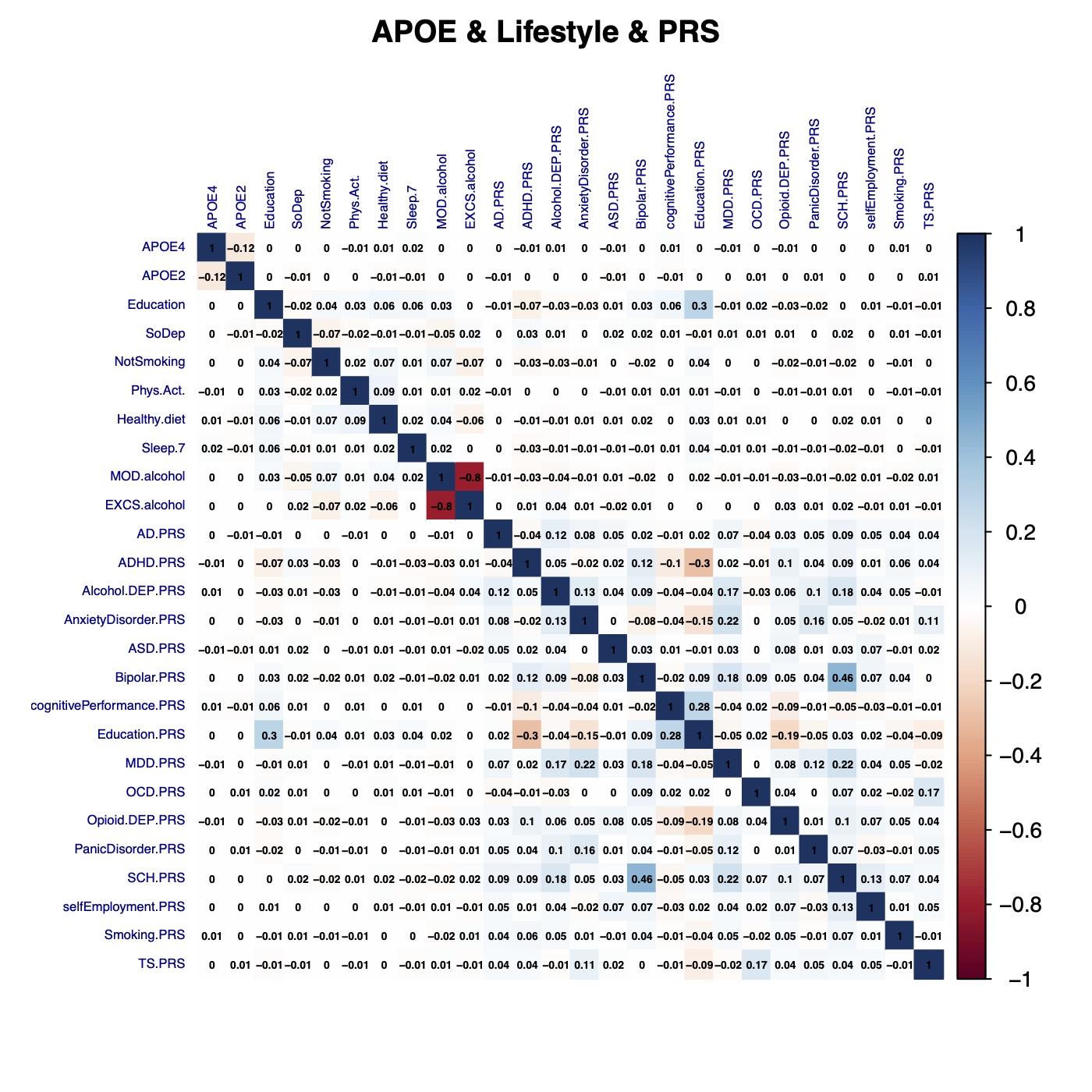

**Supplementary Figure S1** Correlation plot of APOE, PRSs and lifestyle variables. SoDep, Social deprivation Index; NotSmoking, not-current-smoker; Phys.Act, physical activity; Healthy.diet, healthy diet; Sleep.7, healthy sleep; MOD.alcohol, moderate alcohol; EXCS.alcohol, excessive alcohol; PRS, polygenic risk score; AD.PRS, PRS for Alzheimer’s Disease; ADHD.PRS, PRS for attention deficit hyper-activity disorder; Alcohol.DEP.PRS, PRS for alcohol dependence; AnxietyDisorder.PRS, PRS for anxiety disorder; ASD.PRS, PRS for autism spectrum disorder; Bipolar.PRS, PRS for bipolar disorder; cognitivePerformance.PRS, PRS for cognitive performance; Education.PRS, PRS for education; MDD.PRS, PRS for major depression disorder; OCD.PRS, PRS for obsessive-compulsive disorder; Opioid.DEP.PRS, PRS for opioid dependence; SCH.PRS, PRS for schizophrenia; selfEmployment.PRS, PRS for self employment; Smoking.PRS, PRS for heavy smoking; TS.PRS, PRS for Tourette syndrome.

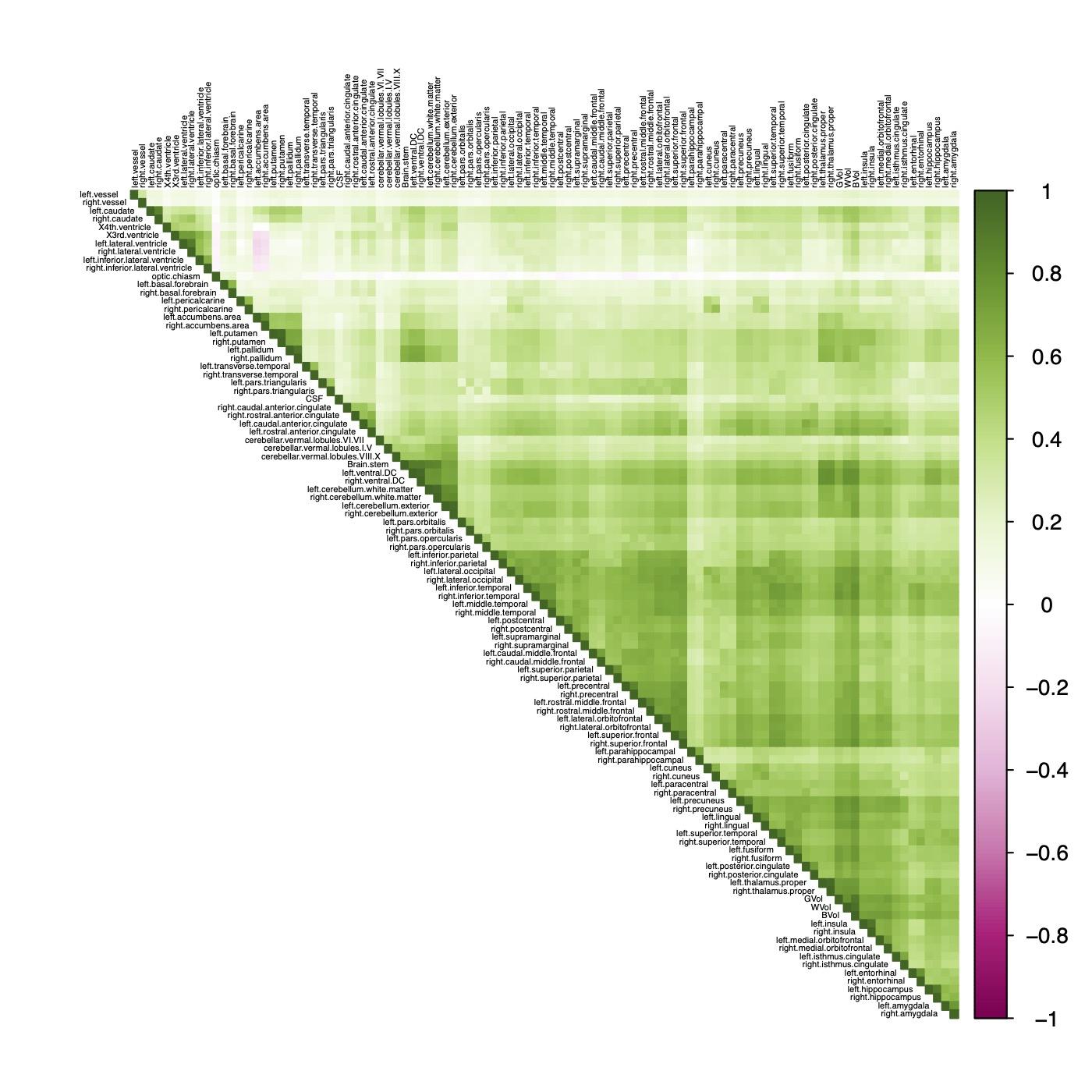

**Supplementary Figure S2** Correlation plot of 101 brain regional volumes (n = 37,939).

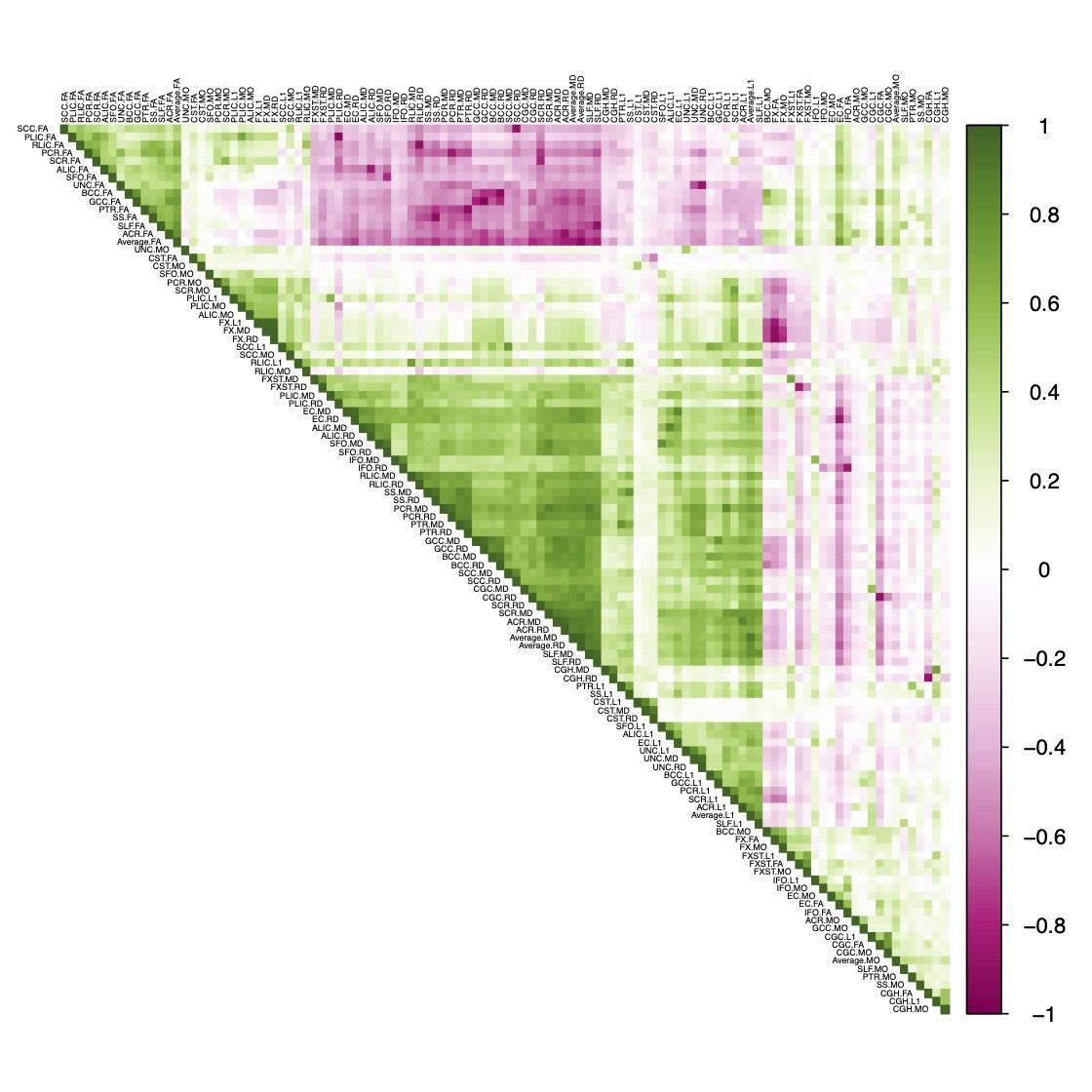

**Supplementary Figure S3** Correlation plot of 110 diffusion tensor imaging phenotypes (n = 35,921).

**(A)**

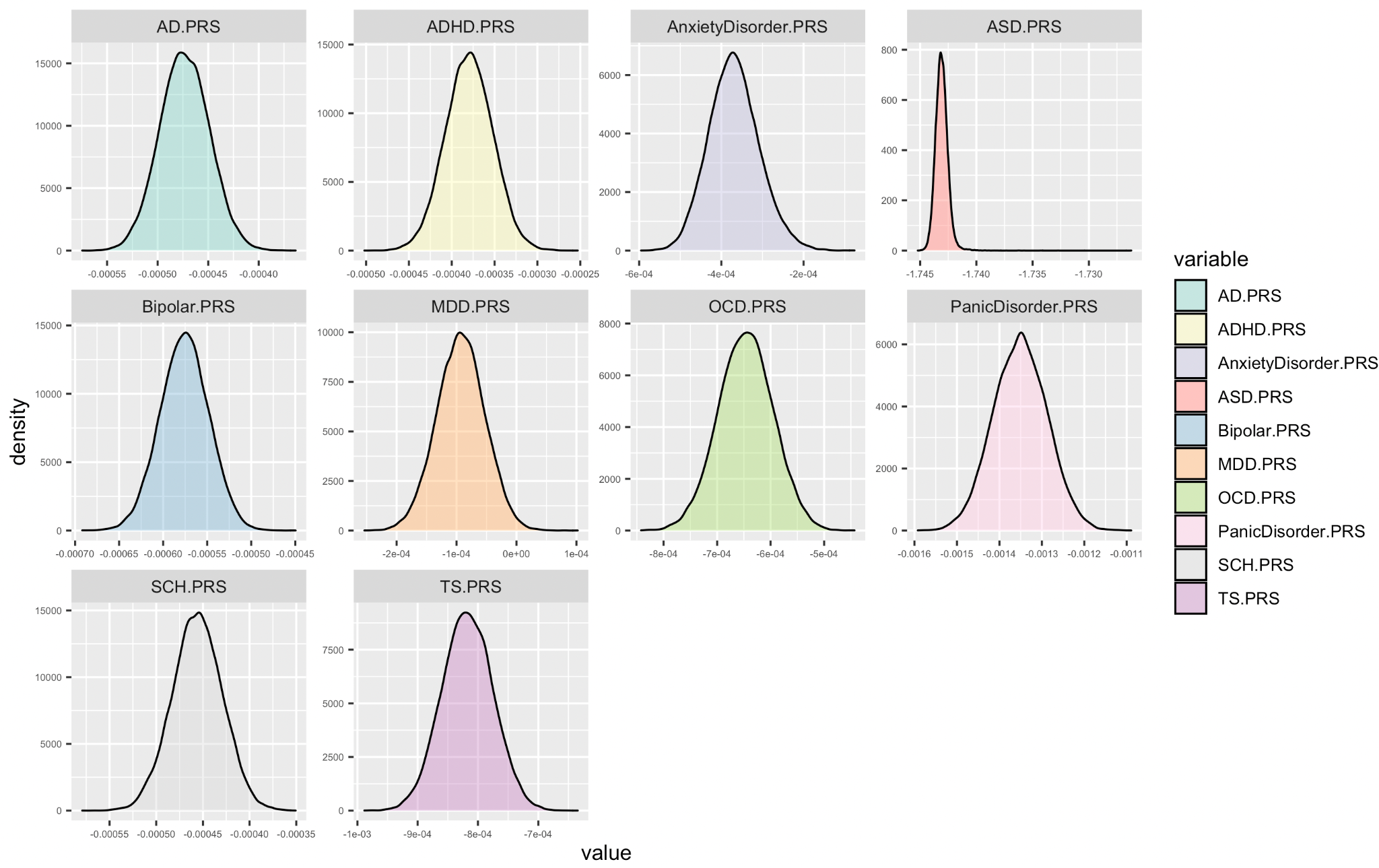

（B）

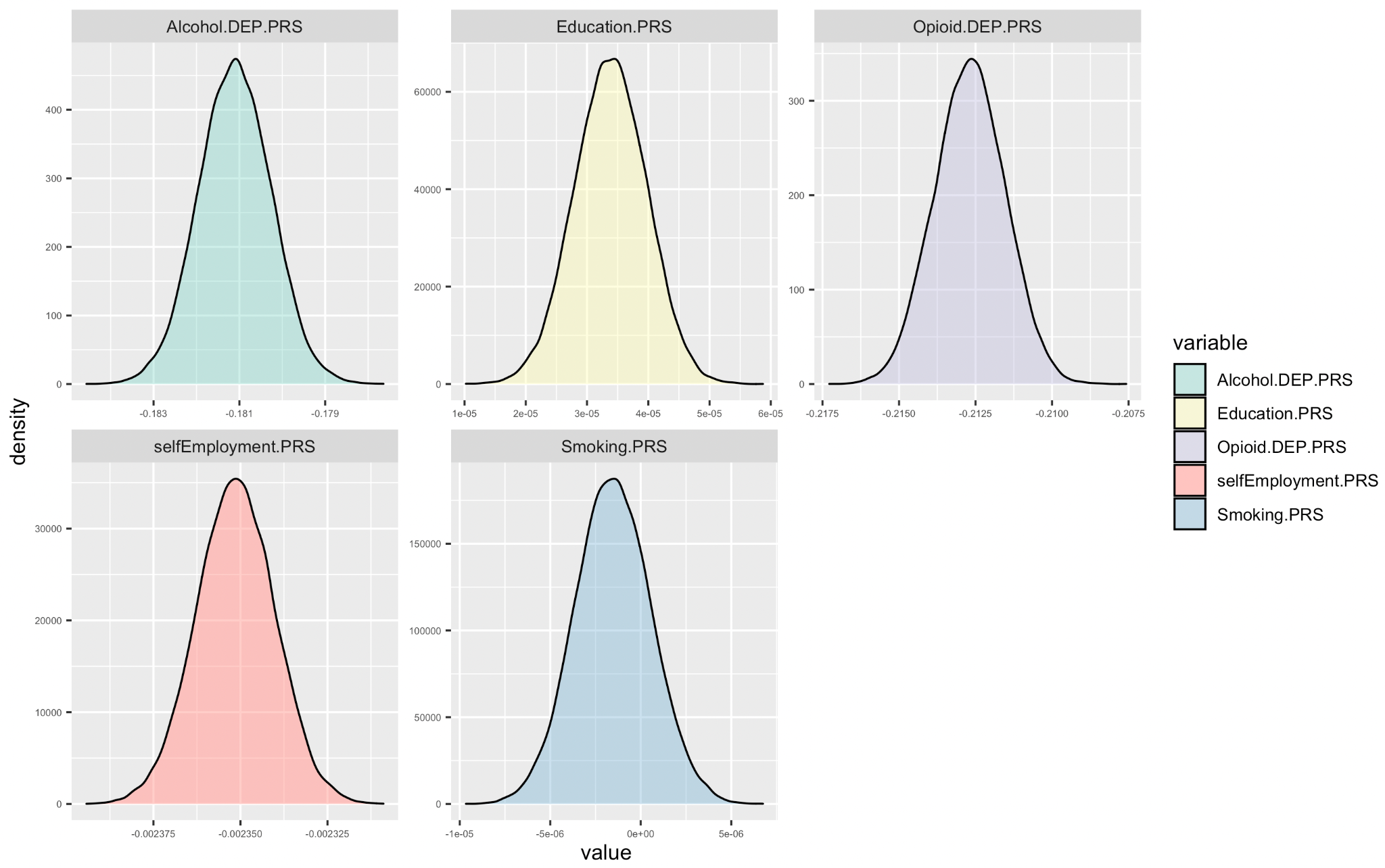

**Supplementary Figure S4** Polygenic risk scores (PRS) distributions of the White British subjects. **(A)** for 10 disease polygenic scores. **(B)** for 5 behavioral polygenic scores. AD, Alzheimer’s Disease; ADHD, attention deficit hyper-activity disorder; Alcohol.DEP, alcohol dependence; ASD, autism spectrum disorder; Bipolar, bipolar disorder; MDD, major depression disorder; OCD, obsessive-compulsive disorder; Opioid.DEP, opioid dependence; SCH, schizophrenia; TS, Tourette syndrome.

Female

Male

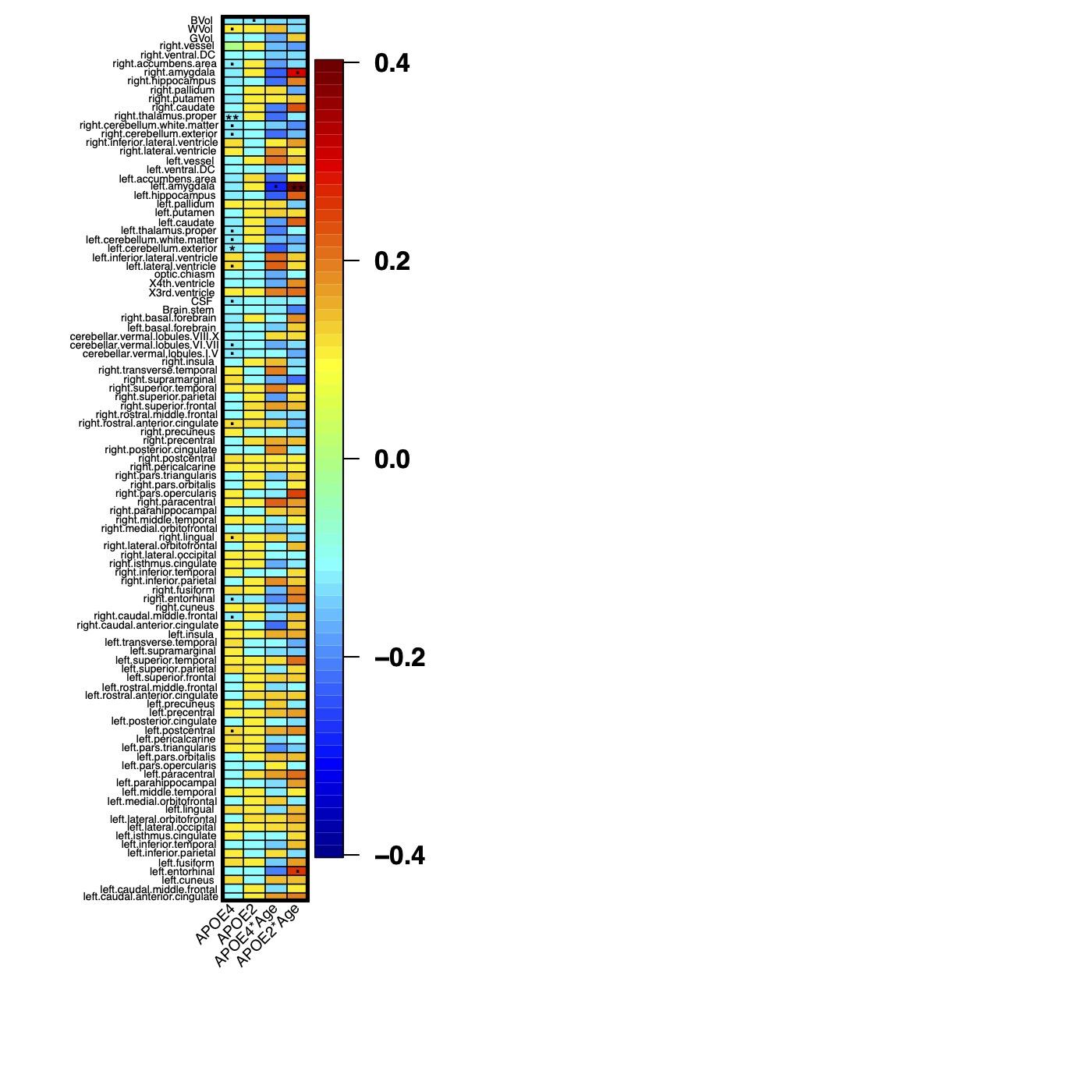

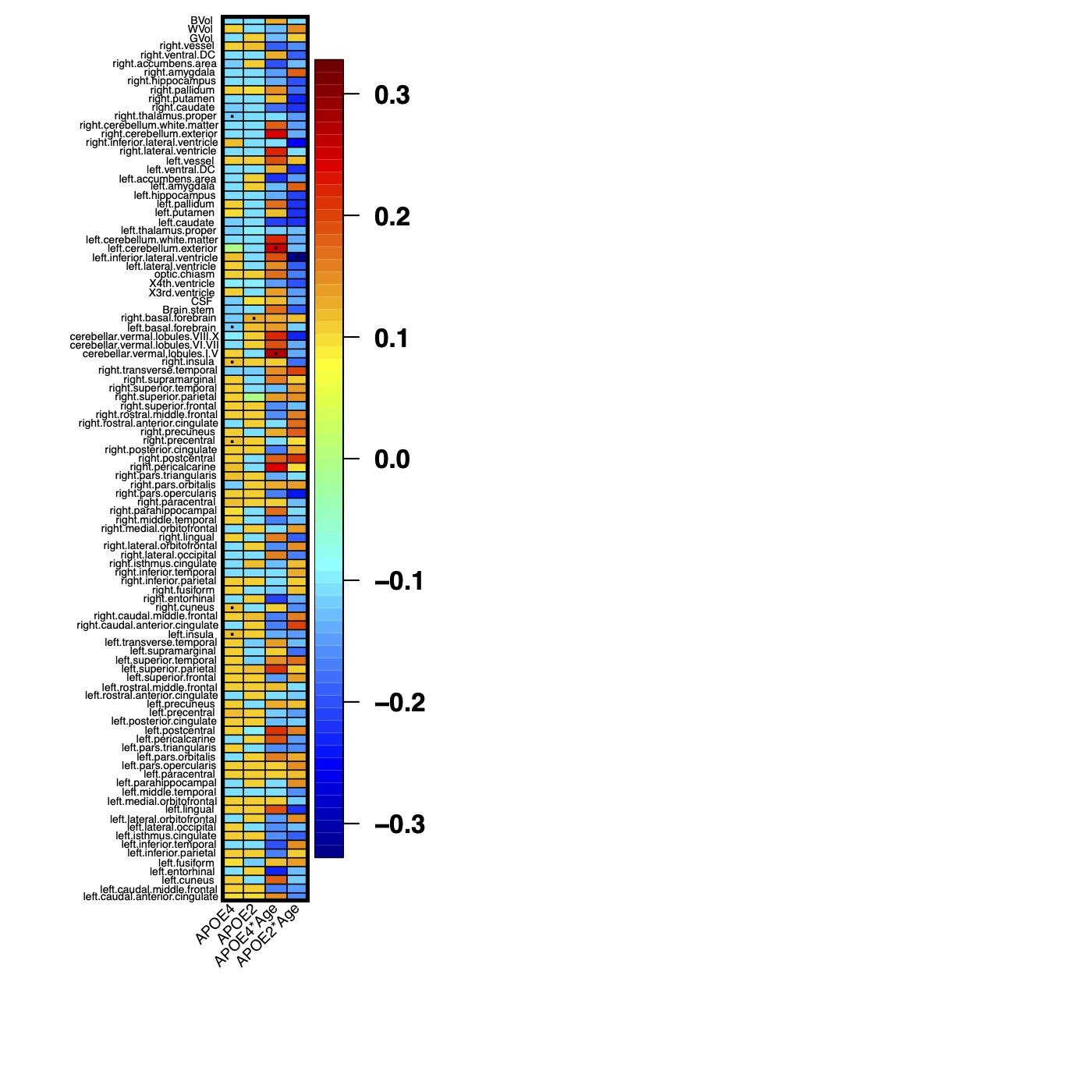

**Supplementary Figure S5** The effect size of *APOE*-ε2, *APOE*-ε4 and their age interaction terms on brain regional volumes in females (Left Panel) and males (Right Panel) (based on model P1, **Table S2**). Results that passed *0.01*, *2.37E-4* and *4.74E-5* significance levels are denoted as (.), (*) and (**), respectively.

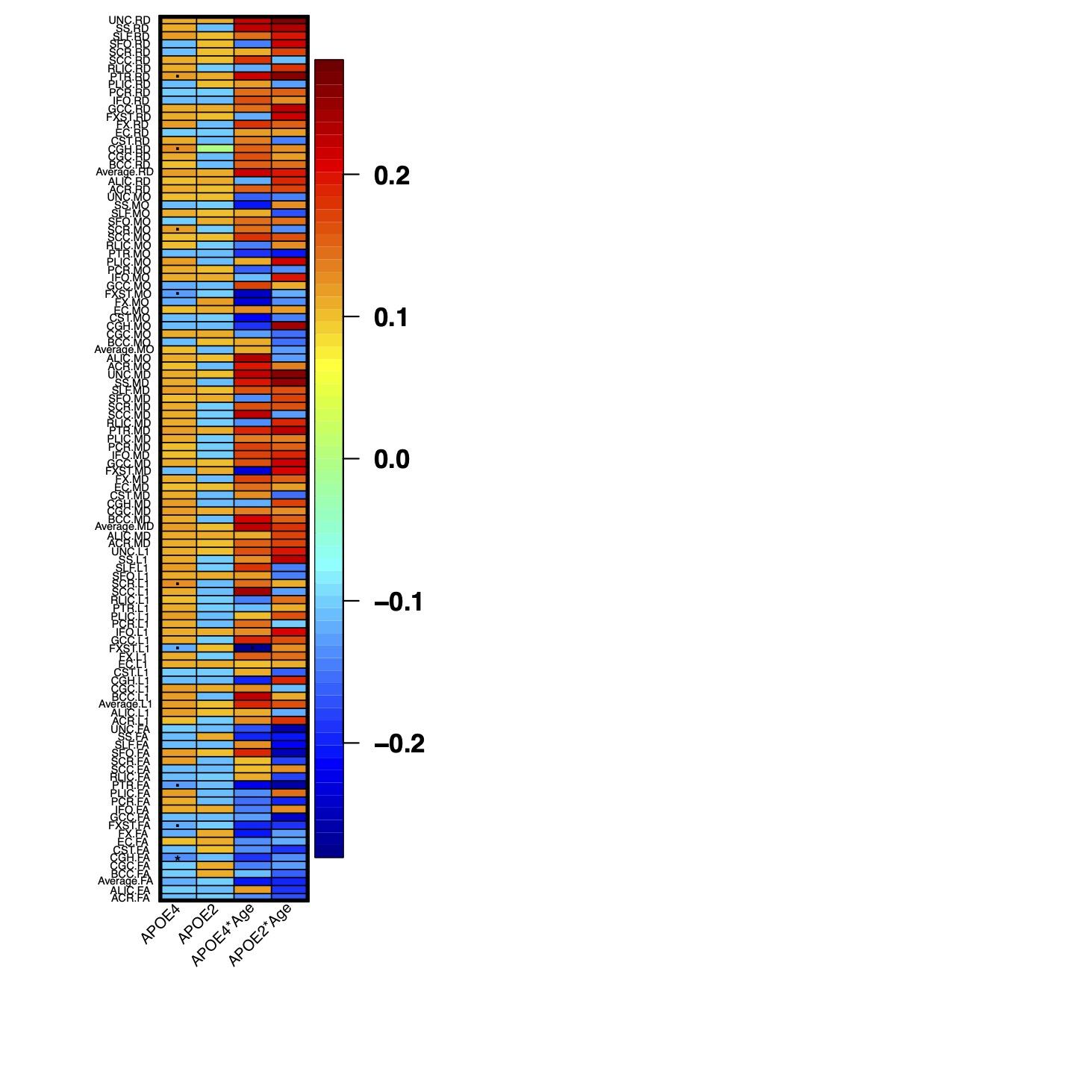

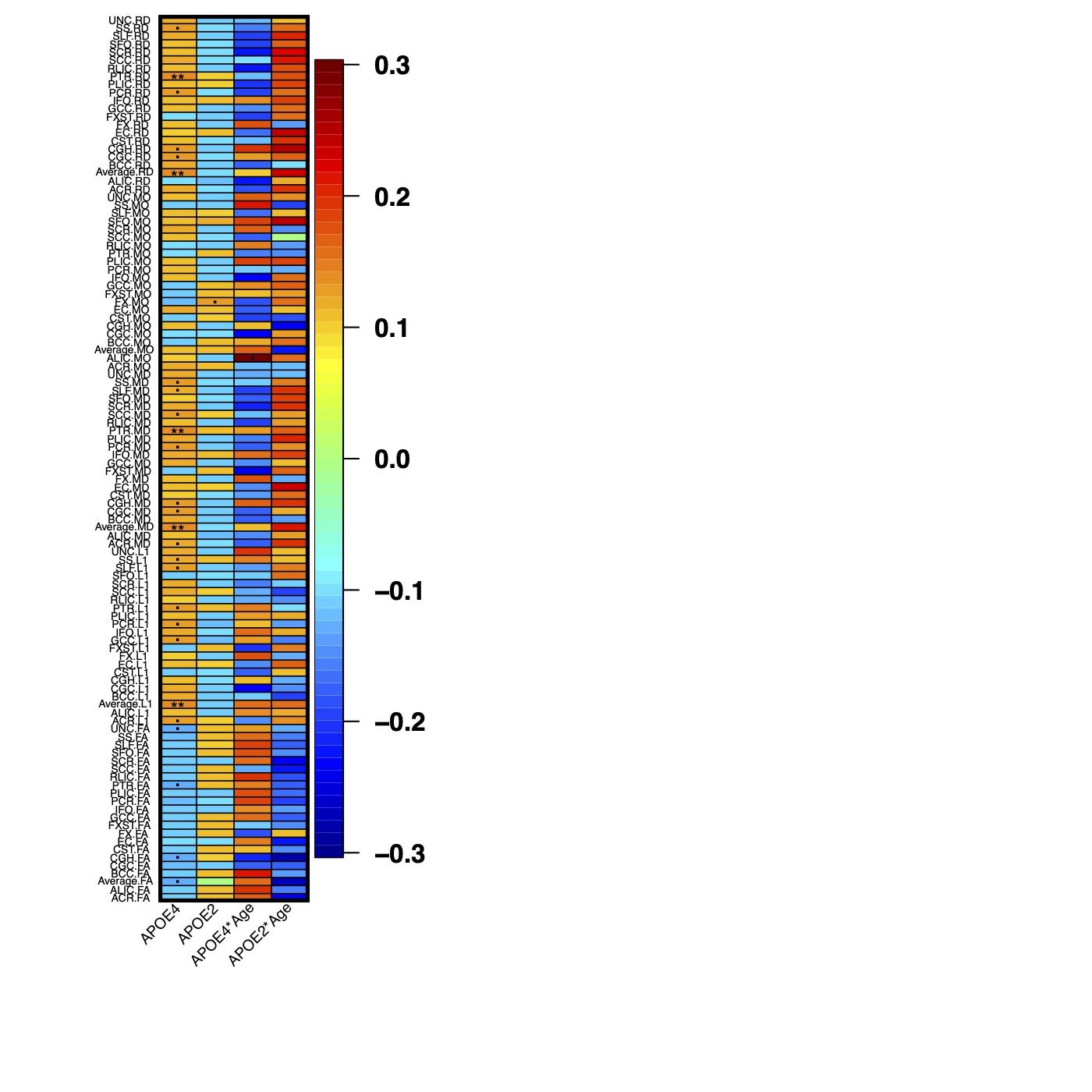

Female

Male

**Supplementary Figure S6** The effect size of *APOE*-ε2, *APOE*-ε4 and their age interaction terms on diffusion tensor imaging measurements in females (Left Panel) and males (Right Panel), respectively (based on model P1, **Table S2**). Results that passed *0.01*, *2.37E-4* and *4.74E-5* significance levels are denoted as (.), (*) and (**), respectively.

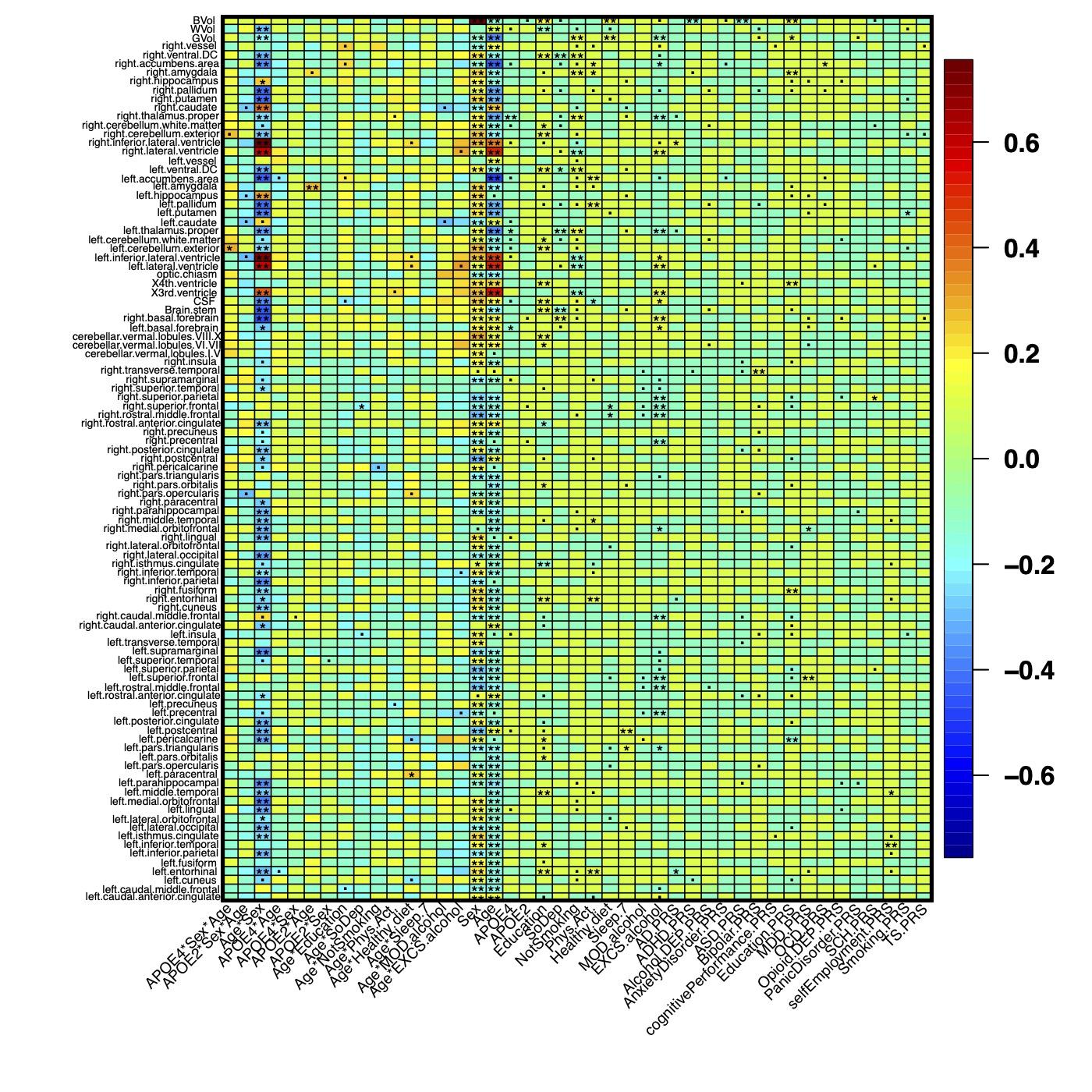

**Supplementary Figure S7.** Heatmap of all the identified effects on ROI volumes (based on model M1, **Table S2**). Results that passed *0.01*, *2.37E-4* and *4.74E-5* significance levels are denoted as (.), (*) and (**), respectively.

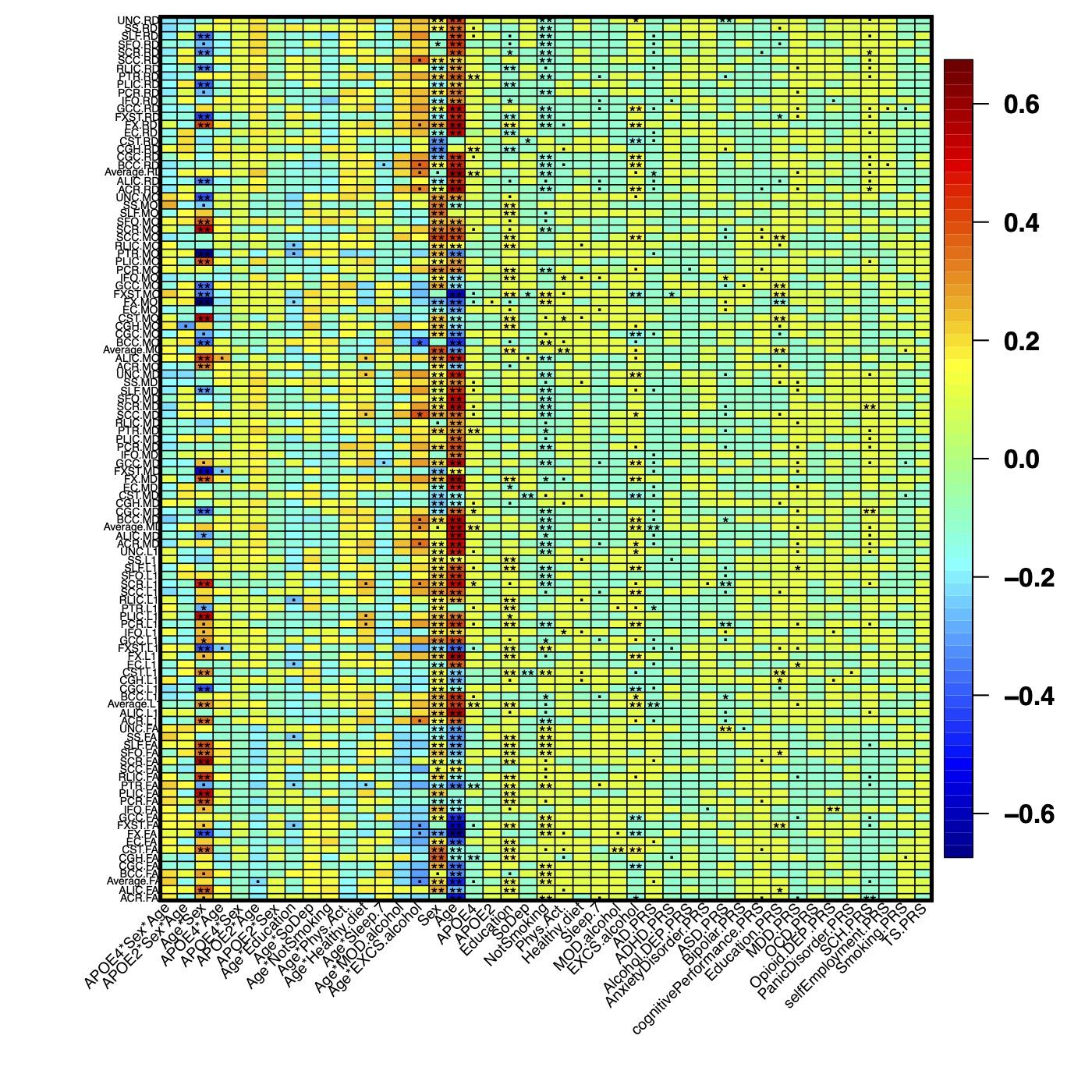

**Supplementary Figure S8.** Heatmap of all the identified effects on DTI measurements (based on model M1, **Table S2**) Results that passed *0.01*, *2.37E-4* and *4.74E-5* significance levels are denoted as (.), (*) and (**), respectively.

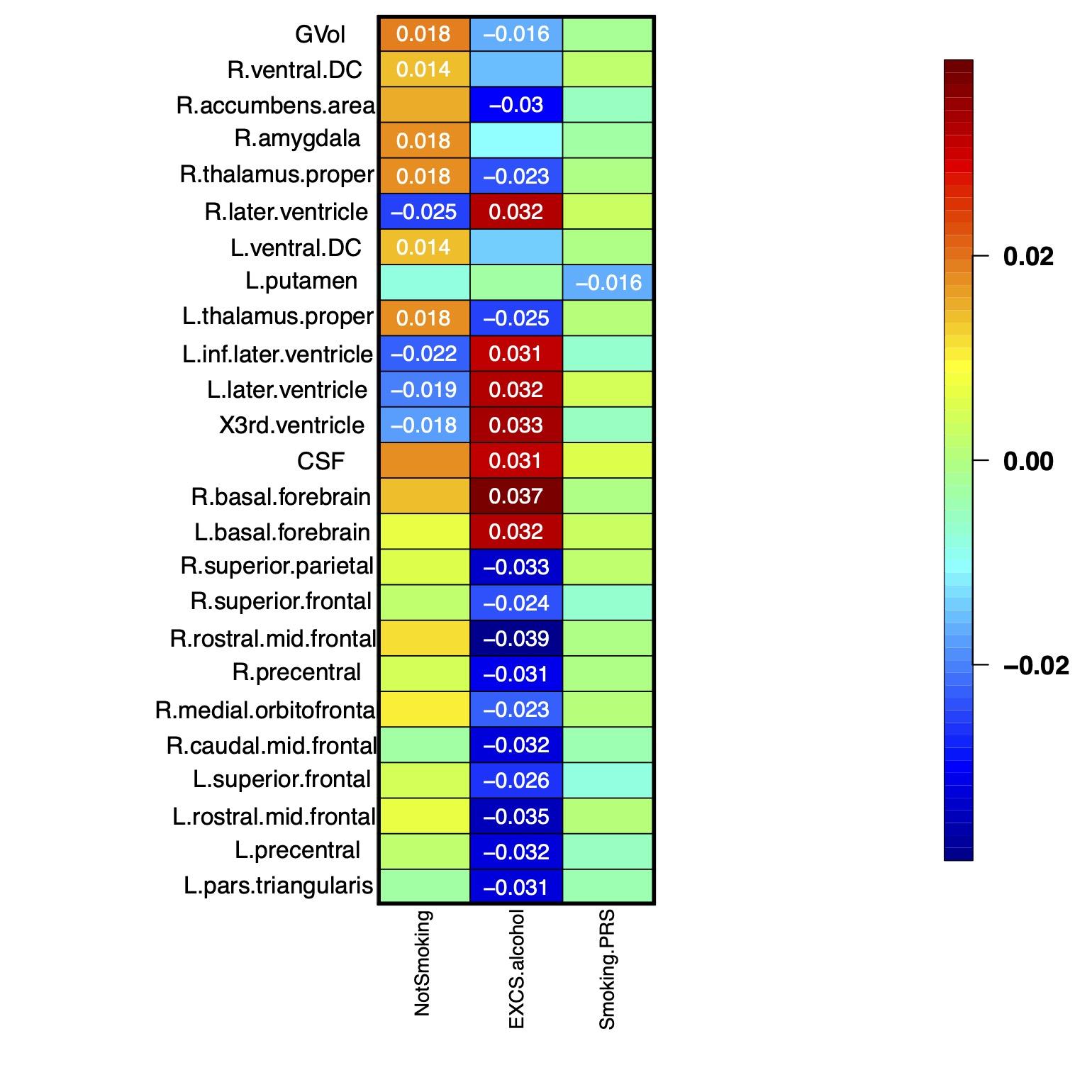

**Supplementary Figure S9** Heatmap of the identified main effects (standardized coefficients) of smoking, alcohol and smoking polygenic scores on brain regional volumes (based on model M1, **Table S2**). Only results that passed the *2.37E-4* significance level were shown.

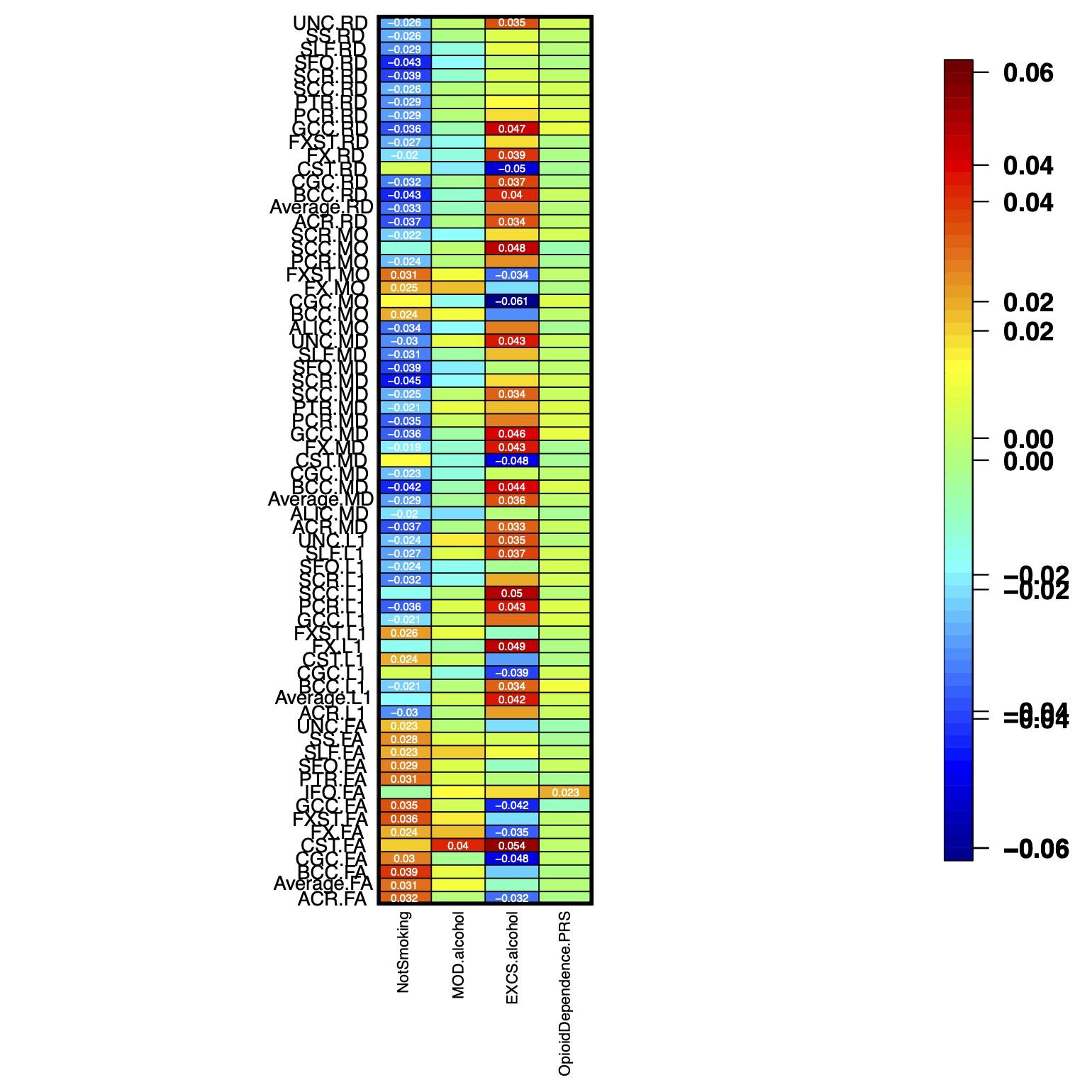

**Supplementary Figure S10** Heatmap of the identified main effects (standardized coefficients) of smoking, alcohol and opioid on DTI measurements (based on model M1, **Table S2**). Only results that passed the *2.37E-4* significance level were shown.

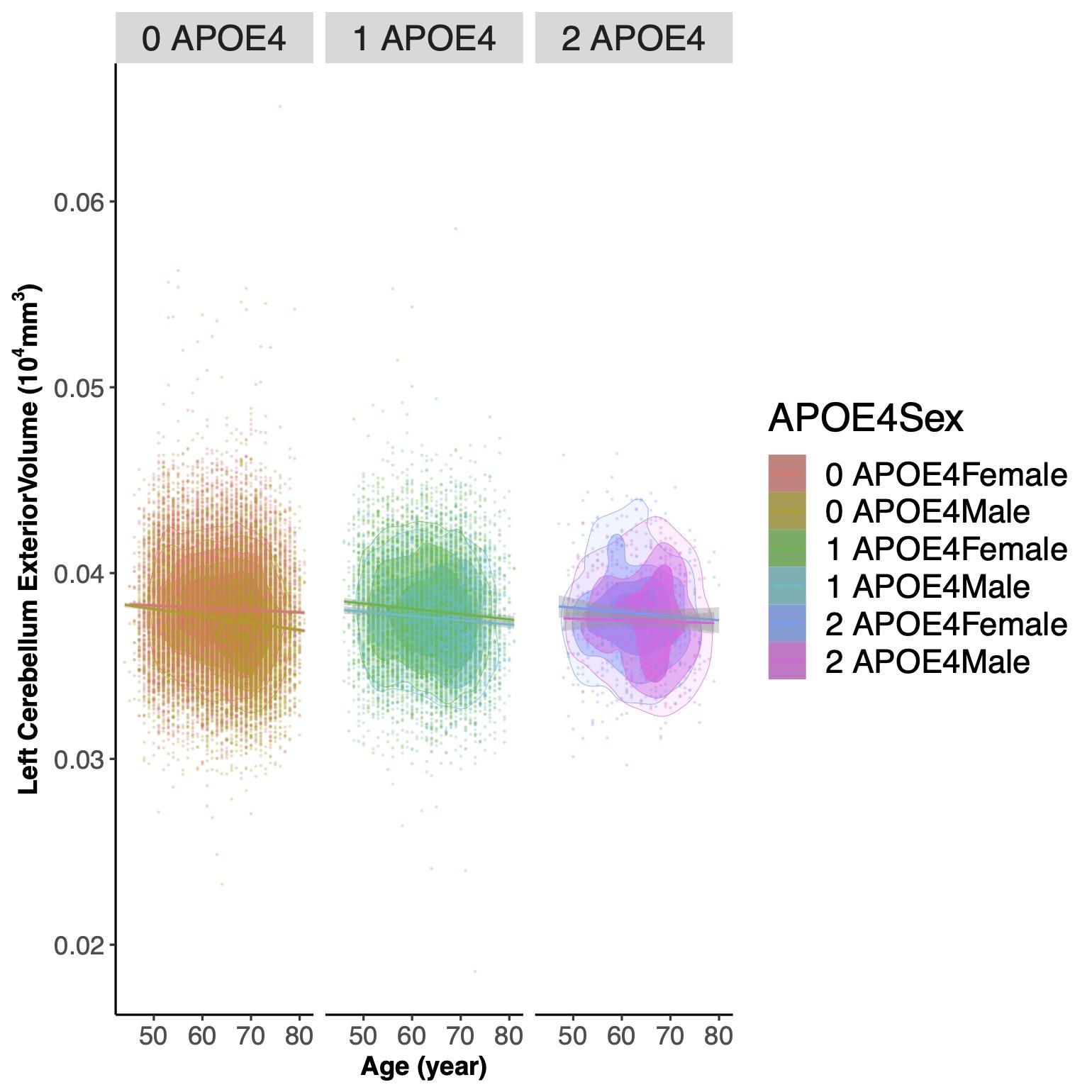

**Supplementary Figure S11** The scatterplots of left cerebellum exterior volume against age grouped by sex and *APOE*-ε4. Contour plots are used to show the density of two-dimensional distributions; regression lines and 95% confidence intervals were also drawn.

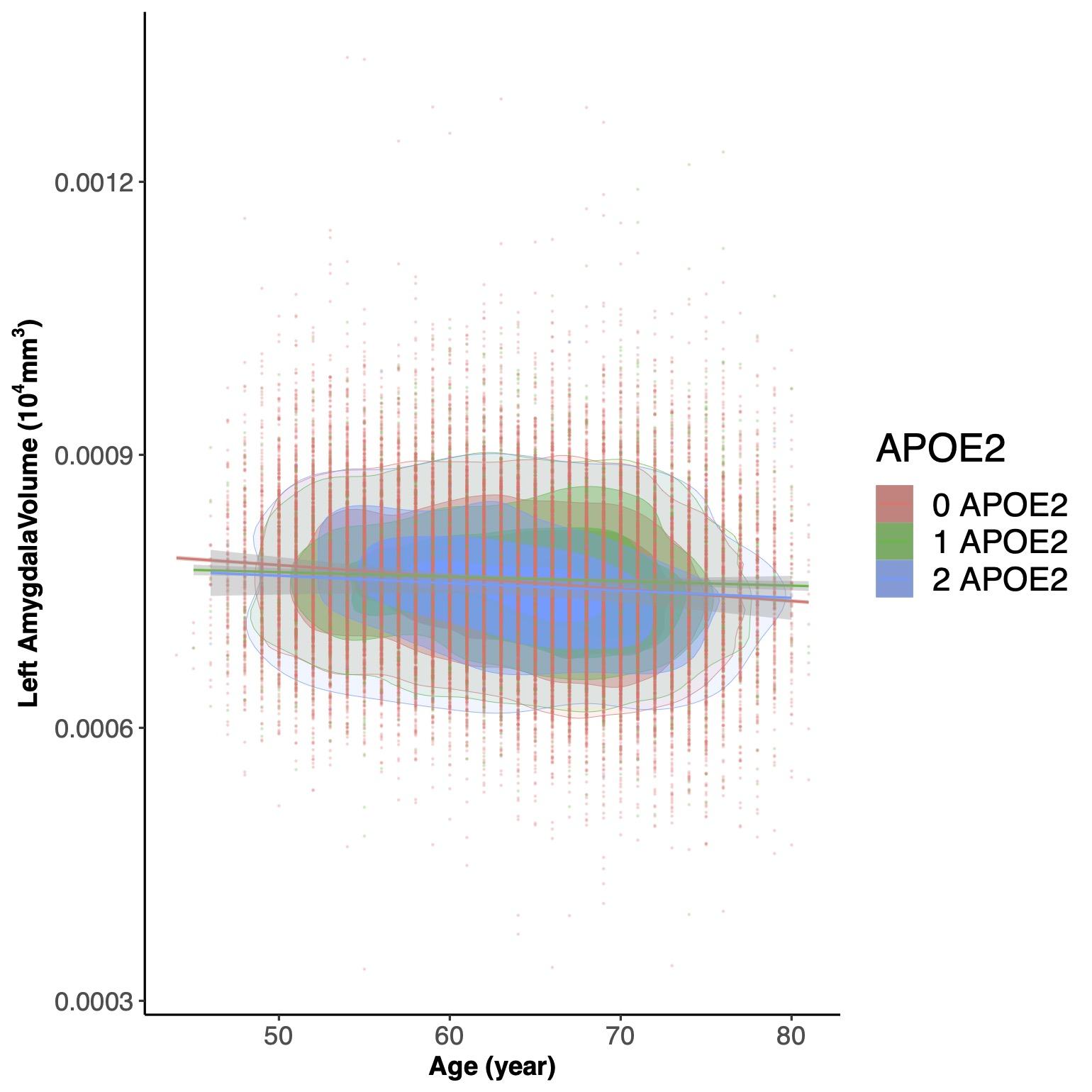

**Supplementary Figure S12** The contour plot of the left Amygdala volume against age grouped by sex. Contour plots are used to show the density of two-dimensional distributions; regression lines and 95% confidence intervals were also drawn.

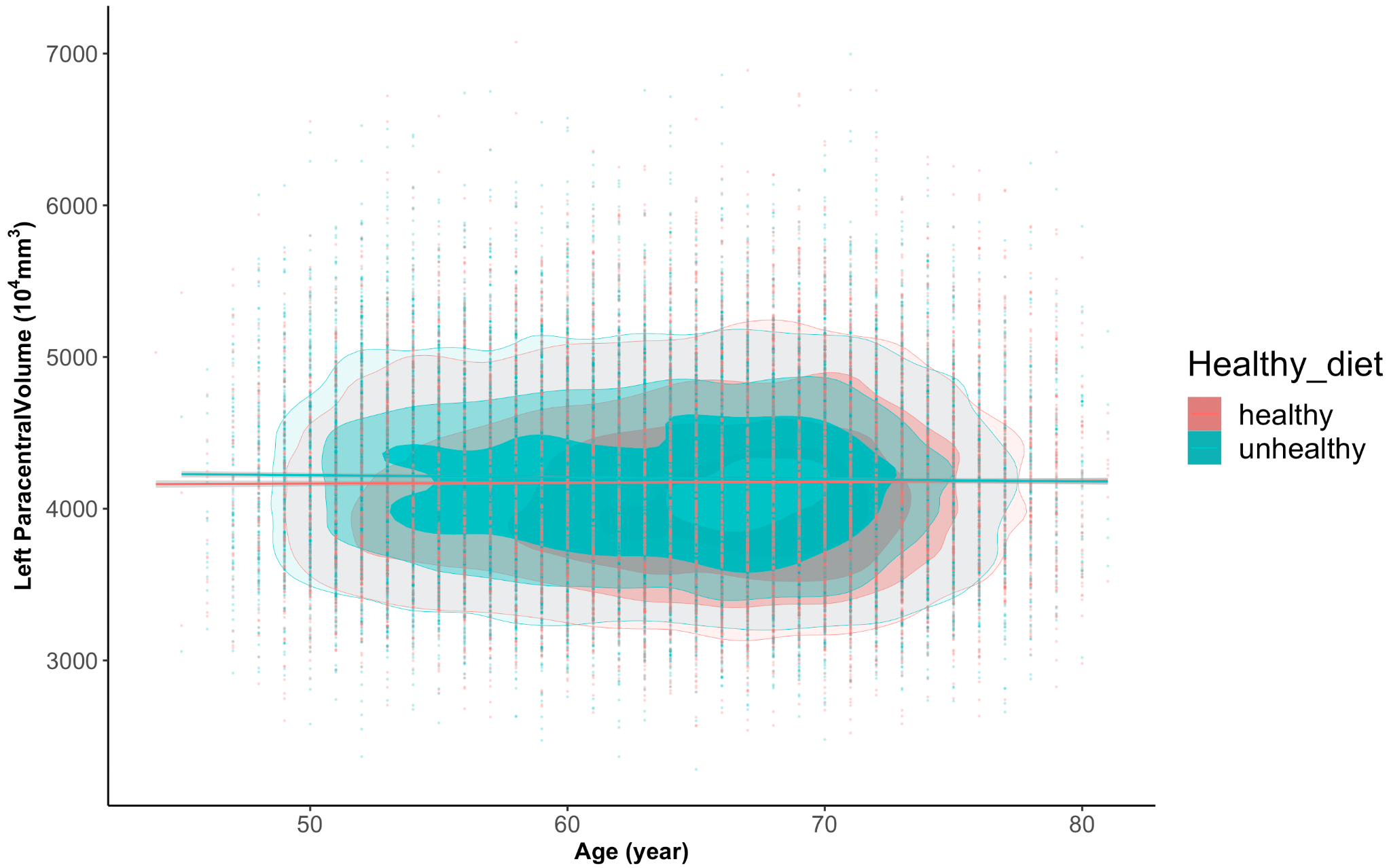

**Supplementary Figure S13** The contour plot of the left Paracentral volume against age grouped by eating habits. Contour plots are used to show the density of two-dimensional distributions; regression lines and 95% confidence intervals were also drawn.

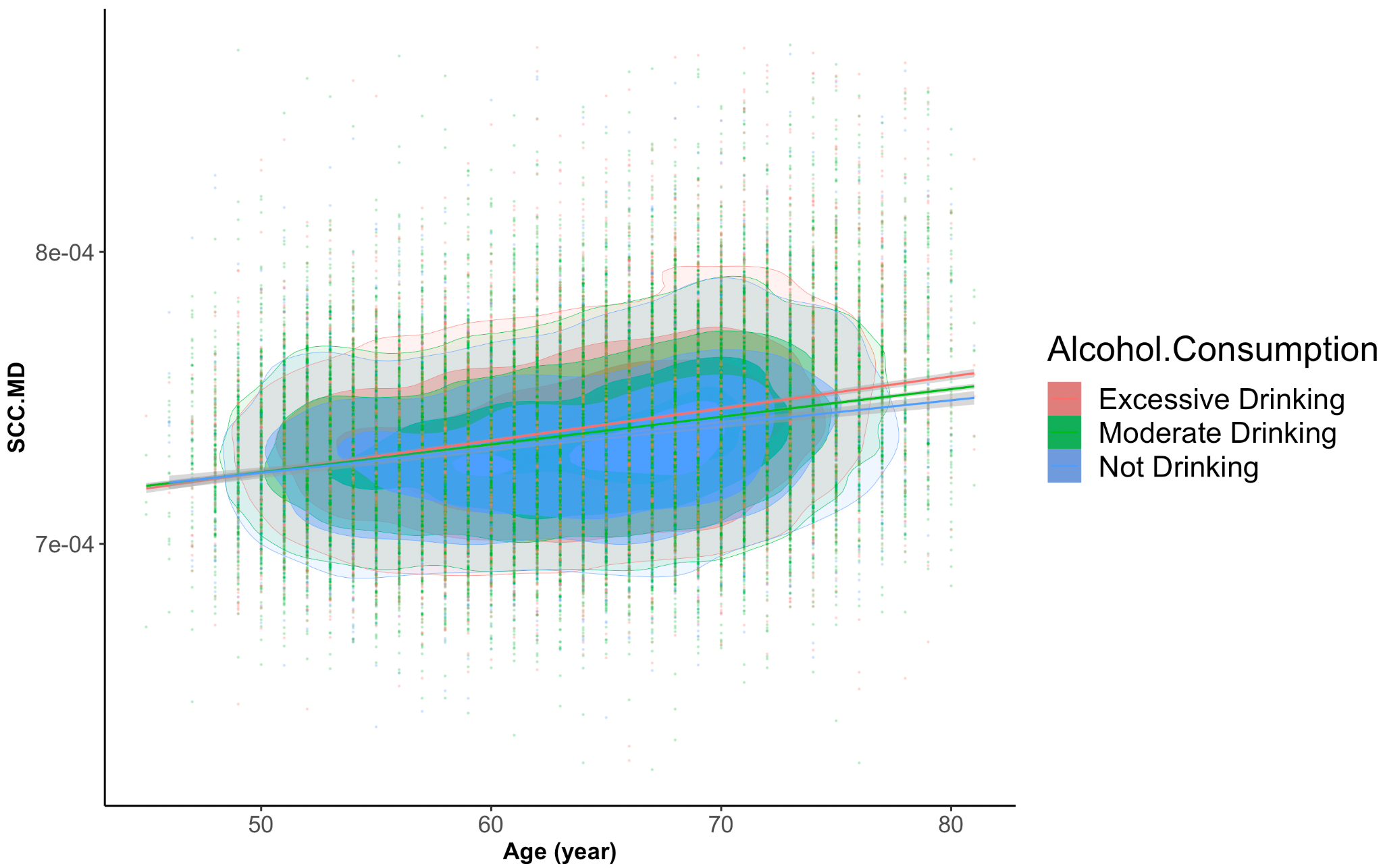

**Supplementary Figure S14** The contour plot of the SCC.MD against age grouped by alcohol consumption. Contour plots are used to show the density of two-dimensional distributions; regression lines and 95% confidence intervals were also drawn

**Reproducibility**

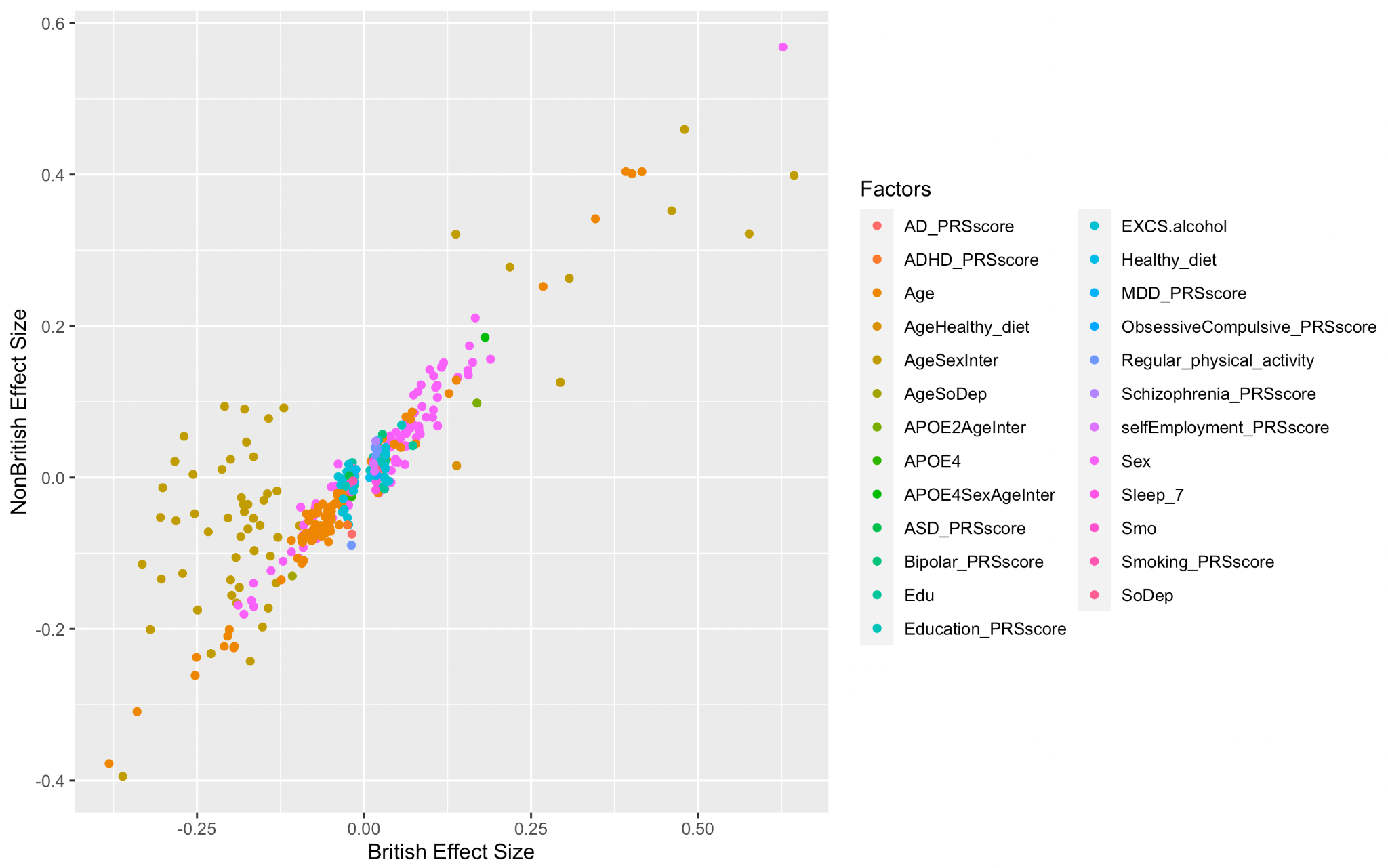

**Supplementary Figure S15** Plot of effects on ROI volumes identified in the main discovery (n= 35,021) versus the effect size in non-British dataset (n = 2,918).

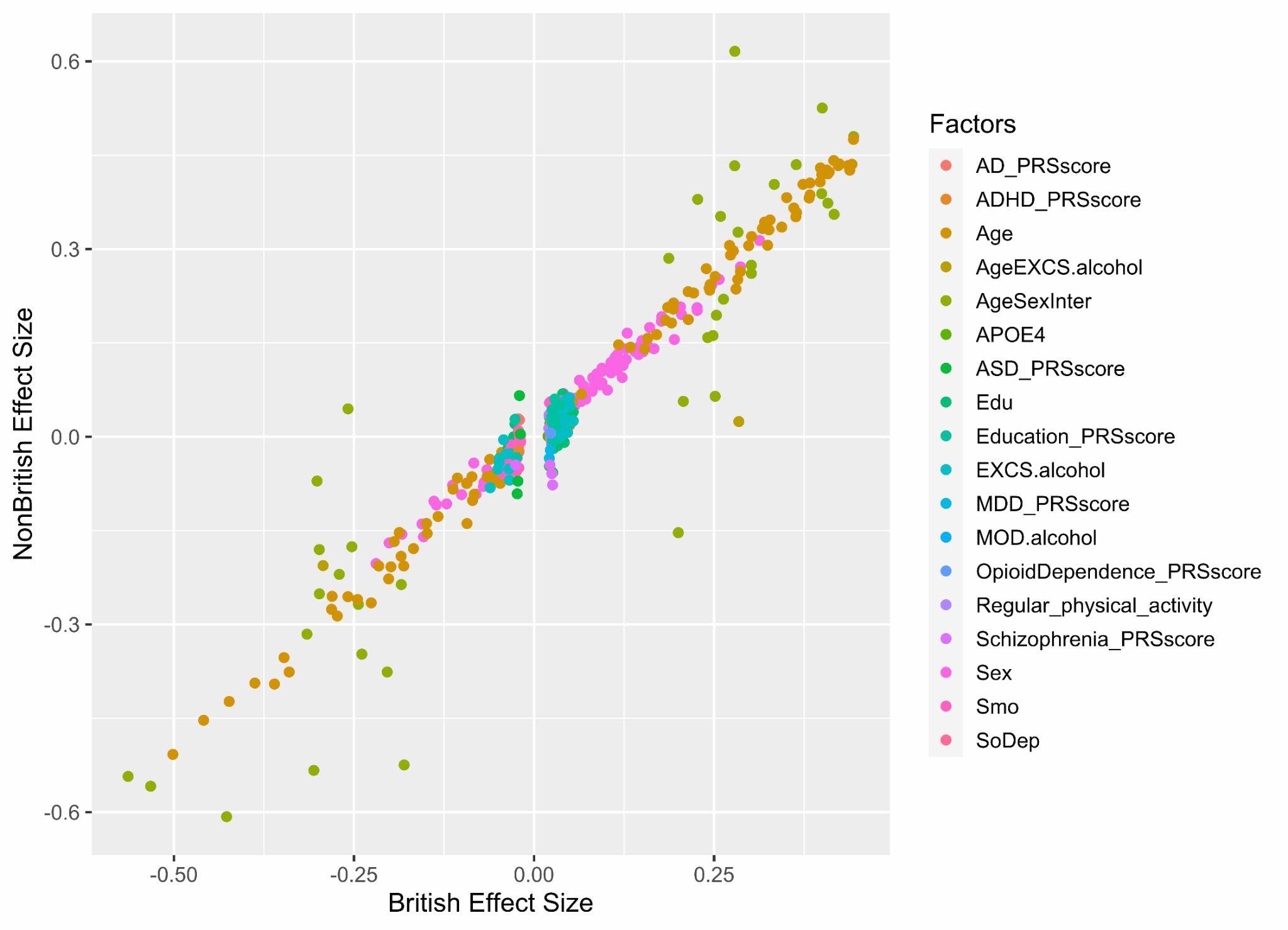

**Supplementary Figure S16** Plot of effects on DTI measurements identified in the main discovery (n = 33,167) versus the effect size in non-British dataset (n = 2,754) (based on model M1, **Table S2**).

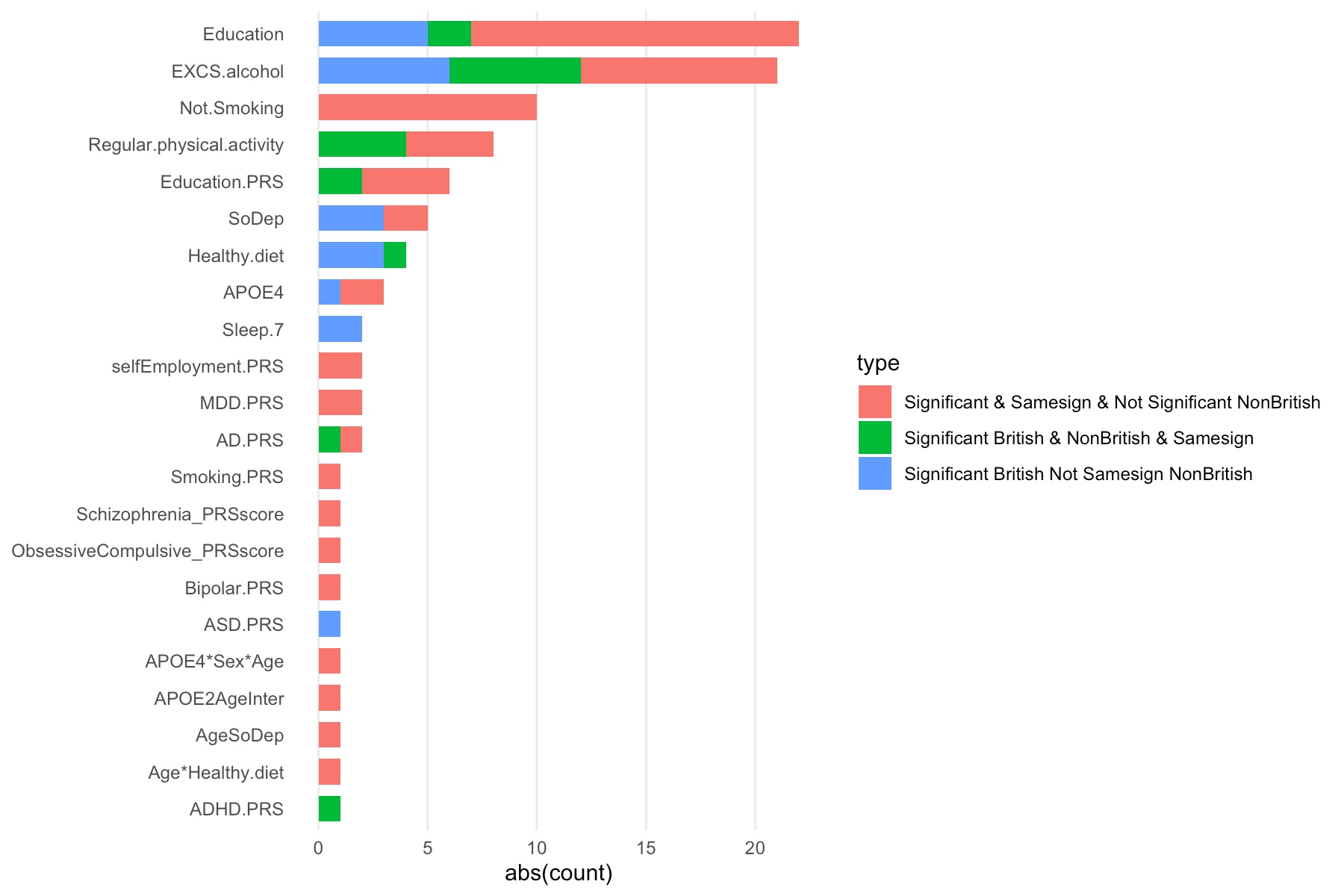

**Supplementary Figure S17** Barplots of effects on brain ROI volume identified in the main discovery (n = 35,021) and effects identified in non-British dataset (n = 2918).

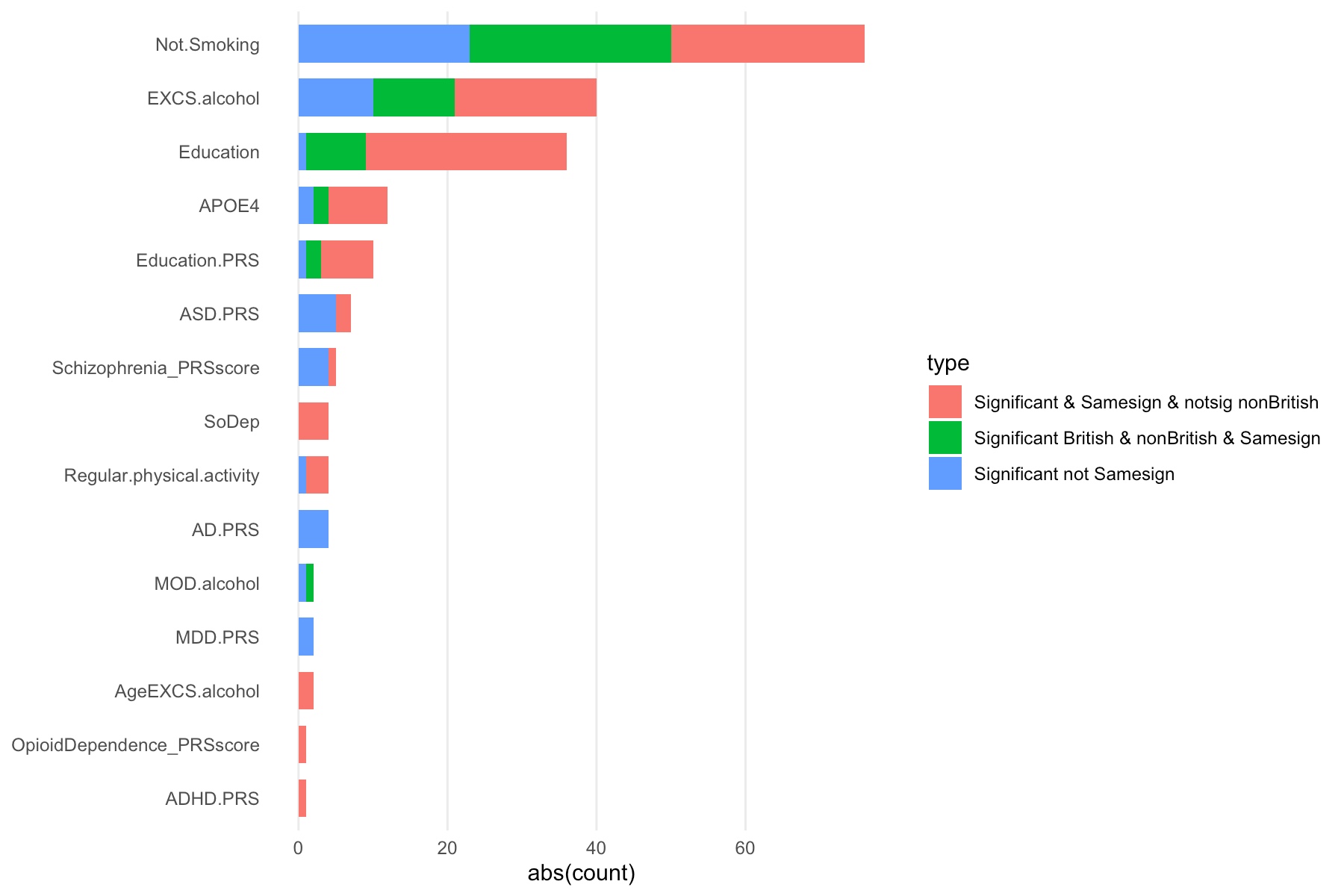

**Supplementary Figure S18** Barplots of effects identified on white matter diffusion parameters in the main discovery (n = 33,167) and effects identified in non-British dataset (n = 2,754).

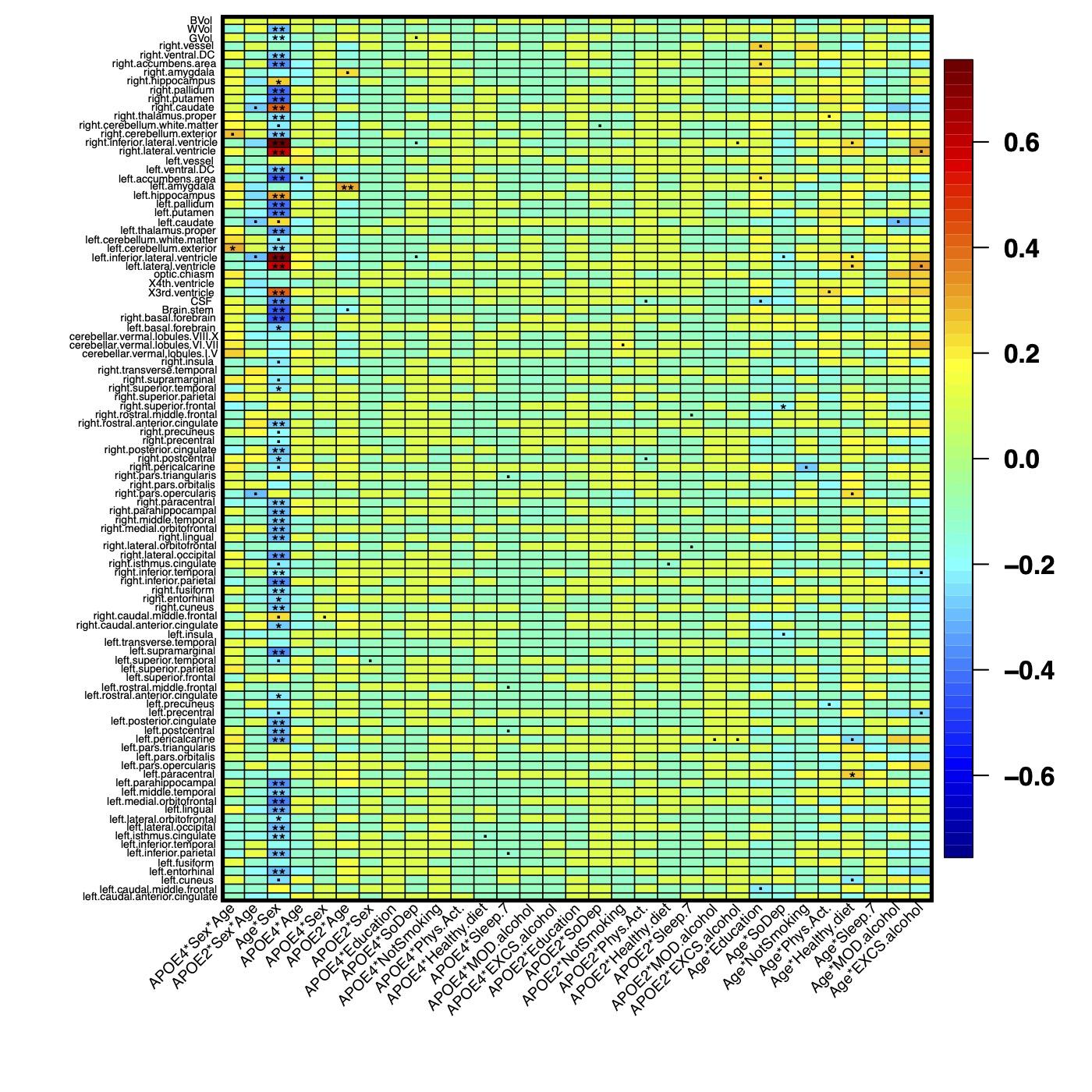

**Supplementary Figure S19.** Heatmap of the interaction effects of *APOE* and lifestyle factors on ROI volumes based on model M0 in **Table S2**. Results that passed *0.01*, *2.37E-4* and *4.74E-5* significance levels are denoted as (.), (*) and (**), respectively.

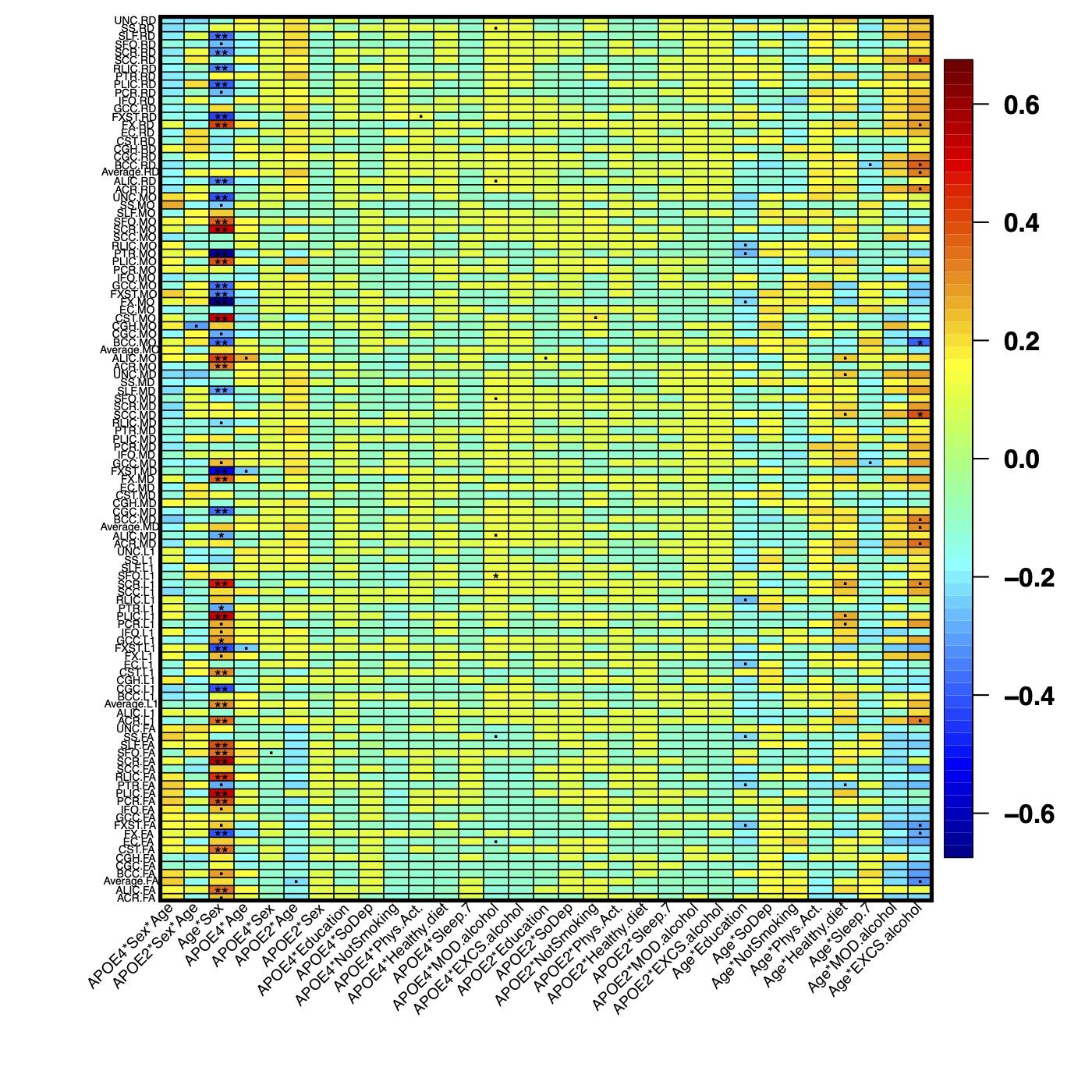

**Supplementary Figure S20.** Heatmap of the interaction effects of *APOE* and lifestyle factors on DTI measurements based on model M0 in **Table S2**. Results that passed *0.01*, *2.37E-4* and *4.74E-5* significance levels are denoted as (.), (*) and (**), respectively.

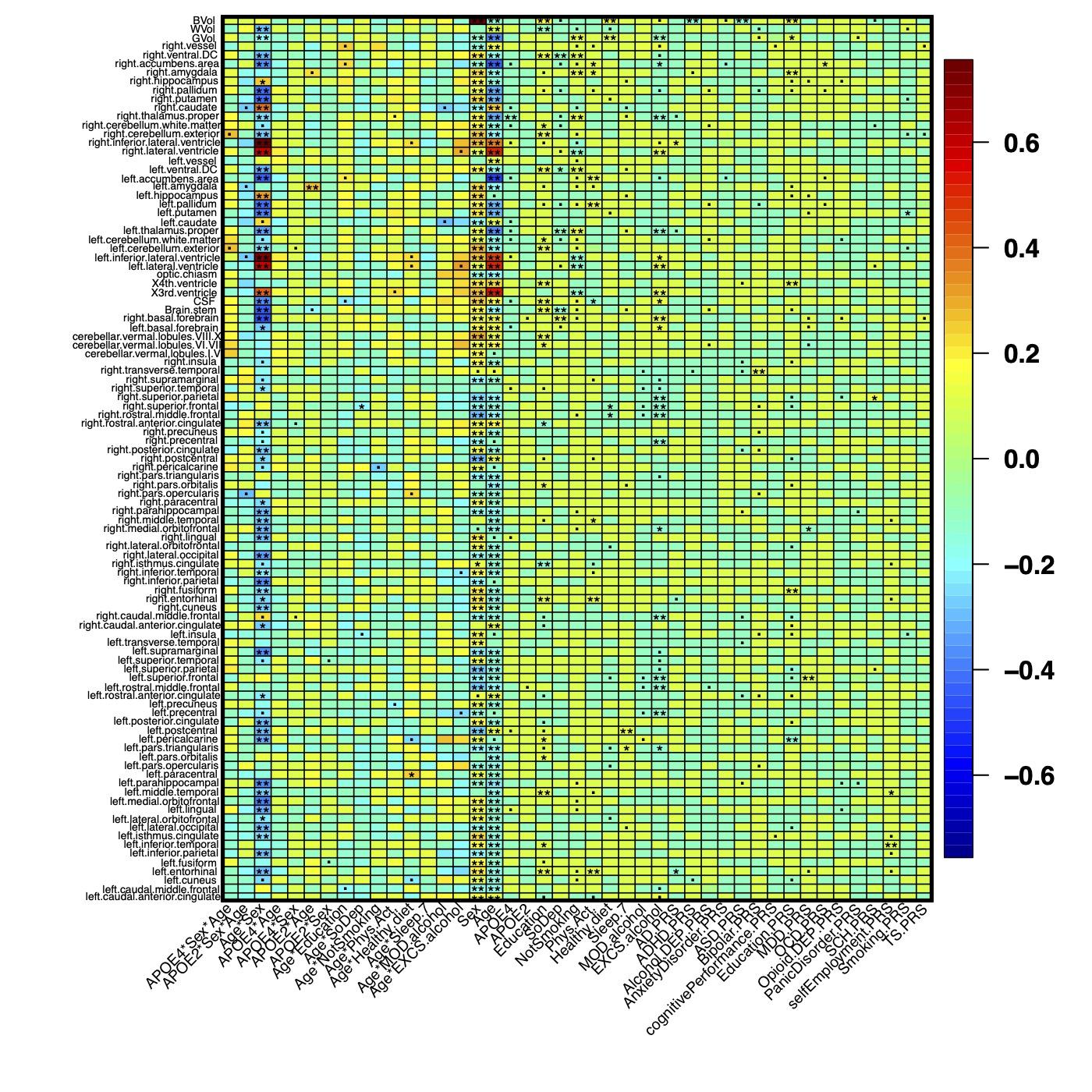

**Supplementary Figure S21.** Heatmap of the interaction effects of *APOE* and lifestyle factors on ROI volumes based on model P2 in **Table S2** (*APOE* as 2 categories). Results that passed *0.01*, *2.37E-4* and *4.74E-5* significance levels are denoted as (.), (*) and (**), respectively.

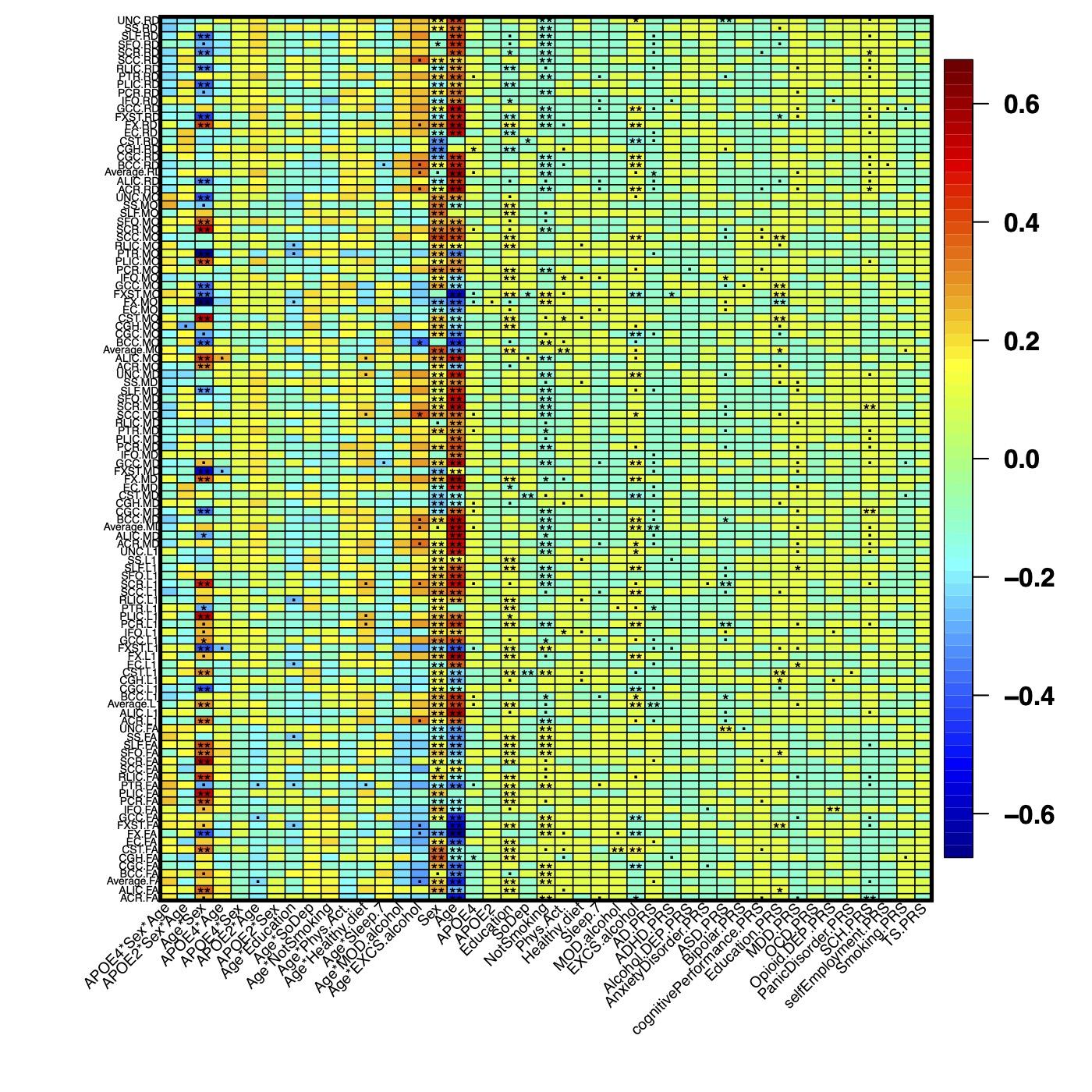

**Supplementary Figure S22.** Heatmap of the interaction effects of *APOE* and lifestyle factors on DTI measurements based on model P2 in **Table S2** (*APOE* as 2 categories). Results that passed *0.01*, *2.37E-4* and *4.74E-5* significance levels are denoted as (.), (*) and (**), respectively.

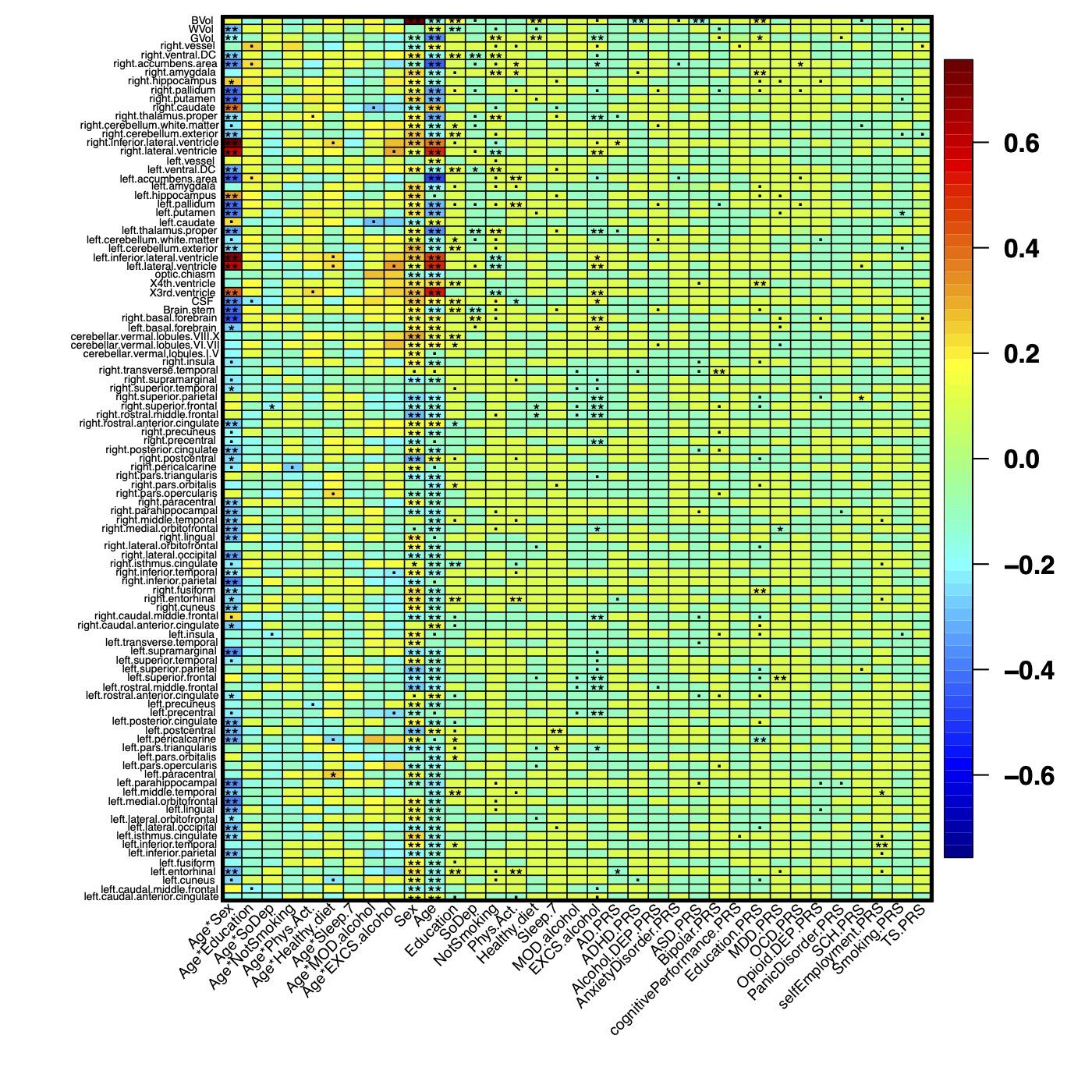

**Supplementary Figure S23.** Heatmap of the interaction effects of *APOE* and lifestyle factors on ROI volumes based on model P3 in **Table S2** (not adjusting for *APOE* terms). Results that passed *0.01*, *2.37E-4* and *4.74E-5* significance levels are denoted as (.), (*) and (**), respectively.

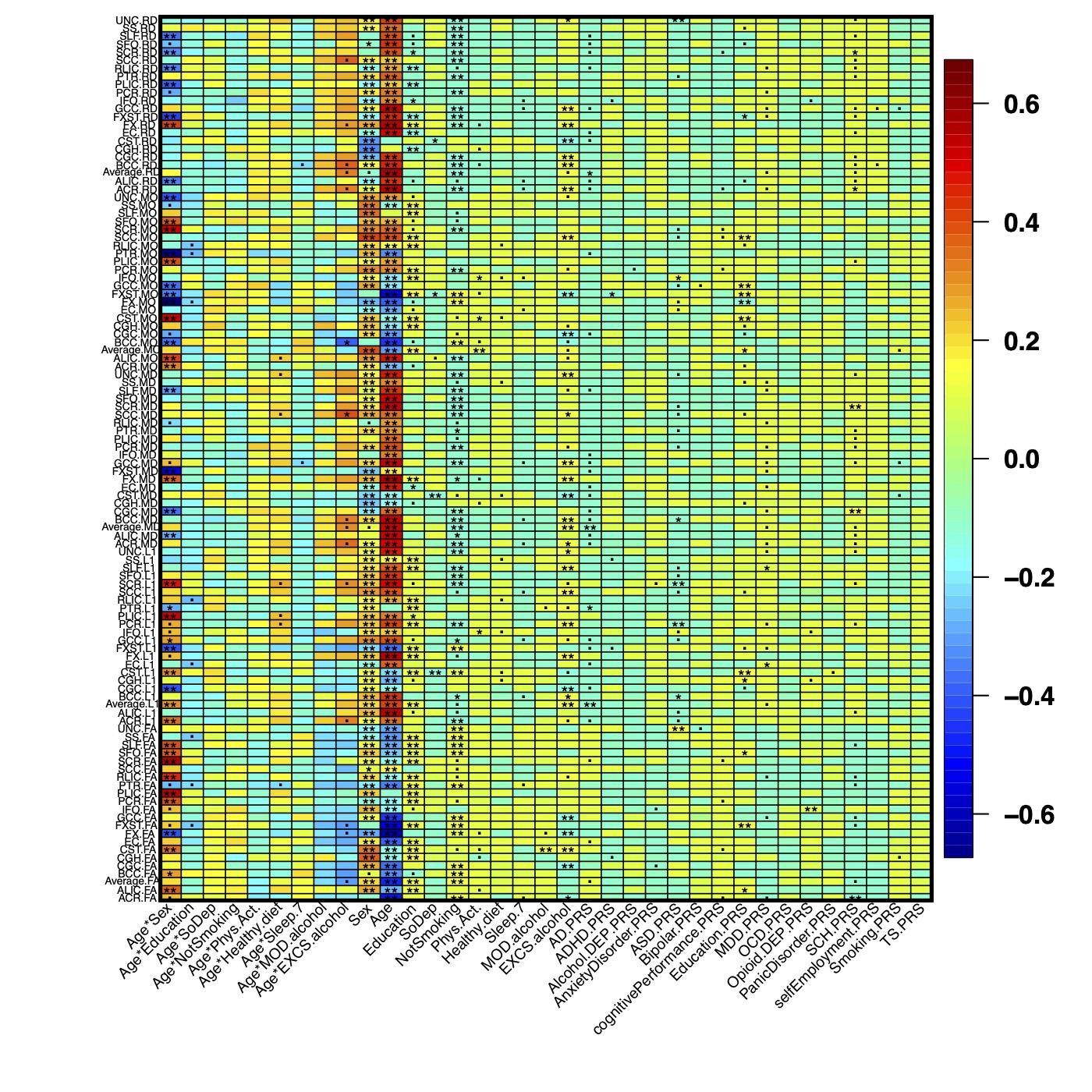

**Supplementary Figure S24.** Heatmap of the interaction effects of *APOE* and lifestyle factors on DTI measurements based on model P3 in **Table S2** (not adjusting for *APOE* terms). Results that passed *0.01*, *2.37E-4* and *4.74E-5* significance levels are denoted as (.), (*) and (**), respectively.

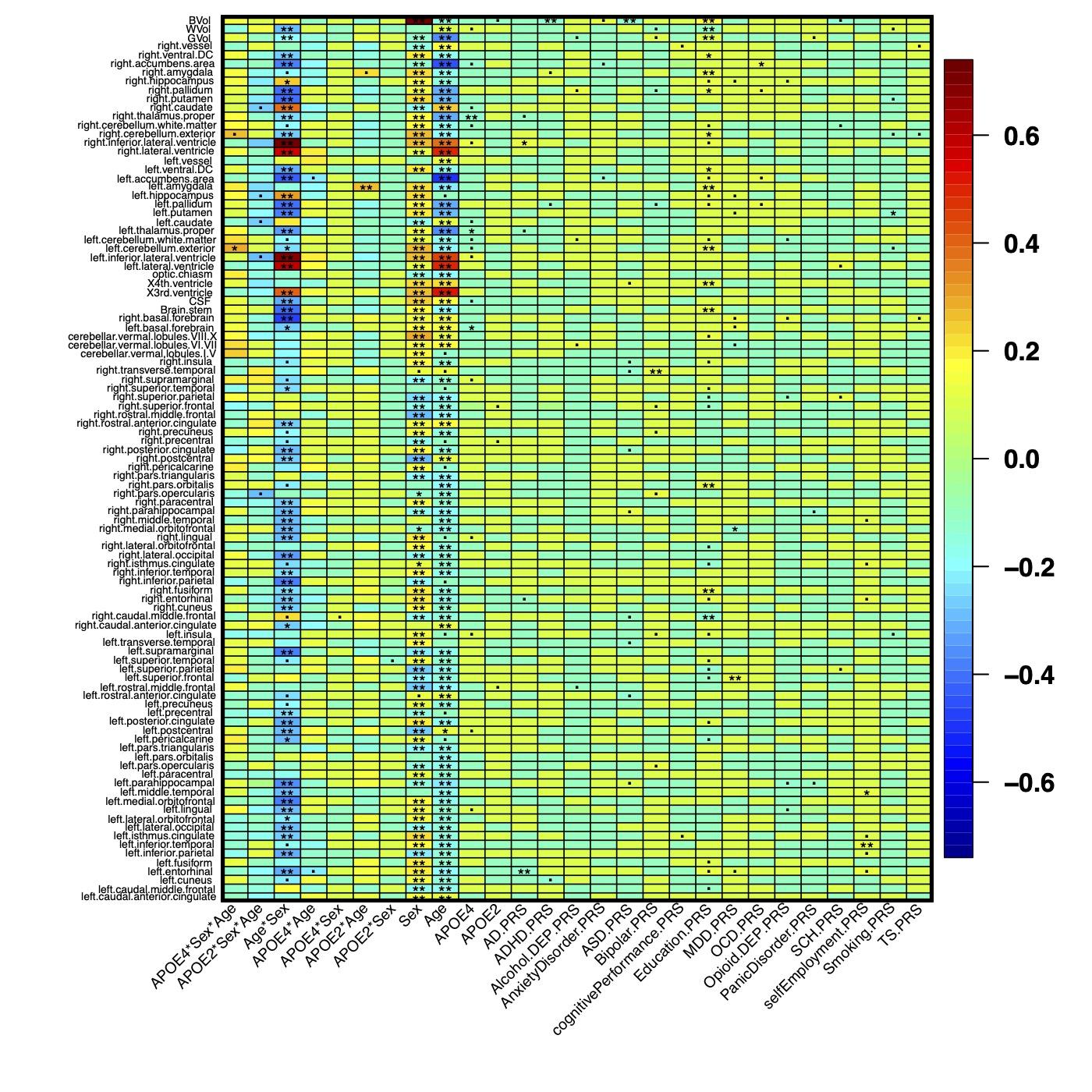

**Supplementary Figure S25.** Heatmap of the interaction effects of *APOE* and lifestyle factors on ROI volumes based on model P4 in **Table S2** (not adjusting for SES and lifestyle factors). Results that passed *0.01*, *2.37E-4* and *4.74E-5* significance levels are denoted as (.), (*) and (**), respectively.

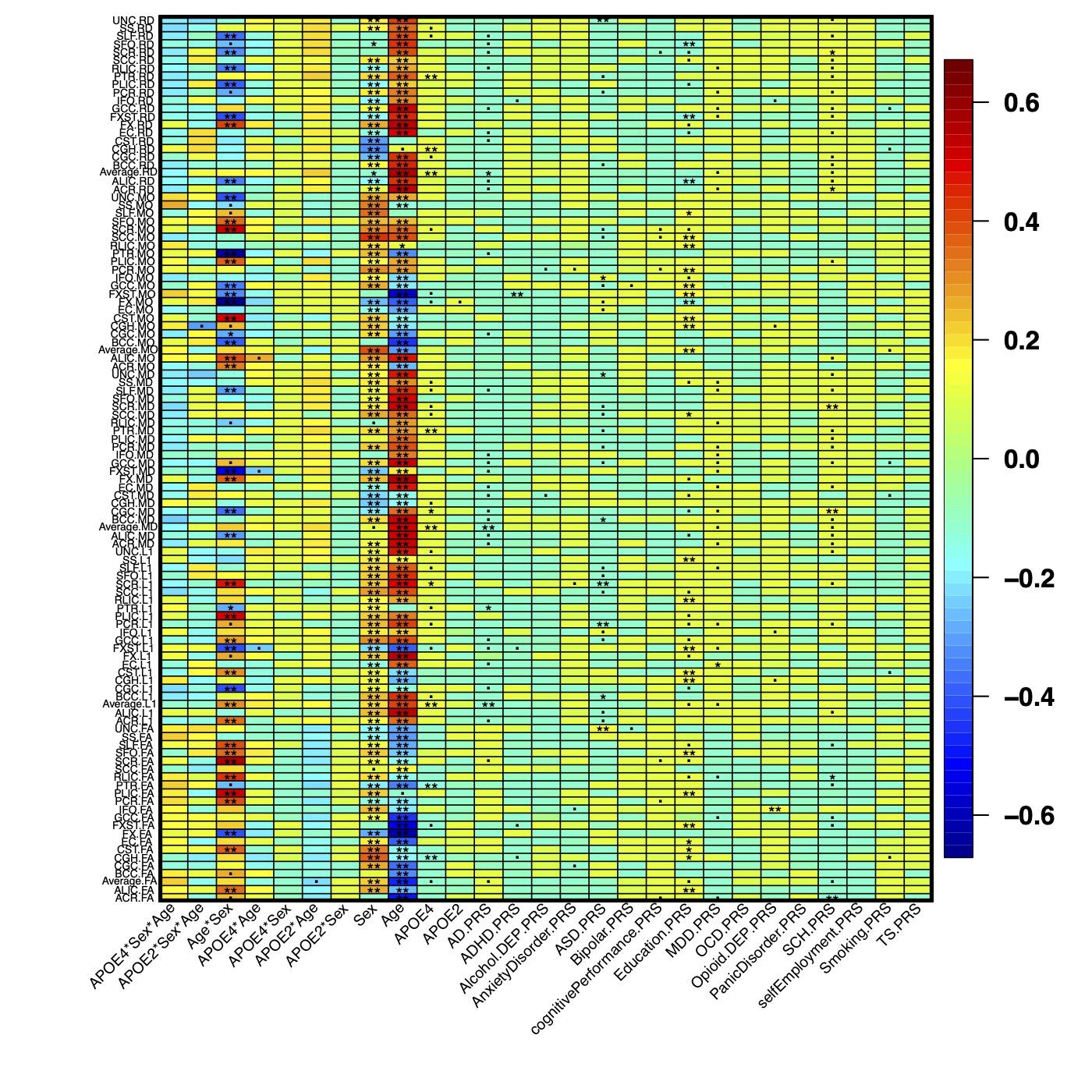

**Supplementary Figure S26.** Heatmap of the interaction effects of *APOE* and lifestyle factors on DTI measurements based on model P4 in **Table S2** (not adjusting for SES and lifestyle factors). Results that passed *0.01*, *2.37E-4* and *4.74E-5* significance levels are denoted as (.), (*) and (**), respectively.
